# Personalized Nutrition Recommendations Using a Bayesian Mixture Model of Concentration Constraints and Intake Preferences

**DOI:** 10.1101/2025.02.04.25321663

**Authors:** Jari Turkia, Ursula Schwab, Ville Hautamäki

## Abstract

Maintaining proper nutrition is crucial for preserving health and preventing disease. However, what constitutes proper nutrition may vary among individuals; evidence indicates that the effects of diet and even single nutrients can differ considerably because of personal characteristics. This personal variability can be observed through blood markers, such as concentrations of plasma cholesterol and insulin, and captured using a hierarchical multivariate model.

We leverage this variability and propose a conditional two-component Bayesian mixture model for generating personalized diet recommendations. The model uses the Nordic Nutrition Recommendations 2023 as a prior for healthy intake and infers individualized recommendations as posterior distributions. The first component identifies dietary options predicted to produce healthy levels across all considered blood markers, while the second selects, among these valid options, the diet closest to predefined personal preferences. The preference component is configurable and, in this study, was used to minimize dietary adjustments to support recommendation adherence while providing well-defined targets for nutrients less critical to concentration regulation.

The method was evaluated using nutritional data from two studies: one in prediabetic individuals and one in patients with kidney dysfunction. Numerical simulations showed that the individualized diets could restore or approach normal plasma concentrations when the estimated personal nutrient effects indicated biological feasibility. As the results align with current nutritional literature, the Bayesian approach offers a principled way to leverage observational nutrition data. However, future clinical studies are needed to validate the results and modeling approach before these can be translated into evidence-based personalized nutritional counseling.

## 1 INTRODUCTION

Proper nutrition plays a vital role in maintaining health and preventing disease ^1^. However, it has become increasingly evident that the impact of nutrition varies between individuals due to differences in nutrient metabolism and regulation ^2,3^. Personalized nutrition ^4^ aims to address this variability by providing personalized dietary guidance tailored to individual characteristics, going beyond generic recommendations ^3^.

Although nutritionists provide personal guidance, accurately determining individual responses and creating evidence-based diets remains challenging without rigorous statistical tools. Personalized guidance is particularly crucial for people with chronic diseases such as kidney dysfunction. For example, the National Kidney Foundation’s KDOQI guidelines recommend personalized dietary adjustments to maintain balanced serum concentrations, but leave implementation to nutritionists ^5^. Statistical models of individual responses could greatly improve the development of such personalized recommendations.

A notable example of personalized nutrition is the prediction of postprandial glucose responses (PPGR), where studies have shown that individual characteristics and meal composition can predict PPGR with high precision, outperforming generic diets in prediabetes and type 2 diabetes ^6,7,8^. However, these approaches focus only on a single response in glucose concentration and do not address the broader clinical need to balance multiple concentrations. This balance, clinically referred to as homeostasis ^9^, is essential to maintain physiological function. For example, people with kidney dysfunction require careful regulation of plasma phosphorus and potassium concentrations to preserve bone health, cardiac rhythm, and neuromuscular activity ^10^. Therefore, maintaining homeostasis remains fundamental, while personalized nutrition can also have other goals, such as weight loss or energy management ^11,3^.

A systematic review of machine learning approaches in personalized nutrition ^12^ showed that the methods focus mainly on meal classification without explicitly modeling the underlying nutrient-concentration relationships required for concentration balance. However, Bayesian hierarchical models ^13^ are already well-established in estimating subject-specific effects of nutrients in the N-of-1 intervention trials ^14^, where repeated measurements within individuals are used to estimate the personalized effects of nutrients. These designs are considered to provide stronger evidence for individual-level efficacy than conventional randomized controlled trials ^15^.

We generalize these intervention-based N-of-1 designs into an observational context by modeling individual responses from real-world data accumulated during routine eating behavior. This observational approach requires a less involved study design and supports iterative personalization from general guidelines to personal recommendations. Experimental intervention studies in vulnerable populations, such as renal patients, may pose ethical challenges ^16^ that observational studies can avoid. In addition to observational focus, we extend the N-of-1 methods by jointly modeling multiple response variables, such as plasma concentrations of phosphorus, potassium, and albumin. In real-world nutrition, a single nutrient can influence several physiological pathways ^17^.

Instead of classifying prepared meals, we estimate personal responses and ask what composition of nutrient intake would be optimal. This inverse formulation allows those joint response variables to be considered together and enables more dynamic and precise recommendations. An important inspiration comes from Gleason and Harris ^18^, who outlined how registered nutritionists can apply Bayesian reasoning to combine prior clinical knowledge with emerging evidence for informed decision making.

Our previous work demonstrated that Bayesian hierarchical models can effectively capture individual nutrient effects from observational data ^19^. Building on the biology of personal nutrition ^3^, we developed a model that uses nutrient intake as a predictor of plasma concentrations, with latent variables representing individual differences in nutrient absorption and metabolism. Using this framework, we proposed a Bayesian diet inference method that optimizes the intake of a limited set of nutrients while leaving the remaining nutrient intake of the diet unchanged ^20^. Simulations showed that this approach could effectively regulate plasma phosphate and potassium concentrations, preventing hyperphosphatemia and hyperkalemia in renal patients. However, these targeted adjustments were insufficient to achieve the desired concentration levels in all patients, underscoring the need for a more comprehensive approach and motivating the present study.

In this work, we generalize our previous recommendation method to address concentration control by inferring the most effective personal intake adjustments. The main contribution of this work is a Bayesian inference model that personalizes diet recommendations through posterior inference, using the Nordic Nutrition Recommendation 2023 (NNR 2023) ^21^ as a prior distribution of healthy intake. Our model identifies optimal intake levels that satisfy specified concentration constraints while aligning with individual dietary preferences. This is achieved through a conditional mixture: the first component prioritizes diets that satisfy the concentration constraints, and only among these valid options does the second component select the configuration that best matches personal preferences.

Concentration constraints are implemented using smooth sigmoid functions, enabling the model to tolerate invalid prior values and support gradient-based estimation methods. Sensitivity analyses of personal recommendations showed that such invalid values may occur at the edges of the intake ranges defined by general dietary recommendations (for example, NNR 2023). Initially, these recommendations caused the inference model to assign zero probability to certain diets, highlighting not only a computational challenge but also the limitations of broad dietary guidelines in accounting for individual variability. By smoothing the concentration constraints, the model was able to initialize from partially invalid intake values and iteratively adjust toward valid diets.

The preference component complements this by addressing identifiability issues: although multiple intake combinations may yield healthy concentrations, incorporating a clear preference establishes a well-defined mode in the posterior distribution. The preference definition can be modified, but here we seek minimal deviations from the current diet, assuming that small, yet effective adjustments enhance the practicality of personalized nutrition. The resulting mixture model seeks to align the expected values of the intake distributions with the preference, while the concentration constraints guide the posterior toward healthy concentrations, aiming to keep a sufficiently confident portion of the credible interval within acceptable ranges.

We demonstrate how personalized diets, guided by established dietary guidelines, can theoretically balance plasma concentrations in individuals with unhealthy levels. To evaluate this, we apply our inference model to two groups of patients: prediabetes and kidney dysfunction, using observational data from dietary records and corresponding laboratory analyzes. The simulation results show that our model can generate personalized diet recommendations that substantially improve the balance of key plasma concentrations with minimal dietary adjustments, supporting the feasibility of individualized dietary guidance. In most cases, plasma concentrations were simulated to reach their target ranges.

We adhere to the Bayesian Analysis Reporting Guidelines ^22^ (BARG), providing comprehensive details on data, code, models, computational processes, posterior distributions, inferences, and sensitivity analyses for transparency and reproducibility.

In summary, the key contributions of this work are:

- Demonstrating that a hierarchical multivariate model effectively captures individual variations in nutrient effects at multiple plasma concentrations from observational data.
- Developing a conditional two-component mixture model that integrates general nutritional guidelines with individual preferences to infer personalized dietary recommendations as posterior distributions.
- Validating the proposed method through simulations on two groups of patients, showing that in the simulation setting it can balance plasma concentrations within healthy ranges by leveraging the estimated nutrient effect functions.

## 2 THE INFERENCE PROCESS AND APPLIED NUTRITIONAL DATA

Our inference framework for personal diets consists of two main steps: first, constructing individual models that describe how nutrient intake relates to plasma concentration compositions; and second, inferring the nutrient intakes that are likely to modify those concentrations as desired. Based on our previous studies ^19,20^, we have prior evidence that the relationships between nutrient intake and concentration levels vary substantially between individuals. To reliably estimate these personal effects, we assume the data include repeated pairs of food records and laboratory analyses collected within a physiologically relevant time window, ensuring that the reported intake can plausibly influence the measured concentrations. We rely on the study design to provide such paired observations. In addition to repeated personal records, the data may include nested hierarchies, such as treatment phases or relevant groupings.

Beyond estimating nutritional effects, the observational data are also used to determine current intake levels, which later define personal preferences. During the inference phase, we do not use the observational data directly; instead, we assign prior proposal distributions to the inferred nutrients based on general dietary recommendations and derive their personalized posterior distributions. Figure 1 summarizes this process and the data used at each stage.

**FIGURE 1.**
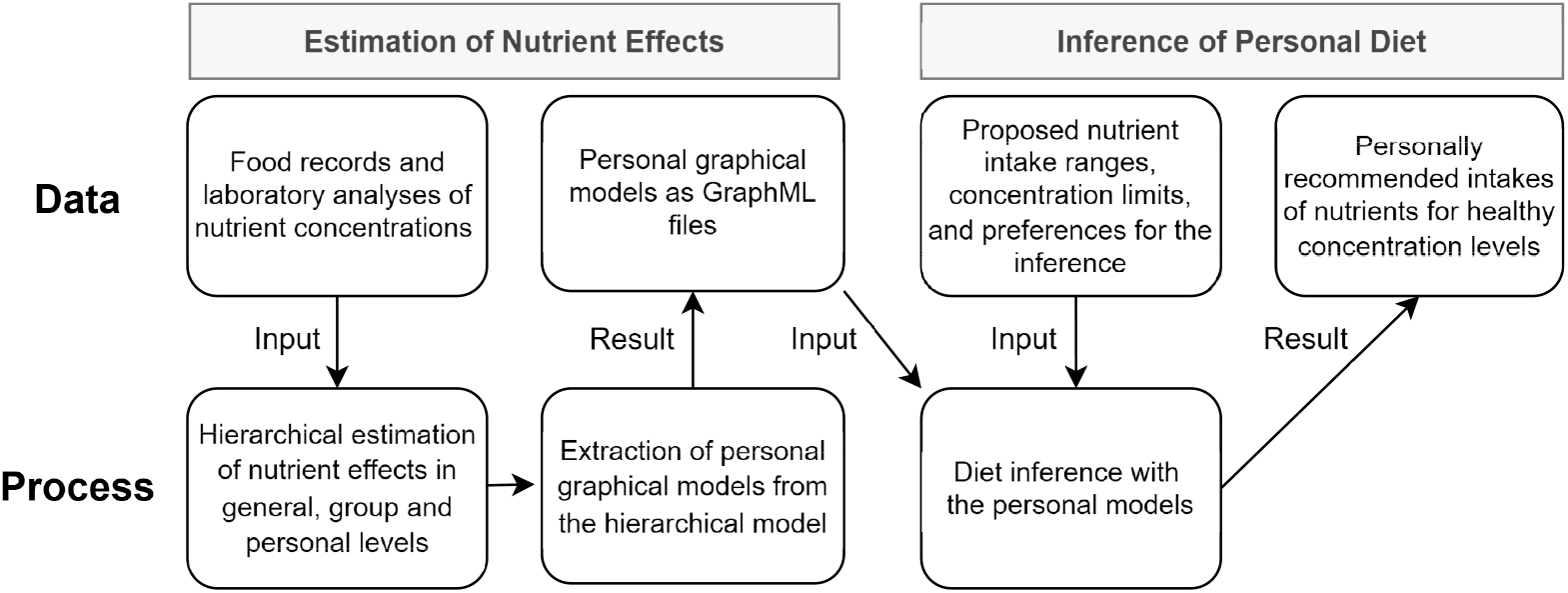
Two-stage process for inferring a personal diet. In the estimation stage, repeated food records and laboratory analyses are used to estimate nutrient effects at general, group, and personal levels, which are then summarized as personal graphical models. In the inference stage, these personal models are combined with generally proposed intake ranges and concentration limits, together with individual preferences, to generate personalized dietary recommendations that maintain concentrations within healthy ranges.

To evaluate the effectiveness of our method, we analyze two datasets that align with our described structure but involve different patients and variables. The first dataset, Sysdimet, originates from a dietary intervention study of the same name ^23^. This study included 106 subjects with impaired glucose metabolism and features of the metabolic syndrome. Data collection occurred at four time points over 12 weeks. At baseline, the subjects kept a 4-day food record and provided blood samples for laboratory measurements. At weeks 3, 7, and 11, new food records were kept, and follow-up blood samples were drawn within the subsequent week (weeks 4, 7, and 12), ensuring that each dietary record corresponded to a physiologically relevant response window. The primary goal of personalized nutrition in this context was to achieve reference values for specific fasting plasma concentrations, including high-density lipoprotein (HDL-C), low-density lipoprotein (LDL-C), total cholesterol (TC), insulin, and glucose. Predictors of concentration compositions encompass sex, cholesterol-lowering medication status, and 15 different nutrients, all detailed in Table 1. While the original Sysdimet trial randomly assigned subjects to groups to study the effects of different diets, our analysis treats subjects as independent, disregarding the randomization to groups.

**TABLE 1.**
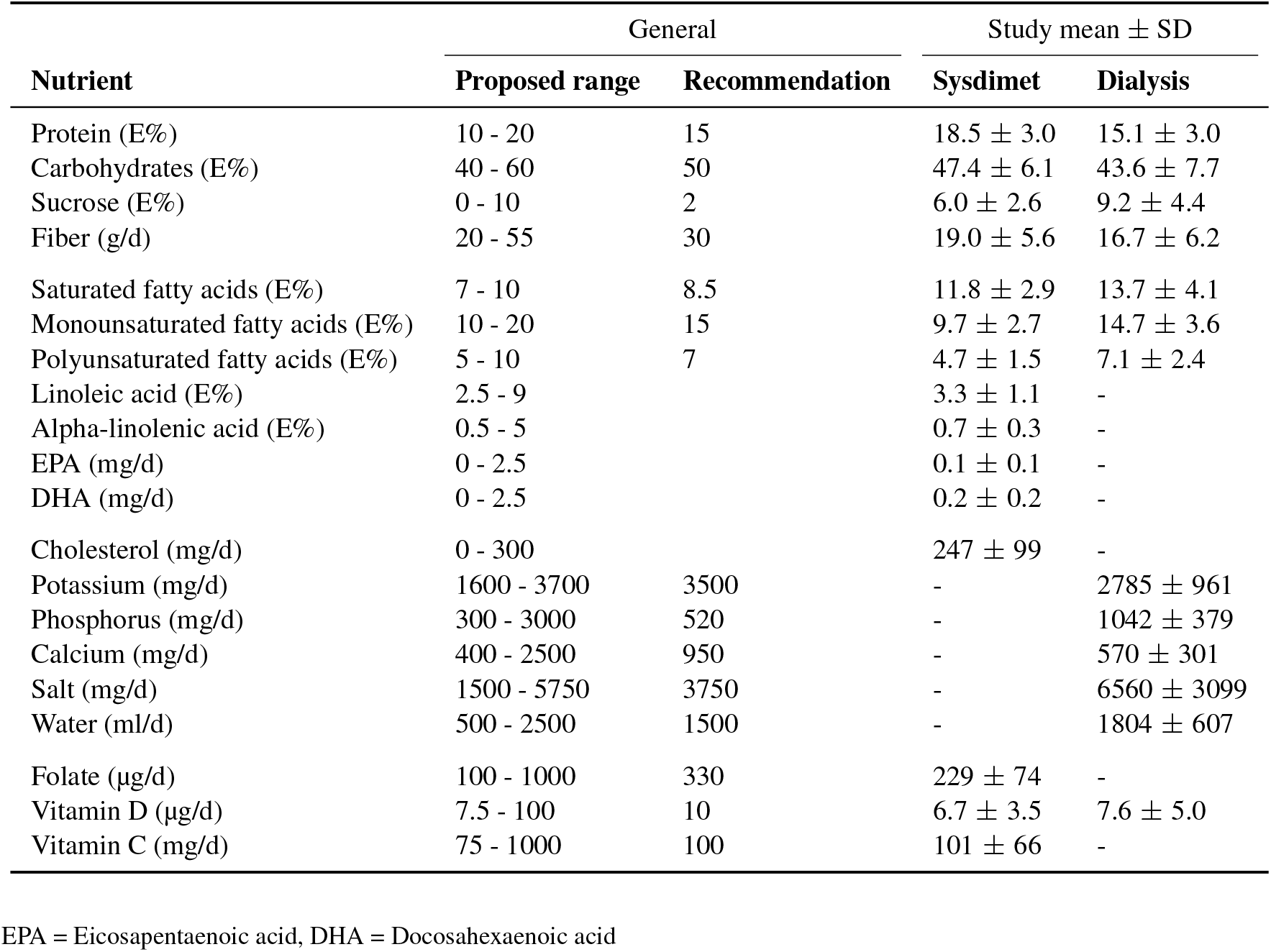
Nutrients analyzed and included in the inference of personal recommendations in one or both studied examples. The prior distributions for the nutrient random variables are implemented as a mixture of two distributions to balance between fully personalized recommendations and general guidelines. The proposed range, from 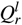 to 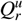, defines a uniform distribution, while a normal distribution centered at a specified general recommendation 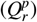 can be enforced with a hyperparameter *λ*_6_ to influence the degree of adherence to general recommendations and steer personal recommendations away from extreme values when necessary.

| Nutrient | General | | Study mean ± SD | |
| --- | --- | --- | --- | --- |
|  | Proposed range | Recommendation | Sysdimet | Dialysis |
| Protein (E%) | 10 - 20 | 15 | $18.5 \pm 3.0$ | $15.1 \pm 3.0$ |
| Carbohydrates (E%) | 40 - 60 | 50 | $47.4 \pm 6.1$ | $43.6 \pm 7.7$ |
| Sucrose (E%) | 0 - 10 | 2 | $6.0 \pm 2.6$ | $9.2 \pm 4.4$ |
| Fiber (g/d) | 20 - 55 | 30 | $19.0 \pm 5.6$ | $16.7 \pm 6.2$ |
| Saturated fatty acids (E%) | 7 - 10 | 8.5 | $11.8 \pm 2.9$ | $13.7 \pm 4.1$ |
| Monounsaturated fatty acids (E%) | 10 - 20 | 15 | $9.7 \pm 2.7$ | $14.7 \pm 3.6$ |
| Polyunsaturated fatty acids (E%) | 5 - 10 | 7 | $4.7 \pm 1.5$ | $7.1 \pm 2.4$ |
| Linoleic acid (E%) | 2.5 - 9 | | $3.3 \pm 1.1$ | - |
| Alpha-linolenic acid (E%) | 0.5 - 5 | | $0.7 \pm 0.3$ | - |
| EPA (mg/d) | 0 - 2.5 | | $0.1 \pm 0.1$ | - |
| DHA (mg/d) | 0 - 2.5 | | $0.2 \pm 0.2$ | - |
| Cholesterol (mg/d) | 0 - 300 | | $247 \pm 99$ | - |
| Potassium (mg/d) | 1600 - 3700 | 3500 | - | $2785 \pm 961$ |
| Phosphorus (mg/d) | 300 - 3000 | 520 | - | $1042 \pm 379$ |
| Calcium (mg/d) | 400 - 2500 | 950 | - | $570 \pm 301$ |
| Salt (mg/d) | 1500 - 5750 | 3750 | - | $6560 \pm 3099$ |
| Water (ml/d) | 500 - 2500 | 1500 | - | $1804 \pm 607$ |
| Folate (μg/d) | 100 - 1000 | 330 | $229 \pm 74$ | - |
| Vitamin D (μg/d) | 7.5 - 100 | 10 | $6.7 \pm 3.5$ | $7.6 \pm 5.0$ |
| Vitamin C (mg/d) | 75 - 1000 | 100 | $101 \pm 66$ | - |

EPA = Eicosapentaenoic acid, DHA = Docosahexaenoic acid

The second dataset (Dialysis) comprises data from 37 end-stage renal failure patients undergoing dialysis ^20^. The cohort includes 15 women and 22 men, aged 26 to 81 years (mean age: 61). For each patient, two observations were collected three months apart. Patients were interviewed about their dietary intake over the preceding 48 hours during each observation period, and laboratory tests were conducted within a week of the interview. For this analysis, we focus on three target concentrations from the laboratory tests: plasma potassium (P-K), fasting plasma phosphorous (fP-Pi), and plasma albumin (P-Alb). Our predictors for these concentrations include 13 dietary nutrients and several patient characteristics: sex, type of dialysis treatment, and current medications (including blood lipid medication, phosphate binder medication, diabetes medication, Renavit supplementation, and hydroxycholecalciferol supplementation). Table 1 presents the nutrients considered in both datasets, along with their mean intake and standard deviation.

The inference phase begins by establishing healthy ranges for adjustable nutrient intakes, as detailed in Table 1 for both datasets. These ranges are derived from the Nordic Nutrient Recommendations 2023^21^, which provide key reference values for each nutrient, including the recommended intake (RI) or adequate intake (AI), the average requirement (AR) or provisional AR, and the upper limit (UL). We aligned our ranges with the most appropriate lower bound depending on the nutrient, while the upper bounds generally followed the UL, representing the maximum intake typically tolerated without adverse effects. Although the primary objective is to determine personally optimal intake levels, these established ranges serve as essential constraints to ensure that recommendations remain within physiologically and clinically relevant limits. In addition to these thresholds, the inference model can be configured to favor general recommended levels, helping prevent extreme or impractical values in the final personalized recommendations.

Table 2 shows the normal ranges for the concentrations targeted in both studies. The baseline for these ranges follows Finnish national recommendations ^24^. However, the recommended upper limits for insulin (20.0 mU/l), LDL-cholesterol (3.0 mmol/l), and total cholesterol (5.0 mmol/l) were slightly relaxed, as several individuals in the Sysdimet cohort could not reach these thresholds in the simulated dietary adjustments. This exploration reflects insight into the physiological limits of personal dietary adjustment rather than a methodological shortcoming: given the estimated personal responses, no feasible modifications were sufficient to bring some individuals within the strictest concentration ranges. Additionally, fasting plasma glucose levels in this cohort tend to cluster near the upper boundary of the normal range, making the primary objective of our recommendation model to reduce glucose levels while maintaining other concentrations within their respective normal ranges. In contrast, the dialysis cohort presents a different challenge, with average albumin levels frequently falling below the normal range. Here, our model prioritizes increasing albumin levels while carefully managing potassium and phosphate concentrations to ensure overall balance.

**TABLE 2.** Targeted normal ranges for plasma concentrations may vary by age and sex. The problematic concentration limits for each study are highlighted in bold. Normal ranges follow Finnish national recommendations ^24^, except that the upper limits of insulin, LDL- and total cholesterol have been slightly relaxed in the Sysdimet cohort to allow for more feasible dietary adjustments.

| Plasma concentration | Target range | Target group | Study mean ± SD | |
| --- | --- | --- | --- | --- |
|  |  |  | Sysdimet | Dialysis |
| HDL cholesterol | 0.8 - 2.1 mmol/l | Men | 1.5 ± 0.3 | - |
| HDL cholesterol | 1.0 - 2.7 mmol/l | Women | 1.4 ± 0.4 | - |
| LDL cholesterol | 2.0 - **3.5** mmol/l | Everyone | 3.2 ± 0.8 | - |
| Total cholesterol | 3.9 - **5.5** mmol/l | Everyone | 5.2 ± 0.9 | - |
| Insulin | 2.6 - 25 mU/l | Everyone | 12.1 ± 6.3 | - |
| Glucose | 4 - **6** mmol/l | Everyone | **6.0** ± 0.5 | - |
| Potassium | 3.4 - 4.7 mmol/l | Dialysis patients | - | 4.4 ± 0.6 |
| Phosphate | 1.13 - 1.78 mmol/l | Dialysis patients | - | 1.66 ± 0.48 |
| Albumin | **36** - 48 g/l | 39 years or younger | - | **37** ± 0 |
| Albumin | **36** - 45 g/l | 40-69 years old | - | **34** ± 5.3 |
| Albumin | **34** - 45 g/l | 70 years and older | - | **32** ± 5.3 |

We assume that personal factors and multiple nutrients in current diets influence plasma concentrations. Although factors such as treatments, age, and sex remain fixed, nutrient intakes are adjustable within recommended ranges. These ranges, based on established guidelines, ensure that personalized recommendations address specific concentration challenges while staying within healthy limits.

## 3 PERSONAL DIET INFERENCE

The diet inference method aims to identify diet compositions that maintain all plasma concentrations within their normal ranges, with a given probability level denoted *c*. The process flow illustrated in Fig. 1 outlines the main computational steps. First, subject-specific relationships between nutrient intake and corresponding plasma concentrations are estimated using a multivariate hierarchical Bayesian regression. Next, personal graphical models are extracted from the hierarchical model, and finally, personal diet inference is performed on them. During inference, lower and upper limits, 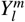 and 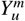, are imposed on all concentrations *m* = 1, …, *M*, and the most feasible nutrient intakes 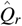 are inferred accordingly. For an individual *k*, this inference task is represented as a directed graphical model *G*_*k*_ in Fig. 2, following the established notation of Koller and Friedman ^25^. The graphical model provides a compact representation of personal nutrient responses and a structured framework for implementing the inference.

**FIGURE 2.**
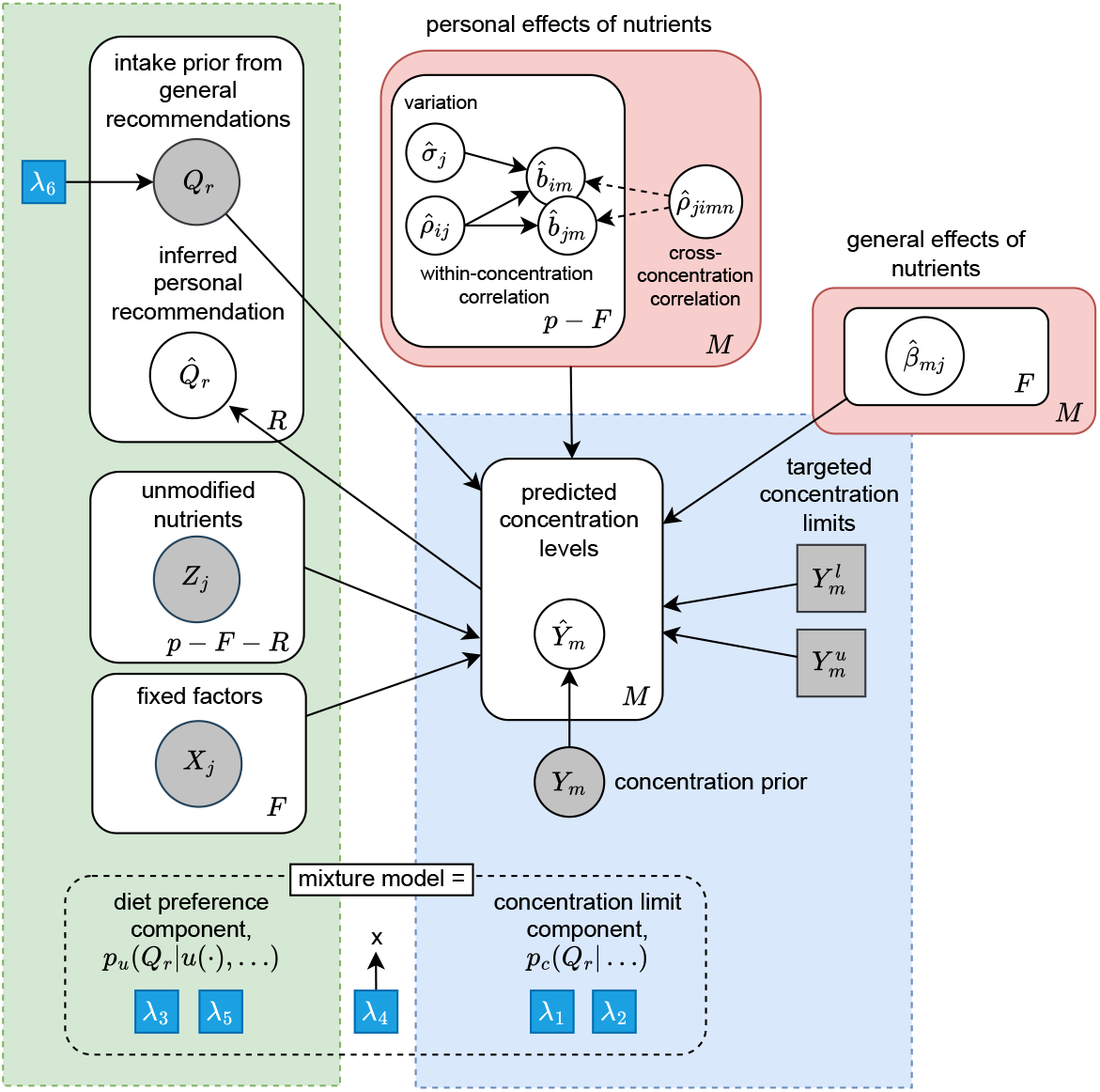
Graphical model 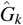 illustrating the formulation of personal nutrient intake recommendations 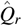 for individual *k*. These are inferred from a mixture model that balances two components: the concentration requirements (dashed blue box), which ensure predicted nutrient concentrations *Y*_*m*_ meet target limits 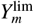, and the intake preferences (dashed green box), derived from personally adjusted general recommendations. The recommendations are based on the estimated effects of nutrients (pink box). Following conventional notation, circles denote random variables, squares indicate fixed hyperparameters of the recommendation process (*λ*) or defined limits, and plates show repetition across nutrients (*R*) or concentrations (*M*).

### 3.1 Hierarchical Model of Concentrations

Concentrations generally follow a strictly positive, right-skewed distribution, with higher values observed more frequently than lower values. In the nutrition literature, both Log − Normal ^26,27^ and Gamma ^28^ distributions have been applied to capture this pattern. In the presented model, the Gamma distribution was adopted because, when used with an identity link, it supports additive regression parameters and allows the direct interpretation of nutrient contributions to concentration levels.

Specifically, the *i*-th observation of concentration *m* for individual *k* is modeled as

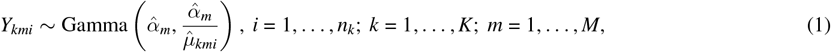

where the concentration shape parameters 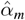 are estimated jointly with the regression effects, and the rate parameters are given by 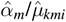, ensuring that the mean of the distribution equals 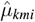. The expected values are obtained from a hierarchical linear predictor

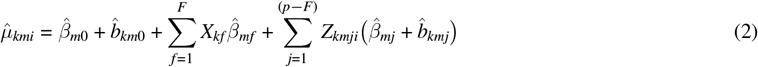

where *X*_*kf*_ denotes fixed personal factors (such as sex and baseline medication) that remain constant over time, while *Z*_*kmji*_ denotes observed predictors that vary across food record observations. The total number of predictors is *p*, consisting of *F* fixed predictors and (*p* − *F*) varying predictors.

The terms 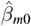 and 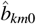 represent the population-level and subject-specific intercepts for concentration *m*, respectively. Subject-specific deviations 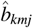 in concentration responses are estimated for nutrient intake, while fixed predictors enter only through fixed effects. An identity link is applied in the Gamma distributions linear predictor to preserve additivity and aid interpretability. All predictors are standardized before estimation, with both additivity and standardization facilitating consistent comparison across variables.

As established in physiology, the intake of a single nutrient can affect several physiological pathways simultaneously ^17^. This motivates not only the need to balance multiple concentrations in diet inference but also the need to model the effects of nutrients jointly rather than in isolation. For this reason, the concentrations are modeled as a system of seemingly unrelated regressions (SUR) ^29^. The SUR framework enables latent relationships between concentration responses to be captured even when their direct dependencies are not explicitly known. For example, it is not specified a priori that increasing one concentration would lower another, but such correlations can be inferred from the data.

In the classical SUR setting, cross-equation correlations arise through correlated error terms. In our hierarchical Bayesian formulation, however, cross-concentration dependence is introduced through subject-specific effects that follow a shared multivariate normal distribution,

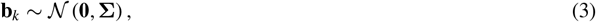

where the structure of the covariance matrix **Σ** captures also the cross-correlation effects. The matrix **Σ** has dimension *M*(*p* − *F* + 1), with *M* denoting the number of concentrations and (*p* − *F*) the number of predictors, including personal intercepts. It is parameterized using a diagonal matrix of standard deviations **T** and a correlation matrix **C**,

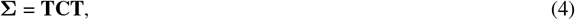

where **T** = diag(*σ*_1_, *σ*_2_, …, *σ*_*M*(*p*−*F*+1)_). The correlation matrix **C** captures the dependencies between subject-specific effects within and between concentrations: its diagonal blocks describe correlations within the same concentration *m*, and its off-diagonal blocks describe correlations between different concentrations *m* and *n* ^20^.

If group-level effects are relevant (e.g., treatment groups, dialysis modality), an additional nested set of random effects **g**_*g*_ can be introduced, defined on the same *Z* predictors with a separate covariance matrix **Σ**_*g*_. This extension yields a distinct variance-covariance structure at the group level, allowing shared variability to be captured both across concentrations and between groups. Alternatively, such group factors can be included as fixed effects in *X* (e.g. sex), while the random-effects specification is preferable when partial pooling between multiple groups is desired.

### 3.2 Recommendation Model for Nutrient Intake

For diet inference, the varying nutrient predictors of the linear predictor are divided into two parts: those nutrients that should remain fixed in the current diet and those that can be modified during inference. This separation allows the baseline contribution of the fixed factors and unmodified nutrients to be evaluated before inference, while optimization is directed only towards the modifiable part. Formally,

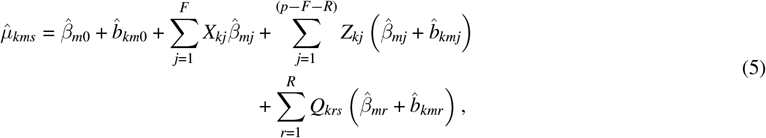

where the last summation corresponds to the *R* modifiable nutrient contributions, while the preceding terms remain constant during inference. The task of diet inference is then to adjust the intake levels *Q*_*krs*_ to achieve balanced concentration profiles. The sample index *s* = 1, …, *S* replaces the observation index *i* at this stage; the variables are no longer tied to specific food record measurements but instead represent draws from the posterior distribution optimized for dietary recommendations.

To formalize the conditional probabilistic dependencies between nutrient intakes and plasma concentrations in the graphical structure introduced in Fig. 2, we define the personal model for an individual *k* as a directed probabilistic graphical model ^25^,

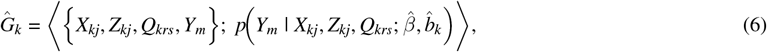

which corresponds to a system of seemingly unrelated regressions (SUR) for multiple concentration outcomes (*Y*_*m*_, *m* = 1, …, *M*). The personal recommendations are then obtained by conditional query of the graph under the imposed concentration limits,

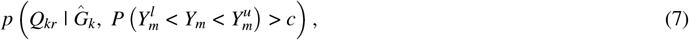

where the lower and upper limits 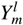 and 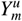 for each concentration *Y*_*m*_, *m* = 1, …, *M*, may vary according to age and sex, as shown in Table 2.

The healthy intake limits specified in general nutritional guidelines define the prior for recommended intakes *Q*_*r*_, ensuring that no recommendations are given outside these bounds. To balance fully personalized recommendations with general guidance, this support is combined with a normal distribution centered on the general recommended intake 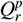:

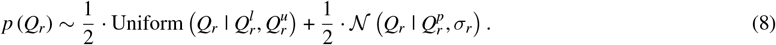

This mixture distribution keeps the recommendations within clinically healthy ranges 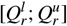 while allowing their alignment with the general guidelines to be adjusted.

There are several hyperparameters *λ* to adjust the posterior distribution of recommendations, indicated with blue squares in Fig. 2. In particular, the hyperparameter *λ*_6_ is used to scale the standard deviation of this distribution normal regardless of the length of the range of healthy intake of nutrients: 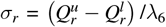. This formulation allows for flexible adjustment of the influence of general recommendations in all nutrients while maintaining the specific ranges of healthy intake for each nutrient.

The subject-specific effects of nutrients were estimated using a normalized intake scale, which also requires normalizing the proposed nutrient ranges by subtracting the mean and dividing the limits by the standard deviation. After inference, the original intake scales are restored in the nutrient recommendations.

#### 3.2.1 Implementing Confident Concentration Limits

Conditional concentration limits, 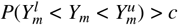, can be simplified by acknowledging that predictive uncertainties, given by the shapes of the concentration distributions, are already estimated in the graphical model 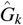. This allows for the inference of expected values that maintain the required cumulative probability within the original limits.

For 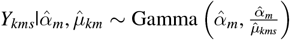, the expected value of the distribution satisfies 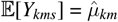. To adjust the limits of the expected values, we calculate the quantiles of the Gamma distribution using its cumulative density function (CDF) with

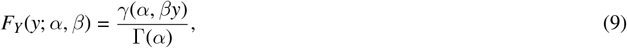

where Γ(*α*) is the gamma function of *α*, and *γ*(*α, βy*) is the lower incomplete gamma function. Using this CDF, we define the central quantiles corresponding to the desired confidence level *c* between the expected concentrations

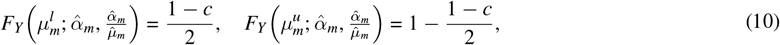

and the corresponding lower and upper expected concentration limits are then solved as

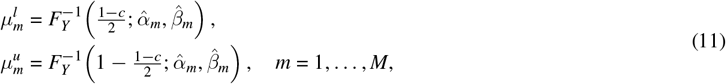

where 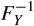 denotes the quantile (inverse CDF) function of the Gamma distribution. These adjusted limits constrain the expected concentration values 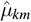 rather than the direct concentration values *Y*_*km*_, thereby simplifying the joint conditional distribution to

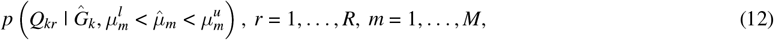

where 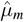 represents the expected concentration value of *Y*_*m*_, and the constraints 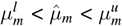 ensure the desired cumulative probability. This approach effectively balances predictive uncertainty and concentration constraints by using only expected values 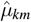.

#### 3.2.2 Robust Model for Multiple Limits

The implementation of strict concentration limits using hard constraints as in Eq. (12) can pose challenges for optimization algorithms, particularly those relying on gradient information ^30^. Furthermore, achieving the targeted concentration limits may not always be feasible with the initial proposal distributions of *Q*_*r*_, as their priors from general recommendations can be broad and may result in concentrations outside the limits for some individuals.

To address these problems, we implement the concentration limits with an inverse logit function that is differentiable and can be adjusted to favor normal concentration ranges while maintaining support outside these limits with

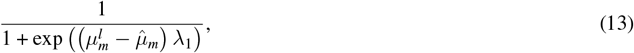

that smoothly transitions from 0 to 1 as the expected concentration value 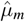 approaches the limit 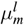. The hyperparameter *λ*_1_ controls the steepness of this transition, with higher values approximating a hard limit more closely. Figure 3 illustrates this concept. The blue curve represents the inverse logit function rising from 0 to 1 as 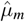 approaches the adjusted lower limit 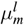, while the red curve shows a similar transition for the upper limit 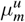 .

**FIGURE 3.**
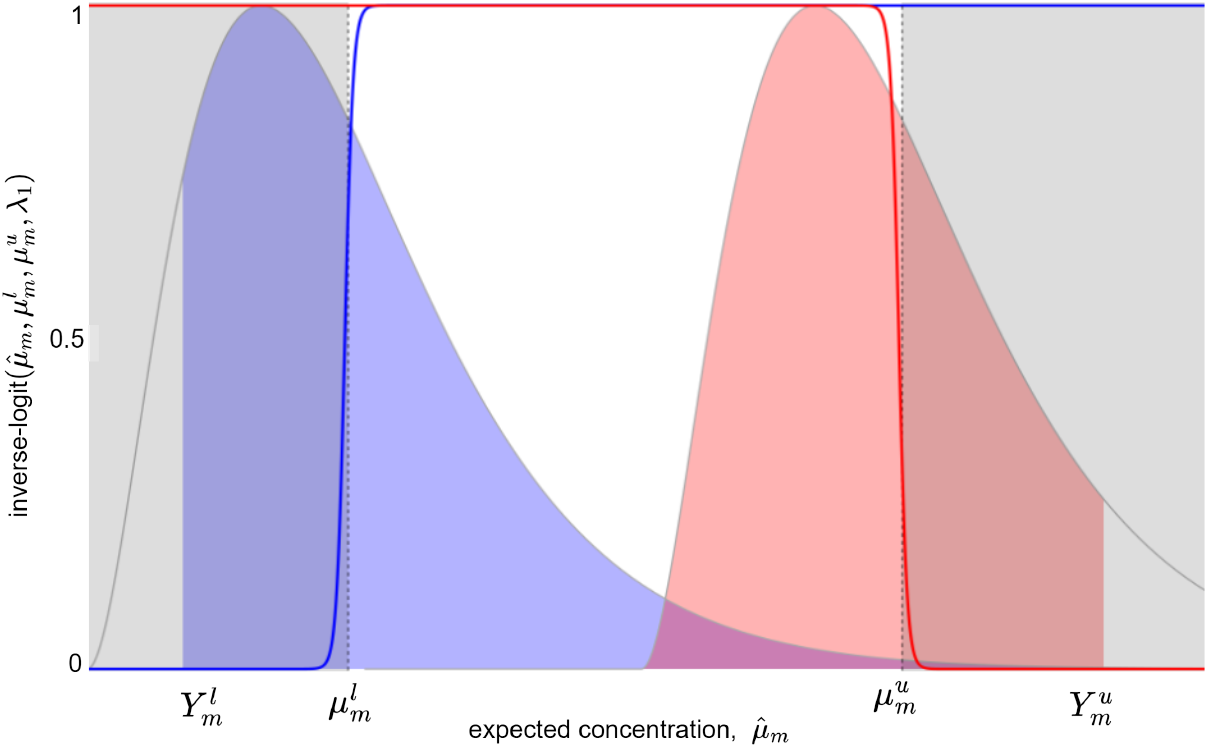
Illustration of transforming the original concentration limits 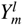 and 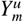 into corresponding expected values 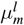 and 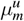 . The inverse logit functions (blue and red curves) provide smooth transitions at the limit values, facilitating differentiable optimization.

To quantify the degree to which both adjusted limits, 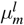 and 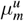, are met, we define a function

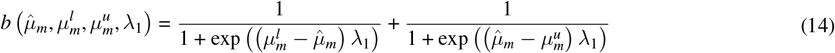

that increases smoothly as the targeted limits are reached and attains its maximum value of 2 when both limits are satisfied. The hyperparameter *λ*_1_ determines the steepness of the transitions and should be calibrated to ensure non-zero probability for initial proposal values.

We can then define a probability distribution associated with satisfying both limits of all the *M* concentrations as

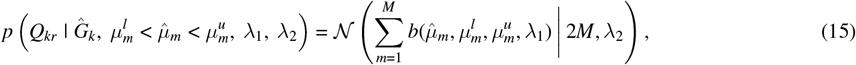

where the normal distribution with mean 2*M* reaches its maximum when all the limits are satisfied, and the hyperparameter *λ*_2_ as controls the allowed standard deviation from these limits. Although optimizing this distribution efficiently takes nutrient concentrations within the desired ranges, it can result in multiple equally valid diets. To resolve this ambiguity, we introduce a component of personal preference to guide the selection among these possibilities.

### 3.3 Mixture of Concentration Control and Personal Preference

Although many diet configurations can satisfy concentration constraints, this abundance of feasible solutions can make the inference distribution unidentifiable and complicate estimation. To address this, we use the concentration constraints from Eq. (15) as a primary component *p*_c_(·) of a conditional mixture distribution and introduce a complementary preference component that activates only after all target concentration limits have been satisfied. The preference component, denoted *p*_u_(·), is defined through a preference function *u*(·) that quantifies the degree to which a proposed diet aligns with a desired criterion. In this study, function *u*(·) is implemented in Eq. (18) as a measure of minimal deviation from the current diet, but the framework remains flexible for other preference functions in future applications.

With these definitions, the overall conditional mixture distribution becomes

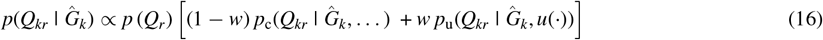

where *p* (*Q*_*r*_) is the prior intake distribution defined in Eq. (8). The coefficient *w* governs the conditional weighting of the mixture and is defined using the inverse logit function, analogous to the concentration limits,

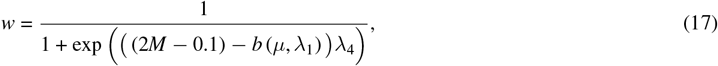

where *b* (*µ, λ*_1_) denotes the count of satisfied limits from Eq. (14) and 2*M* is the total number of limits. Subtracting 0.1 from this expected maximum ensures that *w* reaches 1 promptly once all limits are satisfied. The hyperparameter *λ*_4_ adjusts the steepness of this transition, providing a steep but smooth transition between the components of the mixture. The only requirement for the preference component *p*_u_ is that it has a unique maximum when the most preferred diet is achieved. This ensures that the posterior distribution has a single, well-defined mode corresponding to the optimal intake under the given preference.

#### 3.3.1 Personal Preference of Minimal Diet Modification

Although the definition of the preference component can vary, in this study it represents minimal deviation from the current diet to promote adherence to personalized recommendations. This objective is quantified through a preference function

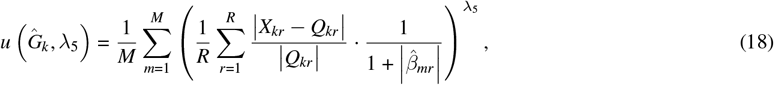

where *X*_*kr*_ and *Q*_*kr*_ are current and proposed intake levels, and 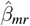 represents the effect of nutrient on concentration *m* of the personal model 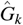 . The preference error is calculated as the relative difference between intake levels, scaled by the inverse effect of the nutrients. This scaling prioritizes minimal changes for nutrients with weaker effects while allowing greater adjustments for those with stronger effects, balancing dietary modification and concentration targets. To ensure consistency, the error is normalized by the number of nutrients (*R*) and the concentrations (*M*), making it independent of the size of the data set or the configuration of the model. The hyperparameter *λ*_5_ controls the sensitivity, penalizing larger deviations more heavily to enforce minimal changes to the current diet.

The preference component *p*_u_ should reach its maximum when the proposed intake *Q*_*r*_ aligns closely with the preference. This is modeled using an exponential distribution

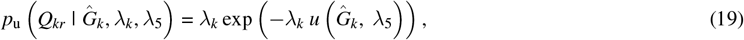

where *λ*_5_ globally shapes the preference for minimal changes, while *λ*_*k*_ adapts the scale for each individual *k*, as described in the next section.

#### 3.3.2 Determining a Safe Limit of Personal Preference Strength

As the proposed intake levels deviate further from the current values, the likelihood under the exponential distribution decreases and eventually approaches zero. To ensure that the model remains well-defined, the probability must stay positive and above a small threshold *ϵ* determined by computational precision. Maintaining this lower bound prevents numerical instability, which may otherwise occur when likelihood values approach zero.

We can calculate a safe threshold for the slope of the exponential distribution in Eq. (19) that still yields the likelihood *ϵ* at the maximum preference error. First, we calculate the maximum error *e* by comparing the current diet with the intake limits of each nutrient, following the same logic as in Eq. (19) with

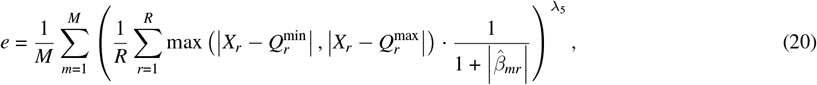

where 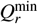 and 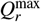 are the minimum and maximum recommended intake levels for nutrient *r*. Using this maximum error *e*, we can solve for the highest safe limit of *λ*_*k*_ that keeps the likelihood above *ϵ*,

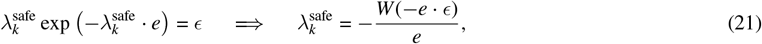

where *W* is the principal branch of the Lambert W function ^31^, applicable when the argument −*e* · *ϵ* ≥ −1/*e*, or equivalently *e* ≤ 1/*ϵ*.

Larger values of *λ*_*k*_ may still be preferred to emphasize personal preferences, though they can introduce computational instability if diet proposals approach intake limits. To provide flexibility, we introduce an adjustable hyperparameter *λ*_3_ that can add weight above the minimum level with

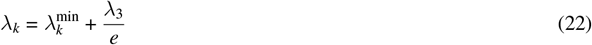

where *λ*_3_ can be directly added because the maximum error *e* is already normalized by the number of modeled concentrations (*M*) and nutrients (*R*). This formulation keeps the interpretation of *λ*_3_ consistent across different configurations.

If current nutrient intake *X*_*r*_ falls outside the range 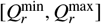, the resulting preference error necessarily exceeds the safe threshold defined above, as the most preferred intake cannot be achieved even when the likelihood remains above *ϵ*. Moreover, when personal preferences are strongly emphasized through large values of *λ*_3_, the resulting recommendations can become narrowly centered around the current diet, potentially biasing concentration balance and overlooking alternative dietary options that could yield better outcomes if more flexibility were allowed.

Together with the concentration component, this formulation of the preference ensures the numerical stability of the mixture model while seeking minimal dietary modifications within healthy concentration limits. Table 3 summarizes the model hyperparameters, their purposes, and guidelines for selecting appropriate values.

**TABLE 3.** Summary of hyperparameters used in the recommendation model with their default values for the analyzed data sets.

|  | Purpose | Value description | Defaults |
| --- | --- | --- | --- |
| $\lambda_1$ | Steepness of concentration limits (Eq. 14) | Moderately high value ensures steep transitions at concentration limits; lower to assign non-zero probabilities to initial proposals. | 8 |
| $\lambda_2$ | Concentration limits requirement (Eq. 15) | High value enforces strict adherence to concentration limits; adjust to relax or tighten requirement. | 15 |
| $\lambda_3$ | Personal preference strength (Eq. 22) | Values over 1 increase emphasis on personal preference $\lambda_k$; adjust to balance preference with flexibility. | 30 (10-40) |
| $\lambda_4$ | Transition steepness between mixture components (Eq. 17) | High value provides sharp transition between components; typically suitable, but lowering can help in divergences. | 30 |
| $\lambda_5$ | Preference error penalization (Eq. 19) | Values over 1 add penalization of deviations from current diet; adjust with $\lambda_3$ for desired balance. | 2.0 (1.0-2.0) |
| $\lambda_6$ | General recommendations weight (Eq. 8) | Higher values weight recommendations toward general guidelines, steering away from extreme intakes. | 10 |

## 4 IMPLEMENTATION

The proposed Bayesian models were implemented in the probabilistic programming language *Stan* ^32^. The implementation includes hierarchical models of nutritional effects for the Sysdimet and Dialysis datasets, and the diet inference model for personalized recommendations. The Stan code for these models is provided in the corresponding Supplemental Listings S1-S3. Both hierarchical models share the same core formulation based on the linear predictor at the personal-level in Eq. (2), while the Dialysis model extends this structure with an additional treatment-level hierarchy to capture variability between treatments. This extra layer expands the covariance structure, improving flexibility and precision in estimating treatment-specific effects.

After fitting the hierarchical models, the subject-specific nutrient effects and current intake estimates were extracted for each patient, following the previously described diet inference process. These were stored as personal graphical models in GraphML ^33^ files that provide a compact representation of individual responses. A detailed example of how these personal models were extracted and stored is provided in the Supplementary material for this section. Data management and analysis were performed in R-language ^34^ (version 4.5.1), with personal models constructed using the *iGraph* package ^35^ (version 2.1.4).

In addition to serving as a storage format, the extracted personal models support efficient sensitivity analyses of dietary recommendations without refitting the full hierarchical model. Sensitivity can be explored by modifying the hyperparameters summarized in Table 3, by adjusting the concentration limits, or by posing counterfactual scenarios by modifying the graph, such as “what if the intake of omega-3 fatty acids were increased?”.

The implementation emphasizes modularity, making the inference model adaptable to various sets of nutrients, dietary restrictions, and personal preferences. The preference error, defined in Eq. (19), was implemented as an adjustable Stan function to maintain numerical stability considering the maximum preference error in Eq. (20).

## 5 DATA APPLICATION

Data from the Sysdimet and Dialysis studies were used to estimate hierarchical nutrient effects. These estimated effects were then applied in the personal diet inference models to generate individualized recommendations. The analyses were conducted and reported in accordance with the Bayesian Analysis Reporting Guidelines (BARG ^22^), and all presented figures and posterior analyses were generated directly from the inference results.

### 5.1 Model Evaluation and Posterior Analysis

Posterior inference for both datasets was conducted in Stan using the No-U-Turn Sampler (NUTS) ^36^. Convergence was evaluated using rank-based 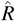 diagnostics ^37^ and effective sample sizes (ESS) following current Stan recommendations ^38^. According to the Stan Development Team and Bayesian Analysis Reporting Guidelines (BARG), rank-normalized 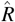 values below 1.01 indicate well-mixed chains, whereas values above 1.05 suggest clear non-convergence. Bulk and tail ESS are recommended to exceed 400 per chain to ensure reliable posterior estimates.

For the Sysdimet model, four parallel chains (4000 iterations each) produced well-mixed samples with 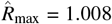 (mean 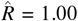) and bulk ESS values exceeding 800 for all parameters. No divergent transitions occurred, and the energy Bayesian fraction of missing information (E-BFMI) values (0.86-0.97) indicated efficient exploration of the posterior distribution.

The Dialysis model, which applied the same formulation with an additional nested group-subject hierarchy, was fitted with six parallel chains under otherwise comparable settings. Convergence was similarly satisfactory (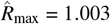, mean 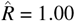), with a bulk ESS exceeding 1600 across parameters. Only 0.08 % of post-warmup transitions diverged, distributed sparsely across chains, and no truncation of treedepth was observed despite the more complex posterior geometry. The sampler was configured with a high target acceptance rate (*adapt*_*delta* = 0.985) and sufficient chain length to ensure the convergence of all parameters.

Predictive precision was evaluated using the normalized root mean square error (NRMSE; Supplementary Table S5). In Sysdimet data, HDL-C, LDL-C and total cholesterol achieved NRMSE values below 0.05, glucose showed the best fit (0.03), and insulin had a slightly higher error (0.14). In the Dialysis data, potassium and albumin had moderate NRMSE values (0.15 and 0.18), while phosphate displayed the largest discrepancy (0.31), reflecting greater variability in this response.

Visual posterior predictive checks (Supplementary Figs. S3-S4) showed close agreement between the simulated and observed data, indicating that the models reproduced the central tendency and variability of the observed concentrations without systematic bias. Most Bayesian posterior predictive *p*-values (Supplementary Table S6) were near 0.5, consistent with good calibration. Minor deviations in glucose (Sysdimet) and albumin (Dialysis) reflected expected skewness and measurement variability rather than structural model bias.

Overall, the diagnostics confirm that both models converged reliably and captured the main distributional features of the observed concentrations. Detailed diagnostic summaries, including trace plots and parameter-family statistics, are provided in Supplementary Figures S1 and S2 and Supplementary Tables S1 and S3.

Computation was performed on a laptop equipped with an Intel Core Ultra 5 135H processor (14 cores / 18 threads, 32 GB RAM) running WSL2 Linux under Windows 11 Enterprise. The Stan sampler utilized available CPU cores to execute chains in parallel, but no GPU acceleration was used. The Sysdimet model (four parallel chains) required on average 34109 s (9.5 h) for estimation, whereas the Dialysis model (six chains) required 62775 s (17.4 h). Subsequent personal recommendation inferences were computationally lightweight, averaging 1-3 s per individual. These timings are also reported in detail in the Supplementary Tables S2 and S4.

### 5.2 Diverse Personal Effects of Nutrients

The estimation of the hierarchical models revealed substantial individual variability in the effects of nutrients, which we analyzed using graphical models extracted for each patient. Supplemental Tables S7 and S8 summarize the 20 strongest subject-specific effects for both datasets, identifying the nutrients most influential in maintaining concentration balance.

Our objective in this analysis was to demonstrate the effectiveness of personalized dietary recommendations by focusing on patients whose current concentrations exceeded the recommended limits. The problematic limits are indicated in bold in Table 2, emphasizing their relevance for concentration control.

In the Sysdimet study, the main challenge was to reduce glucose and insulin concentrations within their target ranges. As summarized in Table S7, alpha-linolenic acid (ALA) exhibited the strongest decreasing effect on insulin, with minimal variation between individuals (ranging from -7.68 to -7.74), along with a minor increasing effect on glucose (0.41). This pattern is consistent with controlled studies in which ALA-rich oils such as rapeseed oil reduced fasting insulin and produced small increases in fasting glucose compared to other fats ^39^. In contrast, linoleic acid (LA) strongly increased insulin concentrations (7.56 to 7.60) but slightly decreased glucose (-0.43). Similar tendencies have been reported in dietary trials in which higher LA seed oils were associated with increased insulin release and modestly lower blood glucose levels ^40^. Vitamin C showed variable effects on insulin, ranging from a decrease of -0.29 to an increase of 1.03 in normalized magnitude. This variability is consistent with the wide range of insulin responses reported across randomized vitamin C trials, from decreases to increases depending on study population and dose ^41,42^.

In the Dialysis study, most patients faced difficulties in achieving albumin concentrations above the lower limit. This observation is in line with clinical guidelines that note that many dialysis patients struggle to maintain normal albumin levels due to inflammation, illness, and limited protein intake ^5^. Supplemental Table S8 summarizes the subject-specific effects of various nutrients, revealing substantial variability. For example, salt intake showed increasing effects on albumin ranging from -0.62 to 2.55, consistent with the well-known influence of fluid balance on dialysis: higher salt intake can dilute albumin, while better fluid control can increase its concentration ^43,44^. The effects of polyunsaturated fat (PUFA) on albumin ranged from - 0.41 to 1.32, which is consistent with clinical trials in which omega-3 supplementation produced mixed albumin results in different dialysis populations ^45,46^. Such opposing effects highlight the complexity of balancing concentrations, underscoring the importance of statistical inference to navigate these challenges.

### 5.3 Personally Inferred Dietary Recommendations

Following the diet inference process, personalized dietary recommendations were obtained for each patient by estimating nutrient intake levels that would keep plasma concentrations as close as possible to their healthy ranges. Each nutrient was given an allowed intake range based on general dietary recommendations (Table 1), and the target concentration ranges were taken from clinical reference limits (Table 2). Within these bounds, the model identified combinations of nutrient intakes most likely to improve the overall balance of concentrations, jointly steering them toward the recommended limits.

In Bayesian terms, the initial ranges of nutrient defined their prior distributions (Eq. 8), while the concentration limits served as probabilistic constraints in the conditional mixture model (Eq. 16). Only input data for this inference were the estimated nutrient effects of personal models, which describe how changes in intake are expected to influence concentrations. Using this information, the inference produced a joint posterior distribution over all nutrient intakes and their predicted concentration levels. From this joint distribution, individual posterior summaries were derived for each nutrient, representing the most probable intake levels that guide concentrations toward healthy limits while keeping the diet realistic for each individual.

Figure 4 shows results for five example patients from the Sysdimet dataset. The dashed vertical lines mark the current concentrations, and the solid lines indicate the expected concentrations after the recommended dietary changes. Each point in the distribution represents one feasible diet and its predicted concentration outcome, forming the posterior distribution of concentrations. Lighter colors indicate closer alignment with the current diet, reflecting the personal preference. The figure illustrates how the estimation process explores increasingly probable configurations of diets and concentrations, optimizing personal preference only after the concentration limits are satisfied.

**FIGURE 4.**
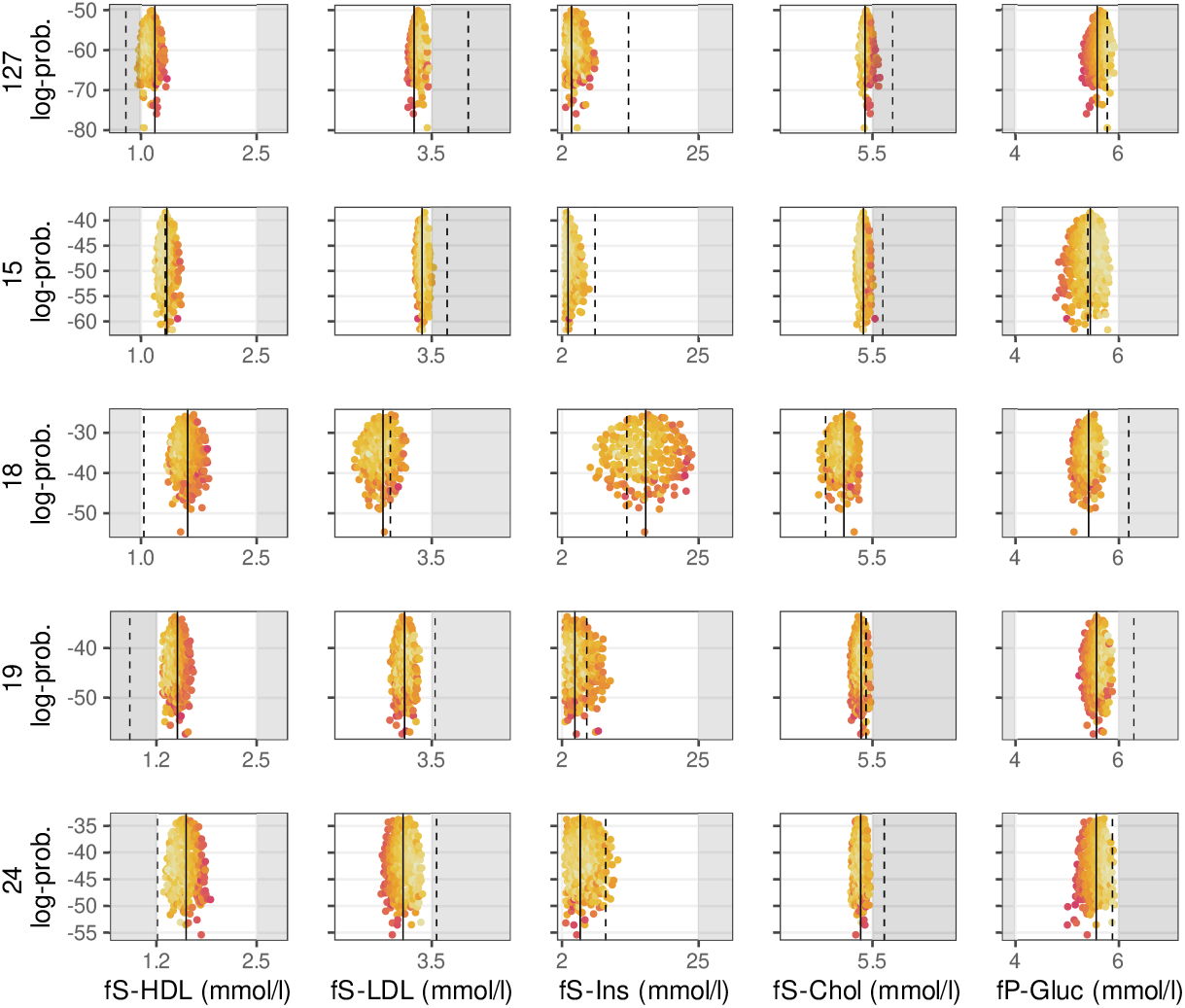
Example of five patients from the control cohort of the Sysdimet study, whose current concentrations deviate from normal ranges but are improved with personalized diet recommendations. The white areas represent the normal concentration ranges, while the dashed vertical lines mark the patients’ current concentrations, some of which fall within the gray areas. Each point within the red-yellow distributions represents an inferred diet proposal, with brighter yellow shades indicating closer proximity to the patients existing diet, which serves as a preference in the recommendation model. Because multiple diet configurations can produce the same concentration levels, points with different preference values can overlap. The solid lines indicate the concentration levels corresponding to the expected value of the joint multivariate recommendation distribution, where the nutrient intake and concentration predictions are jointly optimized. The diet side of this joint distribution reflects the personalized diet recommendations, ensuring that the expected concentrations are within healthy ranges. The figure was created using the ggplot2 package for R (v 3.3.5, https://ggplot2.tidyverse.org).

For most of the patients whose current concentrations were outside the recommended ranges, the model identified dietary adjustments that brought the predicted concentrations within or close to normal limits. For a few individuals, no single diet could satisfy all limits simultaneously, and in these cases the results represent the closest achievable balance under the model assumptions. For individuals already within the target ranges, the outcome depends on the hyperparameter *λ*_6_, which controls how strongly the general dietary recommendations are followed. When *λ*_6_ is set to zero, the model reproduces the current diet, whereas higher values shift the results toward general recommendations as long as the concentrations remain within healthy limits.

Table 4 summarizes the nutrient intake recommendations corresponding to the concentration predictions shown in Fig. 4. The reported values represent the expected (mean) intakes from the recommendation distributions, which correspond to the expected concentration levels indicated by the solid lines in the figure. The given credible intervals (CI) show the range of intakes still predicted to achieve the target concentration levels with 90% probability. The width of these intervals is clinically meaningful: narrow intervals indicate nutrients that must stay within a tight range to reach the targets, whereas wider intervals mark nutrients that allow more personal flexibility in following individual dietary preferences while maintaining balanced concentrations.

**TABLE 4.** Personally inferred intake recommendations for the five example patients from Sysdimet study in Fig. 4 whose current concentrations were out of the normal ranges.

| | Protein<br>(E%) | Carboh.<br>(E%) | Sucrose<br>(E%) | Fiber<br>(g/d) | SFA<br>(E%) | MUFA<br>(E%) | PUFA<br>(E%) | Linol. acid<br>(E%) | α-linol. acid<br>(E%) | EPA<br>(mg/d) | DHA<br>(mg/d) | Chol.<br>(mg/d) | Folate<br>(μg/d) | Vit. D<br>(μg/d) | Vit. C<br>(mg/d) |
| --- | --- | --- | --- | --- | --- | --- | --- | --- | --- | --- | --- | --- | --- | --- | --- |
| 127 | 17.2→<br>15.4<br>[10.5;19.8] | 54.7→<br>44.2<br>[40.2;51.9] | 8.1→<br>**0.8**<br>[0.0;2.6] | 16.6→<br>**22.6**<br>[20.1;29.3] | 10.2→<br>**7.6**<br>[7.0;8.8] | 7.4→<br>**18.9**<br>[16.1;20.0] | 2.9→<br>**7.6**<br>[5.2;9.9] | 2.4→<br>**3.5**<br>[2.5;5.9] | 0.4→<br>**4.9**<br>[4.5;5.0] | 0.0→<br>**1.8**<br>[1.5;2.2] | 0.1→<br>**0.1**<br>[0.0;0.3] | 230→<br>**48**<br>[1;159] | 149→<br>**968**<br>[898;998] | 4.8→<br>**18.8**<br>[7.8;37.0] | 97→<br>**100**<br>[76;157] |
| 15 | 17.3→<br>17.1<br>[11.6;19.9] | 42.4→<br>**51.1**<br>[40.9;59.6] | 5.4→<br>**3.5**<br>[0.1;9.2] | 17.7→<br>**24.2**<br>[20.1;34.5] | 10.1→<br>7.9<br>[7.0;9.5] | 8.9→<br>**18.7**<br>[15.6;20.0] | 5.1→<br>**6.3**<br>[5.0;9.0] | 4.1→<br>4.6<br>[2.6;8.3] | 0.8→<br>**4.7**<br>[3.9;5.0] | 0.0→<br>**0.4**<br>[0.1;0.9] | 0.1→<br>**0.2**<br>[0.0;0.6] | 232→<br>**91**<br>[3;263] | 252→<br>**430**<br>[153;773] | 6.7→<br>**13.3**<br>[7.7;27.0] | 151→<br>**456**<br>[110;916] |
| 18 | 20.1→<br>**14.9**<br>[10.3;19.6] | 42.9→<br>**49.9**<br>[40.7;59.2] | 4.2→<br>4.6<br>[0.3;9.6] | 17.0→<br>**37.2**<br>[21.0;53.9] | 14.0→<br>**8.5**<br>[7.1;9.9] | 9.4→<br>**14.8**<br>[10.3;19.7] | 4.4→<br>**7.5**<br>[5.1;9.9] | 3.1→<br>**5.8**<br>[2.7;8.8] | 0.9→<br>**2.5**<br>[0.6;4.8] | 0.1→<br>**2.2**<br>[1.6;2.5] | 0.2→<br>**0.5**<br>[0.0;1.3] | 229→<br>**146**<br>[8;292] | 203→<br>**481**<br>[118;947] | 4.1→<br>**61.7**<br>[14.4;97.9] | 48→<br>**517**<br>[99;974] |
| 19 | 18.8→<br>14.9<br>[10.3;19.7] | 47.4→<br>**48.5**<br>[40.3;58.9] | 5.8→<br>**2.8**<br>[0.1;8.4] | 18.0→<br>**31.4**<br>[20.4;51.2] | 13.4→<br>**8.4**<br>[7.1;9.9] | 8.8→<br>**16.7**<br>[11.2;19.9] | 4.7→<br>**7.3**<br>[5.1;9.8] | 3.3→<br>**5.5**<br>[2.6;8.8] | 1.0→<br>**3.9**<br>[1.9;5.0] | 0.1→<br>**2.2**<br>[1.7;2.5] | 0.2→<br>**0.3**<br>[0.0;1.0] | 233→<br>**131**<br>[6;288] | 233→<br>**674**<br>[312;969] | 5.9→<br>**85.0**<br>[52.2;99.5] | 106→<br>**210**<br>[79;507] |
| 24 | 18.1→<br>16.0<br>[10.6;19.9] | 38.7→<br>**51.3**<br>[40.9;59.6] | 7.5→<br>**4.3**<br>[0.2;9.5] | 12.5→<br>**31.9**<br>[20.5;51.7] | 19.8→<br>**8.2**<br>[7.0;9.8] | 11.8→<br>**16.6**<br>[11.0;19.9] | 3.9→<br>**7.7**<br>[5.2;9.9] | 2.5→<br>**5.3**<br>[2.6;8.7] | 0.5→<br>**4.1**<br>[2.3;5.0] | 0.0→<br>**1.4**<br>[0.6;2.3] | 0.1→<br>**0.3**<br>[0.0;0.9] | 339→<br>**100**<br>[3;267] | 247→<br>**863**<br>[560;996] | 4.8→<br>**57.6**<br>[13.3;96.5] | 91→<br>**381**<br>[87;894] |

As an example, patient 127 had an elevated LDL-cholesterol concentration. To lower it, the model recommended increasing the intake of both alpha-linolenic acid and folate (from 0.4 to 4.9 E% and from 140 to 961 *µ*g/d, respectively). These increases slightly raised glucose concentration, which was compensated by a higher EPA intake recommendation (from 0 to 1.8 mg/d). Alpha-linolenic acid and EPA had narrow credible intervals (4.6-5.0 E% and 1.5-2.2 mg/d), indicating their strong and well-defined effects on concentration control. This reasoning, together with the nutrient contribution patterns for all patients in the Sysdimet study, is illustrated in Fig. 5, and the corresponding contributions for the Dialysis study are shown in Fig. 6.

**FIGURE 5.**
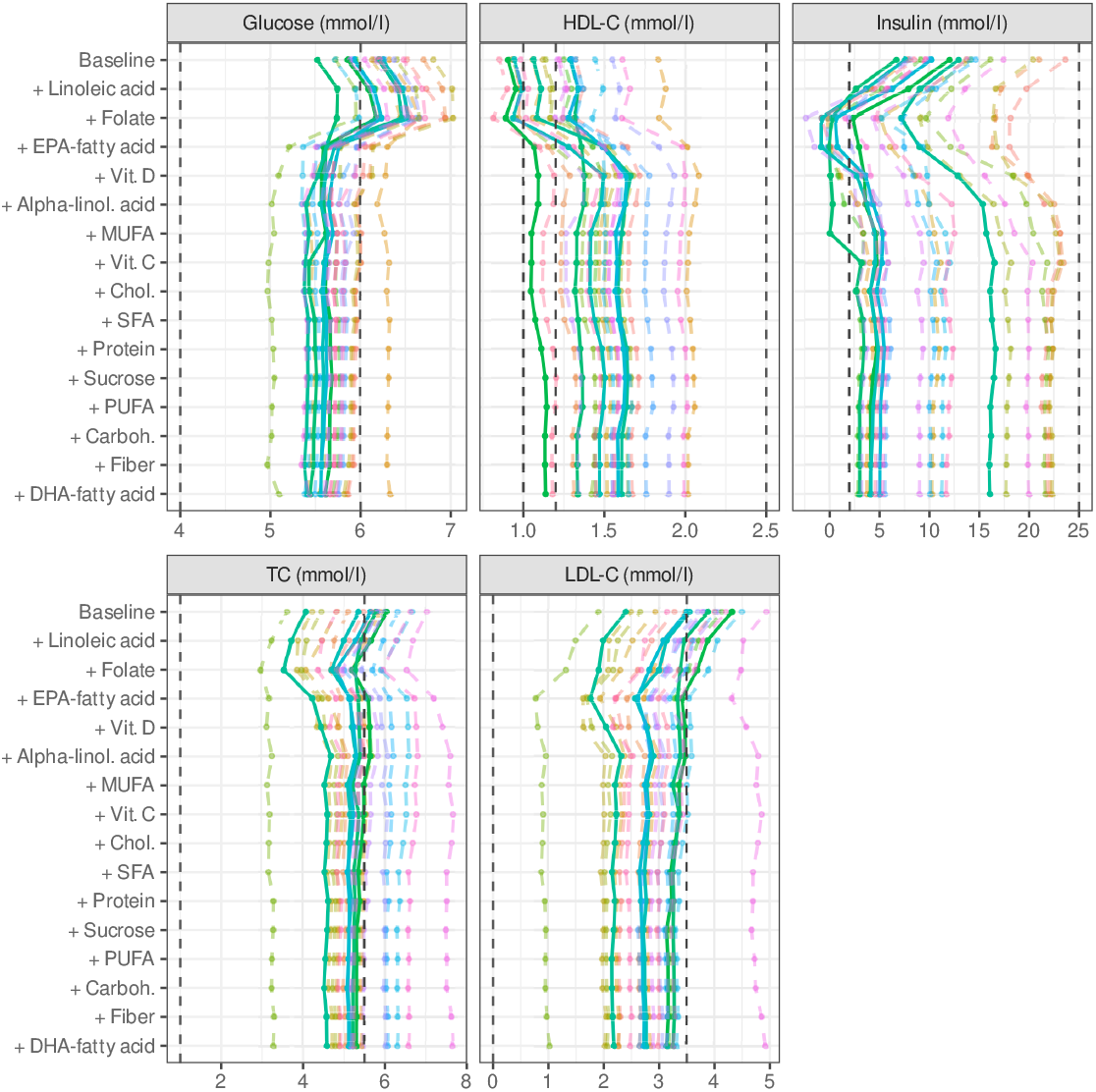
Figure shows personal compositions of concentrations for Sysdimet data set patients, starting with baseline concentrations and continuing with the contributions of each inferred nutrient intake. At the bottom, the final concentrations are predicted to reside within the targeted limits, as the dashed lines show. Nutrients are arranged in decreasing order of average contribution. Each colored line represents an accumulation of concentration for one patient. The solid lines correspond to the example patients in Fig. 4 and Table 4. The figure is plotted with ggplot2 package for R language (v 3.3.5, https://ggplot2.tidyverse.org).

**FIGURE 6.**
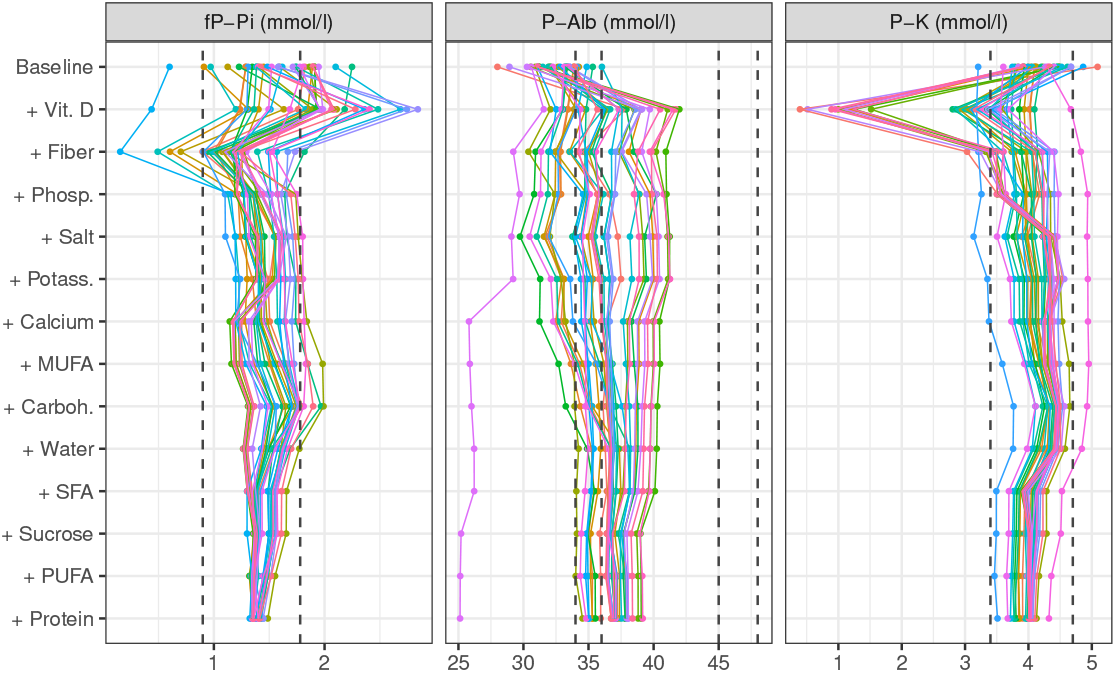
Figure shows personal compositions of concentrations for Dialysis data set patients starting with baseline concentrations and continuing with the contributions of each inferred nutrient intake. At the bottom, the final concentrations are predicted to reside within the targeted limits, as the dashed lines show. Nutrients are arranged in decreasing order of average contribution. The figure is plotted with ggplot2 package for R language (v 3.3.5, https://ggplot2.tidyverse.org).

Comprehensive results are provided in the Supplementary Material. Supplemental Fig. S5 presents the concentrations achieved for the remaining patients in the Sysdimet study, with the corresponding intake recommendations listed in Table S9. In the Dialysis study, 35 of 37 patients initially had concentrations outside the normal ranges. Supplementary Fig. S8 shows examples of concentration distributions achieved for a subset of these patients, and Supplementary Table S10 provides the corresponding intake recommendations for all patients with concentrations outside the target ranges.

The table presents the current levels of nutrients intake from patients, the inferred recommendations and the intervals 90% of the recommendations. The increase in current intake is highlighted in red, and the decrease is highlighted in blue. Bold font highlights over 30% change in the current intake. (DHA = Docosahexaenoic acid; EPA = Eicosapentaenoic acid; MUFA = Monounsaturated fatty acid; PUFA = Polyunsaturated fatty acid)

## 6 DISCUSSION

Personalized nutrition requires methods that can account for individual variability in nutrient responses and balance multiple plasma concentrations simultaneously, an aspect often overlooked by existing approaches. In this study, we addressed this gap by developing a hierarchical multivariate model combined with a conditional two-component mixture framework. This approach integrates general nutritional guidelines with individual dietary preferences to infer posterior distributions of personalized dietary recommendations. By validating the method through simulations on two cohorts of patients, we demonstrated that the model can generate feasible dietary adjustments that restore plasma concentrations to healthy ranges while minimizing changes in current diets.

Although our simulated results are promising, validating these personalized dietary recommendations would require clinical interventions, which have not yet been performed. The absence of ground truth data makes it challenging to compare directly with other recommendation methods. To establish the validity of our approach, we evaluated the alignment of our model’s nutrient effect estimates and intake recommendations with established findings in the nutrition literature, finding a good agreement with current nutritional understanding.

### 6.1 Evaluation of the Inferred Recommendations

Supplementary Table S13 compares our inferred effects of nutrients and intake recommendations from both data sets with existing nutritional literature. Our analysis identifies two distinct nutrient groups: those with consistent directional effects (increasing or decreasing concentrations) that vary only in magnitude between individuals and those with mixed effects that vary in direction between individuals. Although the literature supports our findings for nutrients with consistent effects, mixed effects are less documented in existing research. The alignment of the model with well-established nutritional effects supports its validity, and identifying personally varying effects can reflect limitations in traditional statistical approaches rather than model shortcomings.

Our results demonstrate that the Bayesian framework offers advantages in personalized nutrition modeling. The transparency and interpretability of our customized Bayesian model are evident in our analysis of the contributions of nutrients to plasma concentrations. Figures 5 and 6 illustrate these contributions, ranked by impact magnitude. The figures show how recommended nutrient intakes adjust concentration levels from their baseline values (set by the fixed and unmodified predictors) to achieve expected values according to Eq. (2), providing clear insight into the model’s decision-making process.

The resulting concentration levels reflect complex interactions among the effects of nutrients, personal baselines, and current dietary patterns, rather than simple nutrient-concentration relationships. In the Sysdimet cohort (Fig. 5), the model identified key contributors to insulin concentration, including EPA, folate, alpha-linolenic acid, linoleic acid, and vitamin C. EPA exhibited substantial individual variability with strong increasing effects in some patients, while alpha-linolenic acid showed a more consistent and moderate positive impact. Linoleic acid consistently decreased insulin concentration, while vitamin C showed varying effects, ranging from decreases to increases between individuals. Detailed numerical evidence for these contributions is provided in Supplemental Tables S11 and S12. In particular, these computationally inferred effects, derived from the observational Sysdimet data, agree well with the results confirmed in the intervention cohorts of the same study ^23^. Furthermore, the model indicated a general tendency towards a lower intake of saturated fatty acids and cholesterol for most participants, which is consistent with current nutritional guidelines ^21^ and based purely on observed data. This agreement shows that the model can identify clinically relevant dietary patterns even from limited observational information.

Protein is vital for renal patients due to its positive effect on albumin (-0.05 to 0.45) and its consistent impact on insulin lowering (-0.05 to -0.37). Similarly, sucrose, which increases albumin (0.65 to 2.31) but decreases potassium and insulin, is typically reduced to balance these concentrations. Carbohydrates have moderate effects, supporting adjustments towards a 50% E intake to reduce insulin while increasing albumin and potassium levels ^47,48^. Nutrients with less pronounced effects, such as fiber and water, remain near general recommendations unless their current intake exceeds the suggested ranges.

Our model provides nuanced recommendations for nutrients with complex effects, illustrated by MUFA and PUFA. MUFA exhibits varying individual-specific impacts on insulin (-0.09 to 0.81) and albumin (0.17 to 1.39). However, the model recommends increasing intake towards 15 E%, which aligns with current dietary guidelines ^21^. Similarly, while PUFA generally lowers fasting plasma insulin concentration, its effects on albumin vary substantially between individuals (-0.41 to 1.32), highlighting the importance of personalized recommendations.

The smooth and differentiable concentration limits in our mixture model (Eq. 16) enable meaningful recommendations even when fully optimal solutions are not achievable. For example, Fig. 5 shows patient 55 (pink line), for whom the model could not identify a diet that would reduce LDL and total cholesterol concentrations below their targeted upper limits. Although these particular targets could not be reached given the estimated personal responses and the modeling assumptions, the model still provides recommendations that successfully bring insulin and glucose concentrations within their respective healthy ranges.

### 6.2 Limitations of the Method

Although our Bayesian model provides a robust framework for balancing nutrient intake with plasma concentration targets, several limitations should be acknowledged. First, the model assumes that changes in plasma concentrations are directly attributable to nutrient intake, which may oversimplify complex physiological processes and overlook non-dietary factors. The observed variations in individual dietary responses, captured as latent variables, likely reflect differences in genetics, metabolism, lifestyle, and environmental factors ^3^. Future work could explore more flexible model formulations or optimization strategies to assess whether feasible or more optimal solutions exist for individuals for whom the current model yields only partially attainable recommendations.

To simplify the analysis of limited nutritional data, the model assumes that the expected concentrations are a linear combination of the intake of nutrients and the effects of nutrients. Although actual biological reactions are often non-linear and more complex ^9^, this linear additive assumption applies only to the expected concentration values, allowing a straightforward interpretation of nutritional effects. The model accommodates natural variability in observed concentrations through a Gamma distribution, maintaining analytical simplicity while reflecting inherent biological fluctuations.

Flexibility in the model is achieved through several hyperparameters that control concentration limits and weighting of personal preferences (Table 3). Although these hyperparameters improve adaptability, they also require careful tuning to ensure an appropriate balance between meeting concentration targets and generating realistic dietary recommendations. Sensitivity analysis is essential to select suitable values.

In cases where concentration limits cannot be fully met, the model can produce recommendations that are computationally effective but not biologically optimal. For example, controversially high intake suggestions for vitamins C and D were observed in subjects S18, S19, S104, S107, and S121 (see Supplemental Fig. S6). In these cases, the estimated insulin-lowering effect of vitamin C was used as a last resort for individuals struggling to meet the upper glucose and insulin limits. Although these recommendations are computationally justified, alternative clinical strategies may be more appropriate.

### 6.3 Reliability of the Analysis

The model diagnostics confirmed reliable convergence for both the Sysdimet and Dialysis analyzes. The relatively small number of observations contributed to wider credible intervals for some coefficients of the effect of nutrients, reflecting greater uncertainty in these estimates. This was most evident for nutrients with high individual variability, such as EPA and vitamin C, where the credible intervals remained broad despite good convergence metrics.

The reliability of the resulting recommendations depends more on the quality and variability of the observations than on their absolute count. In an observational N-of-1 setting, the range of nutrient intake arises from natural dietary variation rather than a controlled design. When this range is narrow, estimated effects remain uncertain regardless of sample size. From a Bayesian perspective, the relevant criterion is whether the available data allow the conditional recommendation distribution (Eq. 7) to be estimated with sufficient precision to support a dietary change. This reflects a precision-based view of sample size ^22^, where adequacy is judged by how precisely the model can predict meaningful outcomes rather than by a fixed number of observations. In our work, two pairs of records in the Dialysis study provided only limited evidence for a dietary change, whereas four pairs in the Sysdimet study that covered a broader intake range substantially improved precision.

In practice, the main limitation in nutrition research is the availability of repeated pairs of intake and concentration records rather than the computational burden of model estimation. In our experiments, the amount of data had only a minor effect on the estimation time. Halving the number of observations in the hierarchical Sysdimet model reduced runtime only slightly (from 9.5 h to 9.0 h), whereas halving the MCMC chain length nearly halved it (to 5.0 h). Runtime therefore depends more on model complexity and the required chain length to achieve convergence than on the amount of data. The two-level Dialysis model, which included additional random-effect hierarchies, was markedly slower (17.4 h) despite fewer observations, showing that structural complexity rather than data quantity drives computational cost. Longer chains can nevertheless be necessary when the model structure is more complex or when higher effective sample sizes are required for stable inference.

Overall, the computational demands of Bayesian estimation remain modest compared with the effort required to obtain high-quality data. Increasing the number and diversity of observations is the most effective way to improve the precision of nutrient effect estimates, provided that the underlying data structure is faithfully represented in the model. Consequently, study design should prioritize data quality and coverage, while computation remains a manageable and secondary consideration to ensure reliable Bayesian recommendations.

## 7 CONCLUSIONS

Our results demonstrate that the proposed Bayesian inference model effectively captures the interplay among personal concentration baselines, nutritional effects, and current dietary intake. This enables personalized dietary strategies that align individual health needs with general nutritional guidelines. By balancing multiple plasma concentrations with personal dietary preferences, the model provides actionable individual recommendations that prioritize concentration homeostasis while accommodating secondary goals such as weight loss or minimal dietary change.

Using observational data, we replicated the health benefits of omega-3 fatty acids and vitamin C in improving glucose metabolism and lipid profiles, findings previously confirmed in the Sysdimet study ^23^ through controlled nutritional interventions. This consistency highlights the potential of observation-based dietary guidance as a practical and efficient alternative to resource-intensive intervention studies, bridging the gap between research and real-world application. Furthermore, the Bayesian frameworks capacity to quantify uncertainty ensures robust, evidence-based recommendations, advancing personalized nutrition as a promising approach to improving public health.

## Data Availability

All data produced in the present study are available upon reasonable request to the authors

## Supporting information

Supplementary document provides code listings, figures, and tables referenced in the main article, together with additional methodological details. Its contents are organized to follow the sections of the main manuscript.

## Data Availability Statement

The data supporting the findings of this study are available from the corresponding author upon reasonable request.

