## Supplemental material for "Personalized Nutrition Recommendations Using a Bayesian Mixture Model of Concentration Constraints and Intake Preferences"

#### Contents

This Supplementary Material provides the listings, tables, and figures that complement the main manuscript with extended discussions and detailed results. The material is organized in sections corresponding to those of the main paper to facilitate easy cross-referencing between the main text and the supplementary analyzes.

##### Listings

- **Listing S1.** Stan implementation of the personal nutrient effect model for Sysdimet data (hierarchical multivariate regression).
- **Listing S2.** Stan implementation of the personal nutrient effect model for Dialysis data (two-level hierarchical multivariate regression).
- **Listing S3.** Stan implementation of the diet inference model (mixture of concentration control and preference).

##### Tables

- **Table S1.** Family-wise sampler diagnostics for the Sysdimet case.
- **Table S2.** Hierarchical model estimation times for the Sysdimet case.
- **Table S3.** Family-wise sampler diagnostics for the Dialysis case.
- **Table S4.** Hierarchical model estimation times for the Dialysis case.
- **Table S5.** NRMSE values for predicted plasma concentrations.
- **Table S6.** Posterior predictive $p$-values for the Sysdimet and Dialysis models.

[*]School of Computing, University of Eastern Finland, Joensuu, Finland

[†]CGI Suomi Oy, Joensuu, Finland,

[‡]School of Medicine, Institute of Public Health and Clinical Nutrition, University of Eastern Finland, Kuopio, Finland

[§]Department of Medicine, Endocrinology and Clinical Nutrition, Kuopio University Hospital, Wellbeing Services County of North Savo, Kuopio, Finland

- **Table S7.** Strongest and personally most varying effects of nutrients and medication in the Sysdimet data set.
- **Table S8.** Strongest and personally most varying effects of nutrients in the Dialysis data set.
- **Table S9.** Personally inferred intake recommendations for Sysdimet control cohort.
- **Table S10.** Personally inferred intake recommendations for Dialysis patients.
- **Table S11.** Most personally decreasing and increasing nutrient contributions in the Sysdimet data set.
- **Table S12.** Most personally decreasing and increasing nutrient contributions in the Dialysis data set.
- **Table S13.** Comparison of inferred nutrient effects and intake recommendations with literature.

**Figures**

- **Figure S1.** Trace plots for parameters with largest $\hat{R}$ (Sysdimet model).
- **Figure S2.** Trace plots for parameters with largest $\hat{R}$ (Dialysis model).
- **Figure S3.** Posterior predictive checks for Sysdimet data.
- **Figure S4.** Posterior predictive checks for Dialysis data.
- **Figure S5.** Remaining Sysdimet patients with personalized recommendations.
- **Figure S6.** Elaborated effects of vitamins C and D in Sysdimet study.
- **Figure S7.** Selected Dialysis patients with improved concentrations under personalized diet recommendations.

### Section "4 Implementation"

Listing S1: Stan implementation of the personal nutrient effect model for Sysdimet data with hierarchical multivariate regression.

```
// -----------------------------------------------------------------------------
// Effects of Nutrition - Hierarchical Gamma GLMM (identity link)
// -----------------------------------------------------------------------------
// * Likelihood: y_n ~ Gamma(alpha[m], beta_n) with rate beta_n = alpha[m]/mu_n
// * Link: Identity, with E[y_n] = mu_n on the outcome scale.
// * Positivity enforced via smooth softplus around response-specific floors
// * Random effects at subject (s) with Z structure having full cross-response
//   correlation (Cholesky factors).
// * QR reparameterization for fixed effects (no intercept in QR)
// -----------------------------------------------------------------------------

functions {
  // Softplus with scale s > 0 and per-response floor (>0)
  // keeps the implied Gamma mean strictly positive while preserving
  // identity-link behavior away from the floor.

  real softplus_scaled(real x, real s, real floor) {
    return s * log1p_exp((x - floor) / s) + floor;
  }
}

// -----------------------------------------------------------------------------
// DATA
// -----------------------------------------------------------------------------
data {
  int<lower=0> N;                     // number of observations
  int<lower=1> p;                     // number of fixed-effect predictors
                                      // (incl intercept in X[,1])
  int<lower=1> v;                     // number of responses
  int<lower=1> n_s;                   // number of subjects in data
  int<lower=1> k;                     // number of subject-level predictors
                                      // (incl intercept in Z[,1])

  int<lower=1,upper=n_s> subject[N];  // subject index per row

  matrix[N, p] X;                     // fixed-effect design
                                      // (X[,1] is intercept; we drop it in QR)
  matrix[N, k] Z;                     // random-effect design (Z[,1] is subject intercept)

  // Array-of-vectors: one vector of length N per response
  vector[N] Y[v];                     // responses (must be > 0 for Gamma)

  // Per-response floors for clamping mu to maintain positivity of the Gamma rate
  real<lower=0> mu_floor[v];

  // Softness (s) of the softplus: s > 0. Smaller s = tighter clamp near the floor.
  real<lower=0> mu_soft;
}
```

```
// ---------------------------------------------------------------------------
// TRANSFORMED DATA
// ---------------------------------------------------------------------------
transformed data {
  // Stacked representation across responses to reuse QR and vectorize the likelihood
  matrix[v * N, v * (p - 1)] XM;      // fixed effects (QR computed here), intercept removed
  matrix[v * N, v * k]       ZM;      // random effects (block-diagonal by response)
  vector[v * N]              Y_stack; // outcomes stacked in same order as XM/ZM rows

  // QR components (thin) and scaling
  matrix[v * N, v * (p - 1)] Q_ast;
  matrix[v * (p - 1), v * (p - 1)] R_ast;
  matrix[v * (p - 1), v * (p - 1)] R_ast_inverse;

  // Build stacked block-diagonal design across responses
  {
    XM = rep_matrix(0, v * N, v * (p - 1));
    ZM = rep_matrix(0, v * N, v * k);
    Y_stack = rep_vector(0, v * N);

    for (m in 1:v) {
      for (n in 1:N) {
        int r = (m - 1) * N + n; // stacked row index
        // Fixed-effects block for response m (drop X intercept column)
        XM[r, (m - 1) * (p - 1) + 1 : m * (p - 1)] = (X[n, 2:p]);
        // Random-effects block for response m
        ZM[r, (m - 1) * k + 1 : m * k] = (Z[n, 1:k]);
        // Outcome
        Y_stack[r] = Y[m][n];
      }
    }
  }

  // QR: thin and scaled (on XM)
  Q_ast         = qr_thin_Q(XM) * sqrt(v * N - 1);
  R_ast         = qr_thin_R(XM) / sqrt(v * N - 1);
  R_ast_inverse = inverse(R_ast);
}

// ---------------------------------------------------------------------------
// PARAMETERS
// ---------------------------------------------------------------------------
parameters {
  // Fixed effects
  vector[v]             beta_Intercept;        // one intercept per response (outside QR)
  vector[v * (p - 1)]   theta_q;               // QR coefficients for X (without intercept)

  // Subject-level random effects (non-centered)
  cholesky_factor_corr[v * k] L_s;             // Cholesky factor of subject ranef correlation
  vector<lower=0>[v * k]      stacked_sigma_b_s;  // subject ranef SDs (half-normal prior)
  vector[v * k]               z_s[n_s];        // standardized subject ranefs

  // Per-response Gamma shape (on log scale for stability)
```

```
  vector[v] log_alpha;
}

// ----------------------------------------------------------------------------
// TRANSFORMED PARAMETERS
// ----------------------------------------------------------------------------
transformed parameters {
  vector[v] g_alpha = exp(log_alpha);          // shape parameter on natural scale

  // Subject-level random effects stacked by response
  vector[v * k] b_stack[n_s];
  {
    matrix[v * k, v * k] LsSigma = diag_pre_multiply(stacked_sigma_b_s, L_s);
    for (j in 1:n_s)
      b_stack[j] = LsSigma * z_s[j];
  }
}

// ----------------------------------------------------------------------------
// MODEL
// ----------------------------------------------------------------------------
model {
  // PRIORS -------------------------------------------------------------
  // Intercepts: robust, weakly-informative. Heavy tails avoid over-shrinkage.
  beta_Intercept ~ student_t(3, 0, 25);

  // Fixed effects on QR-thin X: approx unit-slope prior on original scale.
  theta_q ~ normal(0, 1.5);

  // Subject-level ranef SDs: split intercept vs. other Z columns.
  // Assumption: baselines vary more; slopes are tighter
  for (m in 1:v) {
    int base = (m - 1) * k;
    // intercept term
    stacked_sigma_b_s[base + 1] ~ normal(0, 4.5);
    // other Z columns (if any)
    if (k > 1)
      stacked_sigma_b_s[base + 2 : base + k] ~ normal(0, 1.8);
  }

  // Correlation over subject ranefs: LKJ(1) is uniform on correlation matrices
  L_s ~ lkj_corr_cholesky(1.0);

  // Non-centered base for subject ranefs
  for (j in 1:n_s) z_s[j] ~ normal(0, 1);

  // Gamma shape prior (shared across rows per response)
  // Prior expresses CV around ~9% at alpha = 120
  log_alpha ~ normal(log(120), 0.35);

  // LIKELIHOOD ---------------------------------------------------------
  {
    vector[v * N] mu;         // linear predictor on outcome scale
    vector[v * N] mu_pos;     // clamped positive means
```

```
    vector[v * N] floor_rep;  // per-row replicated floors
    vector[v * N] alpha_rep;  // per-row replicated shape
    vector[v * N] g_beta;     // Gamma rate parameters

    // Fixed effects (QR part)
    mu = Q_ast * theta_q;

    // Add per-response intercepts and subject random effects
    {
      int r = 1;
      for (m in 1:v) {
        for (n in 1:N) {
          mu[r] += beta_Intercept[m] + ZM[r] * b_stack[ subject[n] ];
          alpha_rep[r] = g_alpha[m];
          floor_rep[r] = mu_floor[m];
          r += 1;
        }
      }
    }

    // Softplus clamp to ensure   > 0 (identity link with positivity)
    for (r in 1:(v * N))
      mu_pos[r] = softplus_scaled(mu[r], mu_soft, floor_rep[r]);

    // -Shaperate parameterization
    g_beta = alpha_rep ./ mu_pos;

    // Vectorized Gamma likelihood over stacked rows
    target += gamma_lpdf(Y_stack | alpha_rep, g_beta);
  }
}

// ------------------------------------------------------------------------------
// GENERATED QUANTITIES
// ------------------------------------------------------------------------------
generated quantities {
  // Fixed effects on original X scale (excluding intercept)
  vector[v * (p - 1)] beta_stack = R_ast_inverse * theta_q;
  vector[p - 1] beta[v];               // per-response fixed effects (no intercept)

  // Subject ranef correlation matrix and SDs (for inspection)
  corr_matrix[v * k] C_s = multiply_lower_tri_self_transpose(L_s);
  vector[k] sigma_b_s[v];

  // Posterior predictive draws (no holdout)
  vector[N] Y_rep[v];

  // Convenience unstacked outputs for ranefs
  vector[k] b[n_s, v];                 // subject ranefs per response
  vector[p - 1] personal_effect[n_s, v]; // subject slopes (excl intercept)
  vector[k] subject_effects;           // scratch
  real personal_intercept[n_s, v];

  // Unstack fixed effects and SDs
```

```
  for (m in 1:v) {
    beta[m] = beta_stack[(m - 1) * (p - 1) + 1 : m * (p - 1)];
    sigma_b_s[m] = stacked_sigma_b_s[(m - 1) * k + 1 : m * k];
  }

  // Posterior predictive (all rows)
  {
    vector[v * N] mu;
    vector[v * N] mu_pos;
    vector[v * N] floor_rep;
    vector[v * N] alpha_rep;
    vector[v * N] g_rate;
    vector[v * N] Y_rep_stack;

    mu = Q_ast * theta_q;

    {
      int r = 1;
      for (m in 1:v) {
        for (n in 1:N) {
          mu[r] += beta_Intercept[m] + ZM[r] * b_stack[ subject[n] ];
          alpha_rep[r] = exp(log_alpha[m]);
          floor_rep[r] = mu_floor[m];
          r += 1;
        }
      }
    }

    for (r in 1:(v * N))
      mu_pos[r] = softplus_scaled(mu[r], mu_soft, floor_rep[r]);

    g_rate = alpha_rep ./ mu_pos;
    for (r in 1:(v * N))
      Y_rep_stack[r] = gamma_rng(alpha_rep[r], g_rate[r]);

    // Unstack by response
    for (m in 1:v)
      for (n in 1:N)
        Y_rep[m][n] = Y_rep_stack[(m - 1) * N + n];
  }

  // Subject-level ranefs per response and derived intercept/effects
  for (m in 1:v) {
    for (j in 1:n_s) {
      b[j, m] = b_stack[j, (m - 1) * k + 1 : m * k];
      subject_effects = b[j, m];
      personal_intercept[j, m] = beta_Intercept[m] + subject_effects[1];
      if (k > 1)  // assumes p - 1 == k - 1 for the next line
        personal_effect[j, m] = beta[m] + subject_effects[2:k];
    }
  }
}
```

Listing S2: Stan implementation of the personal nutrient effect model for Dialysis data with two-level hierarchical multivariate regression.

```
// ------------------------------------------------------------------------------------
// Effects of Nutrition - Nested Two-level Hierarchical Gamma GLMM (with identity link)
// ------------------------------------------------------------------------------------
// * Likelihood: y_n ~ Gamma(alpha[m], beta_n) with rate beta_n = alpha[m]/mu_n
// * Link: Identity, with E[y_n] = mu_n on the outcome scale.
// * Positivity enforced via smooth softplus around response-specific floors
// * Random effects at: group (g) and subject (s), same Z structure,
//   with full cross-response correlation (Cholesky factors).
// * QR reparameterization for fixed effects
// ------------------------------------------------------------------------------------

functions {
    // Softplus with scale s > 0 and per-response floor (>0)
    // keeps the implied Gamma mean strictly positive while preserving
    // identity-link behavior away from the floor.

    real softplus_scaled(real x, real s, real floor) {
        return s * log1p_exp((x - floor) / s) + floor;
    }
}

// ------------------------------------------------------------------------------
// DATA
// ------------------------------------------------------------------------------
data {
    int<lower=0> N; // number of observations
    int<lower=1> p; // number of fixed-effect predictors (incl intercept in X[,1])
    int<lower=1> v; // number of responses
    int<lower=1> n_g; // number of groups in data
    int<lower=1> n_s; // number of subjects in data
    int<lower=1> k; // number of personal predictors (incl intercept in Z[,1])


    int<lower=1,upper=n_g> group[N]; // group index per row
    int<lower=1,upper=n_s> subject[N]; // subject index per row
    int<lower=1,upper=n_g> group_for_subject[n_s]; // subject-group mapping


    matrix[N, p] X; // fixed-effect design (X[,1] is intercept; dropped in QR)
    matrix[N, k] Z; // random-effect design (Z[,1] is intercept for g/s levels)


    // Array-of-vectors: one vector of length N per response
    vector[N] Y[v]; // responses (Gamma support: > 0; no transform)


    // Per-response floors for positivity of mu (Gamma rate needs mu > 0)
    real<lower=0> mu_floor[v];


    // Softness of the softplus clamp (s > 0)
    real<lower=0> mu_soft;
```

```
    }

// ------------------------------------------------------------------------------
// TRANSFORMED DATA
// ------------------------------------------------------------------------------
transformed data {
    // Stacked response representation  to reuse QR and vectorize the likelihood
    matrix[v * N, v * (p - 1)] XM; // fixed effects (QR computed here)
    matrix[v * N, v * k] ZM; // random effects (block-diagonal by response)
    vector[v * N] Y_stack; // outcomes stacked in same order as XM/ZM rows

    // QR components (thin) and scaling
    matrix[v * N, v * (p - 1)] Q_ast;
    matrix[v * (p - 1), v * (p - 1)] R_ast;
    matrix[v * (p - 1), v * (p - 1)] R_ast_inverse;

    // Build stacked block-diagonal design across responses
    {
        XM = rep_matrix(0, v * N, v * (p - 1));
        ZM = rep_matrix(0, v * N, v * k);
        Y_stack = rep_vector(0, v * N);

        for (m in 1:v) {
            for (n in 1:N) {
                int r = (m - 1) * N + n; // stacked row index
                // Fixed-effects block for response m (drop X intercept column)
                XM[r, (m - 1) * (p - 1) + 1 : m * (p - 1)] = X[n, 2:p]';
                // Random-effects block for response m
                ZM[r, (m - 1) * k + 1 : m * k] = Z[n, 1:k]';
                // Outcome
                Y_stack[r] = Y[m][n];
            }
        }
    }

    // QR: thin and scaled (on XM)
    Q_ast = qr_thin_Q(XM) * sqrt(v * N - 1);
    R_ast = qr_thin_R(XM) / sqrt(v * N - 1);
    R_ast_inverse = inverse(R_ast);
}

// ------------------------------------------------------------------------------
// PARAMETERS
// ------------------------------------------------------------------------------
parameters {
  // Fixed effects:
  vector[v]            beta_Intercept;       // one intercept per response (outside QR)
  vector[v * (p - 1)]  theta_q;              // QR coefficients for X (no intercept)

  // Group-level random effects:
  cholesky_factor_corr[v * k] L_g;           // Cholesky factor of group ranef correlation
  vector<lower=0>[v * k]       stacked_sigma_b_g;  // group ranef SDs
  vector[v * k]                z_g[n_g];           // standard-normal base variables
```

```
  // Subject-level random effects:
  cholesky_factor_corr[v * k] L_s;           // Cholesky factor of subject ranef correlation
  vector<lower=0>[v * k]       stacked_sigma_b_s;  // subject ranef SDs
  vector[v * k]                z_s[n_s];           // standard-normal base variables

  // Gamma shape (alpha) per response, on log scale to enforce positivity
  vector[v] log_alpha;
}

// --------------------------------------------------------------------------------
// TRANSFORMED PARAMETERS
// --------------------------------------------------------------------------------
transformed parameters {
  vector[v]   g_alpha;                 // shape   for each response (positive)
  vector[v*k] b_stack[n_s];            // subject effects stacked across responses
  vector[v*k] g_stack[n_g];            // group effects   stacked across responses

  {
    matrix[v * k, v * k] Lambda_g = diag_pre_multiply(stacked_sigma_b_g, L_g);
    matrix[v * k, v * k] Lambda_s = diag_pre_multiply(stacked_sigma_b_s, L_s);

    for (j in 1:n_s)
      b_stack[j] = Lambda_s * z_s[j];

    for (j in 1:n_g)
      g_stack[j] = Lambda_g * z_g[j];
  }

  g_alpha = exp(log_alpha);
}

// --------------------------------------------------------------------------------
// MODEL
// --------------------------------------------------------------------------------
model {
    // PRIORS --------------------------------------------------------------
    beta_Intercept ~ student_t(3, 0, 25);

    // Fixed effects on QR-thin X: approx unit-slope prior on original scale.
    theta_q ~ normal(0, 2);

    // Subject-level ranef SDs
    stacked_sigma_b_g ~ normal(0, 3);
    stacked_sigma_b_s ~ normal(0, 3);

    // Correlation over subject ranefs: LKJ(1) is uniform on correlation matrices
    L_g ~ lkj_corr_cholesky(1);
    L_s ~ lkj_corr_cholesky(1);

    // Non-centered base for subject ranefs
    for (j in 1:n_g) z_g[j] ~ normal(0, 1);
    for (j in 1:n_s) z_s[j] ~ normal(0, 1);
```

```
    // Gamma shape prior (shared across rows per response)
    // Prior expresses CV around ~9% at alpha = 120
    log_alpha ~ normal(log(120), 0.35);

    // LIKELIHOOD -------------------------------------------------------------
    {
        vector[v * (N - NH)] mu;        // linear predictor (unclamped)
        vector[v * (N - NH)] mu_pos;    // clamped to positive via softplus
        vector[v * (N - NH)] g_beta;    // Gamma rate = alpha / mu_pos
        vector[v * (N - NH)] alpha_rep; //   replicated per row
        vector[v * (N - NH)] floor_rep; // floors replicated per row
        int vn = 1;

        // Fixed effects (QR part); no offset term
        mu = Q_ast * theta_q;

        // Add per-response intercepts and random effects
        vn = 1;
        for (m in 1:v) {
          for (n in 1:(N - NH)) {
            int nn = idx_tr[n];
            mu[vn] += beta_Intercept[m]
                      + ZM_t[vn] * g_stack[group[nn]]
                      + ZM_t[vn] * b_stack[subject[nn]];
            alpha_rep[vn] = g_alpha[m];
            floor_rep[vn] = mu_floor[m];
            vn += 1;
          }
        }

        // Softplus clamp to ensure   > 0 (identity link with positivity)
        mu_pos = mu_soft * log1p_exp((mu - floor_rep) / mu_soft) + floor_rep;

        // Rate parameter (-shaperate parameterization)
        g_beta = alpha_rep ./ mu_pos;

        // Vectorized Gamma likelihood over stacked training rows
        target += gamma_lpdf(Y_t | alpha_rep, g_beta);
        }
}

// ---------------------------------------------------------------------------
// GENERATED QUANTITIES
// ---------------------------------------------------------------------------
generated quantities {
    // fixed effects on original X scale (excl intercept)
    vector[v * (p - 1)] beta_stack = R_ast_inverse * theta_q;
    corr_matrix[v * k] C_g = multiply_lower_tri_self_transpose(L_g);
    corr_matrix[v * k] C_s = multiply_lower_tri_self_transpose(L_s);

    // Posterior predictive draws for all rows (per response)
    vector[N] Y_rep[v];

    // Convenience unstacked outputs
```

```
vector[p - 1] beta[v]; // per-response fixed effects (no intercept)
vector[k] g[n_g, v]; // group ranefs per response
vector[k] b[n_s, v]; // subject ranefs per response

vector[k] group_effects; // unstacked scratch
vector[k - 1] group_effect[n_g, v];
real group_intercept[n_g, v];

vector[k - 1] personal_effect[n_s, v];
real personal_intercept[n_s, v];

vector[k] sigma_b_g[v]; // ranef SDs unstacked by response
vector[k] sigma_b_s[v];

// Recover per-response fixed effects (original X scale) and SDs
for (m in 1:v) {
    beta[m] = beta_stack[(m - 1) * (p - 1) + 1 : m * (p - 1)];
    sigma_b_g[m] = stacked_sigma_b_g[(m - 1) * k + 1 : m * k];
    sigma_b_s[m] = stacked_sigma_b_s[(m - 1) * k + 1 : m * k];
}

// Posterior predictive
{
    vector[v * N] mu;
    vector[v * N] mu_pos;
    vector[v * N] floor_rep;
    vector[v * N] alpha_rep;
    vector[v * N] g_rate;
    vector[v * N] Y_rep_stack;

    mu = Q_ast * theta_q;

    {
        int r = 1;
        for (m in 1:v) {
            for (n in 1:N) {
                mu[r] += beta_Intercept[m]
                + ZM[r] * g_stack[group[n]]
                + ZM[r] * b_stack[subject[n]];
                alpha_rep[r] = exp(log_alpha[m]);
                floor_rep[r] = mu_floor[m];
                r += 1;
            }
        }
    }

    for (r in 1:(v * N))
        mu_pos[r] = softplus_scaled(mu[r], mu_soft, floor_rep[r]);

    g_rate = alpha_rep ./ mu_pos;
    for (r in 1:(v * N))
        Y_rep_stack[r] = gamma_rng(alpha_rep[r], g_rate[r]);

    // Unstack to per-response arrays
```

```
        for (m in 1:v)
            for (n in 1:N)
                Y_rep[m][n] = Y_rep_stack[(m - 1) * N + n];
    }

    // Unstack ranefs per response and build intercept/effect partitions
    for (m in 1:v) {
        for (i in 1:n_g) {
            g[i, m] = g_stack[i, (m - 1) * k + 1 : m * k];
            group_intercept[i, m] = beta_Intercept[m] + g[i, m][1];
            group_effect[i, m] = beta[m] + g[i, m][2:k];
        }
        for (j in 1:n_s) {
            b[j, m] = b_stack[j, (m - 1) * k + 1 : m * k];
            group_effects = g[group_for_subject[j], m];
            personal_intercept[j, m] = beta_Intercept[m] + group_effects[1] + b[j, m][1];
            if (k > 1)
                personal_effect[j, m] = beta[m] + group_effects[2:k] + b[j, m][2:k];
            else
                personal_effect[j, m][1] = 0; // degenerate if only intercept present
        }
    }
}
```

Listing S3: Stan implementation of the diet inference model with a mixture of concentration control and personal preference of nutrient intake.

```
functions {
  // Function calculates the error in dietary preference by minimizing
  // the weighted differences between the proposed and a given reference diet
  // The function emphasizes changes in nutrients with stronger effects,
  // aiming for minimal overall diet change

  real preference_error(row_vector Q, row_vector reference_Q, row_vector[] beta,
  int nutrients, int responses, real penalty) {

    real error_sum = 0;
    row_vector[nutrients] Q_diffs;

    for (m in 1:responses) {
      for (i in 1:nutrients) {

        // Calculate the difference between the proposed Q and the given reference level
        if (size(reference_Q) == size(Q)) {
          Q_diffs[i] = reference_Q[i] != 0 ? fabs(Q[i] - reference_Q[i])
          ./ fabs(reference_Q[i]) : fabs(Q[i] - reference_Q[i]);
        } else {
          // In calculating maximum error, we compare to both lower and upper limits.
          real max_diff = fmax(fabs(reference_Q[i]-Q[i]), fabs(reference_Q[nutrients+i]-Q[i]));
          Q_diffs[i] = Q[i] != 0 ? max_diff / fabs(Q[i]) : max_diff;
        }
      }
```

```
      // Weights the differences using the inverse of nutrient effects to prioritize changes
      // in nutrients with more significant impacts

      row_vector[nutrients] inverse_beta = 1 ./ (1+fabs(beta[m]));
      real weighted_diffs = dot_product(inverse_beta, Q_diffs);

      // Penalize bigger differences with power function
      error_sum += pow(weighted_diffs / nutrients, penalty);
    }

    // Normalize the sum of penalized preference errors by the number of responses
    return error_sum / responses;
  }
}

data {
    int<lower=1> responses;              // Total number of responses (concentrations)
    int<lower=1> p;                      // Number of predictors
    int<lower=0> r;                      // Number of nutrients considered
    row_vector[r] proposal_lowerlimits;  // Lower intake limits for each nutrient
    row_vector[r] proposal_upperlimits;  // Upper intake limits for each nutrient
    row_vector[r] general_RI;            // General intake recommendations
    row_vector[r] current_Q;             // Current nutrient intake levels
    real Y_lower_limits[responses];      // Lower concentration limits
    real Y_upper_limits[responses];      // Upper concentration limits

    // Personal nutritional effects
    real intercept_point[responses];       // Baseline intercepts from parameter distributions
    row_vector[p] X_beta_point[responses]; // Personal effects for fixed-level nutrients
    row_vector[r] Q_beta_point[responses]; // Personal effects for variable nutrient levels
    row_vector[p] X_evidence_point;

    // Inference hyperparameters
    real limit_steepness;                // l1: Steepness parameter for individual soft limits
    real limit_requirement;              // l2: Requirement for meeting all limits
    real preference_strength;            // l3: Weighting for preference in the model
    real transition_steepness;           // l4: Steepness for the transition between components
    real penalty_rate;                   // l5: Rate at which preference errors are penalized
    real general_recommendation_prior;   // l6: Weight of general recommendation
}

transformed data {

  // Calculate personal maximum preference error by combining the current diet to
  // lower and upper intake limits
  real max_personal_preference_error = preference_error(current_Q, append_col(proposal_lowerlimits,
  proposal_upperlimits), Q_beta_point, r, responses, penalty_rate);

  real lambda_k_min = -lambert_w0(-machine_precision()*max_personal_preference_error) /
  max_personal_preference_error;

  // Scaling with max_personal_preference_error make preference_strength ignorant of
  // nutrient and constraint number
  real personal_preference_strength = lambda_k_min + preference_strength /
```

```
  max_personal_preference_error;

  // Calculates the expected concentration value excluding queried predictors
  real mu_q0[responses];

  for (m in 1:responses) {
    mu_q0[m] = intercept_point[m] + dot_product(X_evidence_point, X_beta_point[m]);
  }
}

parameters {
  // Resulting personalized nutrient intake recommendations
  row_vector<lower=proposal_lowerlimits, upper=proposal_upperlimits>[r] Q;
}
transformed parameters {

  real Y_mu[responses];             // Expected concentration levels for proposed diets
  real Y_mu_Q0[responses] = mu_q0;  // Concentration baselines
  real Y_limit[responses*2];        // Sigmoid values indicating when concentration limits are met
  real softlimit_sum = 0;           // Sum of sigmoid values, with a maximum of 2*responses

  real within_limits_lpdf;          // Log probability of meeting all concentration limits
  real preference_lpdf;             // Log probability of adherence to personal dietary preferences
  real preference_error_sum = 0;    // Sum of dietary preference errors
  real in_range;                    // Indicates whether all concentration limits have been met

  row_vector[r] Q_contributions[responses];  // Normalized contributions

  // Computes expected values and adherence to concentration limits for each response
  for (m in 1:responses) {
    // Expected concentrations for diet proposal Q
    Q_contributions[m] = Q_beta_point[m] .* Q;
    Y_mu[m] = mu_q0[m] + sum(Q_contributions[m]);

    // Sigmoid values for the expected concentrations relative to the limits
    Y_limit[2*m-1] = inv_logit((Y_mu[m] - Y_lower_limits[m]) * limit_steepness);
    Y_limit[2*m] = inv_logit((Y_upper_limits[m] - Y_mu[m]) * limit_steepness);
    softlimit_sum += Y_limit[2*m-1] + Y_limit[2*m];
  }

  // Log probabilities of mixture distribution components
  within_limits_lpdf = normal_lpdf(softlimit_sum | 2*responses, 1/limit_requirement);

  preference_error_sum = preference_error(Q, current_Q, Q_beta_point, r, responses, penalty_rate);
  preference_lpdf = exponential_lpdf(preference_error_sum | personal_preference_strength);

  // Sigmoid coefficient for transitioning between model components
  // - subtracting the constant 0.1 adjusts the sigmoid
  // to reach maximum when all limits are met
  in_range = inv_logit((softlimit_sum - (2*responses - 0.1)) * transition_steepness);
}

model {
```

```
  // PRIORS: The inferred nutrients are given healthy prior limits from the general recommendations,
  // with a mixture distribution adjusting the preference of the generally recommended intake

  for (i in 1:r) {
    real pref_sigma = (proposal_upperlimits[i] - proposal_lowerlimits[i]) /
    general_recommendation_prior;
    target += log_mix(0.5,
                    uniform_lpdf(Q[i] | proposal_lowerlimits[i], proposal_upperlimits[i]),
                    normal_lpdf(Q[i] | general_RI[i], pref_sigma));
  }

  // LIKEHOOD: Mixture model that first aims to reach the concentration limits and
  // then follows the diet preference. Mixture components are conditionally weighted
  // with in_range-parameter (denoted with w in the manuscript).

  target += log_sum_exp(preference_lpdf + log(in_range),
                        within_limits_lpdf + log1m(in_range));

}}
```

### Section "4 Implementation"

#### Extracting the Personal Graphical Models

In this work, graphical models serve not only as a theoretical framework but also as a practical tool for storing personal statistics related to estimated nutritional behavior. By separating the estimation of personal models from downstream inference tasks, it becomes possible to conduct efficient sensitivity analyses of dietary recommendations. This separation allows for systematic testing of different diet inference hyperparameters and concentration limits without repeatedly performing the computationally intensive estimation of the full hierarchical model.

Once the general and individual-specific effects were estimated, the Stan model produced distributions of personal effects for each individual. These personal effects, together with related statistics such as variance, were extracted and stored in GraphML files. GraphML is an XML-based file format designed to represent nodes and edges of graph structures (1).

The following is an example of a node from a GraphML file representing the random variable of an individual's personal effect of vitamin D on plasma albumin:

```
<node id="n155">
  <data key="v_name">personal_dvit_palb</data>
  <data key="v_type">personal</data>
  <data key="v_distribution">dvit_palb_beta.rds</data>
  <data key="v_value">1.47796</data>
  <data key="v_value_lCI">-0.03843</data>
  <data key="v_value_uCI">2.96629</data>
</node>
```

In this example, the node captures the mean of the personal effect along with its lower and upper credible intervals and provides a link to the file containing the full distribution. This structure enables the retrieval of specific quantiles or the use of the entire distribution for further analyses. Furthermore, the extracted personal graph includes the current estimates of nutrient intake levels and other properties produced when the hierarchical model is marginalized into a personal model $\hat{G}_k$.

As the personal models are stored in valid GraphML format, they can be analyzed not only with the *iGraph* package but also with any software supporting graph analytics. This enables the use of the full theory of graphical models in downstream analyses and makes it possible to integrate personal models into broader applications beyond the specific workflow described here.

### Section "5.1 Model Evaluation and Posterior Analysis"

#### Estimation and Sampler Health

##### Sysdimet study

The model implementation is defined in Supplemental Listing S1. This one-level version of the hierarchical Gamma GLMM estimates the effects of nutrients on biochemical responses without the nested subject–group structure used in the Dialysis case. Here, data consisted of 106 subjects with 4 observations from each, resulting 424 rows.

**Estimation settings.** The model was estimated in Stan using the No-U-Turn Sampler (NUTS). Four parallel chains were run with 4000 iterations per chain (1200 warmup, 2800 post-warmup draws), initial radius `init_r`=0.2, `adapt_delta`=0.985, `max_treedepth`=12, and step-size jitter of 0.05. These settings balance stable posterior exploration with practical runtime, using a slightly lower treedepth cap than in the Dialysis model due to the simpler hierarchy.

**Sampler health.** Convergence and stability diagnostics were all within recommended thresholds:

- **Divergences:** None of 11,200 post-warmup draws (0%).
- **Treedepth:** Maximum observed treedepth was 11 (cap 12), with only 0.01% of transitions at this depth, indicating no truncation.
- **E-BFMI:** Chain-wise values ranged 0.86–0.97, all above the 0.3 threshold; the lowest chain (0.86) showed no adverse mixing in trace plots.
- **Convergence:** $\hat{R}_{\max} = 1.008$ (mean $\hat{R} = 1.000$).
- **Effective sample sizes:** Bulk ESS $\geq 839$, tail ESS $\geq 1482$, exceeding conservative thresholds for stable estimation of means and intervals.

Overall, diagnostics indicate well-mixed chains and stable sampling behavior with no evidence of pathological geometry.

**Family-wise diagnostics.** Table S1 reports maximum $\hat{R}$ and minimum effective sample sizes by parameter family. The counted parameters reflect the number of structural parameters monitored per family for which $\hat{R}$ and ESS were computed; latent subject-specific effects are not included in these tallies. We report trace plots in Fig. S1 for parameters with worst (largest) $\hat{R}$ per parameter family. Even with the largest $\hat{R}$ trace plots confirmed good mixing and no chain separation, consistent with global model convergence.

Table S1: Family-wise sampler diagnostics for the Sysdimet case.

| Family | $n_{\text{params}}$ | $\hat{R}_{\text{max}}$ | $\text{ESS}_{\text{bulk,min}}$ | $\text{ESS}_{\text{tail,min}}$ |
|---|---|---|---|---|
| Intercepts | 5 | 1.001 | 2727 | 4683 |
| Fixed effects | 85 | 1.001 | 2741 | 4870 |
| Subject SDs ($\sigma_s$) | 90 | 1.008 | 839 | 1482 |
| Subject correlations ($L_s$) | 4005 | 1.003 | 2714 | 4657 |
| Gamma shapes ($\alpha$) | 5 | 1.001 | 3643 | 6290 |

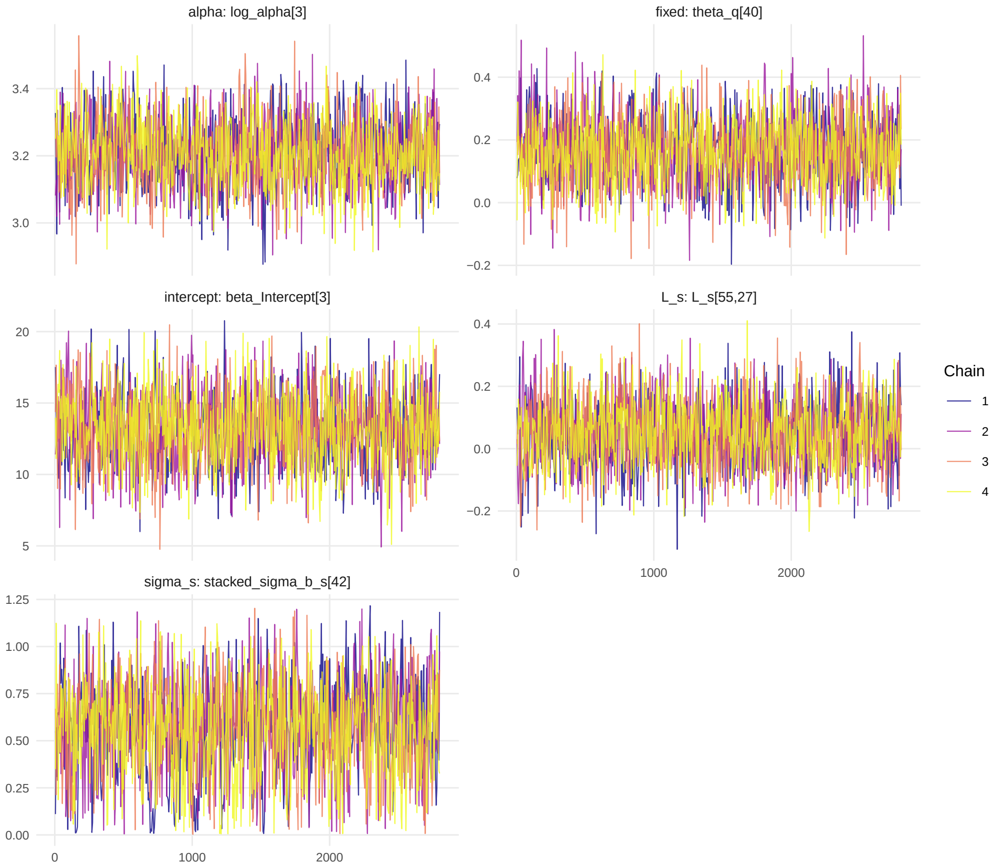


Figure S1: Trace plots for parameters with largest $\hat{R}$ from each family parameters (intercepts, main fixed effects, off-diagonals for subject correlations).

**Estimation time.** Computation time was recorded during both hierarchical model estimation and personal diet inferences for the Sysdimet case. Table S2 reports the warmup and sampling durations for each of the four parallel Markov chains of the hierarchical model. Sampling was executed on a laptop equipped with an Intel Core Ultra 5 135H processor (14 cores/18 threads, 32 GB RAM). The Stan sampler utilized the available CPU cores to run chains in parallel, while GPU acceleration was not used.

Across chains, the total mean runtime was approximately **34,109 s (9.5 h)**, comprising **11,813 s for warmup** and **22,295 s for sampling** on average. After the hierarchical model had converged, personal recommendations were inferred efficiently, averaging **1.29 s ($\pm$ 0.89 s)** per individual.

In addition, the effects of data size and MCMC chain length on estimation time were examined by re-estimating the Sysdimet model under two modified configurations: (i)

Table S2: Hierarchical model estimation times for the Sysdimet case.

| Chain | Warmup (s) | Sampling (s) |
| --- | --- | --- |
| 1 | 11,152.6 | 14,126.0 |
| 2 | 12,265.3 | 25,401.0 |
| 3 | 12,110.5 | 25,183.1 |
| 4 | 11,724.9 | 24,471.6 |
| **Mean** | **11,813.3** | **22,295.4** |
| **Total** | **34,108.8 s (9.5 h)** | |

using half of the original data (53 of 106 subjects) and (ii) using half the chain length (2 000 iterations instead of 4 000). Reducing the dataset to half decreased the total estimation time only marginally, from 9.5 h to 9.0 h, whereas halving the chain length nearly halved the runtime to approximately 5.0 h. Other key sampler settings, such as step size and target acceptance rate, were kept constant across the experiments.

##### Dialysis study

The model implementation is defined in Supplemental Listing S2. Notably, it uses non-centered parameterization to better capture information from the limited number of observations. Here, data consisted of 37 subjects with 2 observations from each, resulting 72 rows.

**Estimation settings.** The Gamma GLMM for the effects of nutrients with nested hierarchy of subjects in treatment groups was estimated in Stan using the No-U-Turn Sampler (NUTS). Six parallel chains were run with 4000 iterations per chain (1200 warmup, 2800 post-warmup draws), initial radius `init_r`=0.2, `adapt_delta`=0.985, `max_treedepth`=15 (default cap), and no step-size jitter. These settings balance stable posterior geometry exploration against runtime.

**Sampler health.** Convergence and stability diagnostics were all within recommended thresholds:

- **Divergences:** 14 of 16,800 post-warmup draws (0.08%), distributed sparsely across chains.
- **Treedepth:** Maximum observed treedepth was 14 (cap 15), with 16.7% of transitions at this depth, consistent with complex posterior geometry but without truncation.
- **E-BFMI:** Chain-wise values ranged 0.85–0.91, all comfortably above the 0.3 threshold.
- **Convergence:** $\hat{R}_{\max} = 1.003$ (mean $\hat{R} = 1.000$).

- **Effective sample sizes:** Bulk ESS $\geq$ 1678, tail ESS $\geq$ 3128, far above conservative thresholds for stable estimation of means and intervals.

Overall, diagnostics indicate well-mixed chains and stable sampling behavior with no evidence of pathological geometry.

**Family-wise diagnostics.** Table S3 reports maximum $\hat{R}$ and minimum effective sample sizes by parameter family. We report trace plots in Fig. S2 for parameters with worst (largest) $\hat{R}$ per parameter family. Even with the largest $\hat{R}$ trace plots for core parameters confirmed good mixing and no chain separation as indication of global model convergence.

Table S3: Family-wise sampler diagnostics for the Dialysis case.

| Family | $n_{\text{params}}$ | $\hat{R}_{\text{max}}$ | $\text{ESS}_{\text{bulk,min}}$ | $\text{ESS}_{\text{tail,min}}$ |
|---|---|---|---:|---:|
| Intercepts | 3 | 1.002 | 5343 | 8343 |
| Fixed effects | 57 | 1.003 | 3684 | 5535 |
| Group SDs ($\sigma_g$) | 60 | 1.001 | 7013 | 6193 |
| Group correlations ($L_g$) | 1770 | 1.002 | 6214 | 8733 |
| Subject SDs ($\sigma_s$) | 60 | 1.001 | 4996 | 6094 |
| Subject correlations ($L_s$) | 1770 | 1.002 | 6268 | 7908 |
| Gamma shapes ($\alpha$) | 3 | 1.002 | 1678 | 3128 |

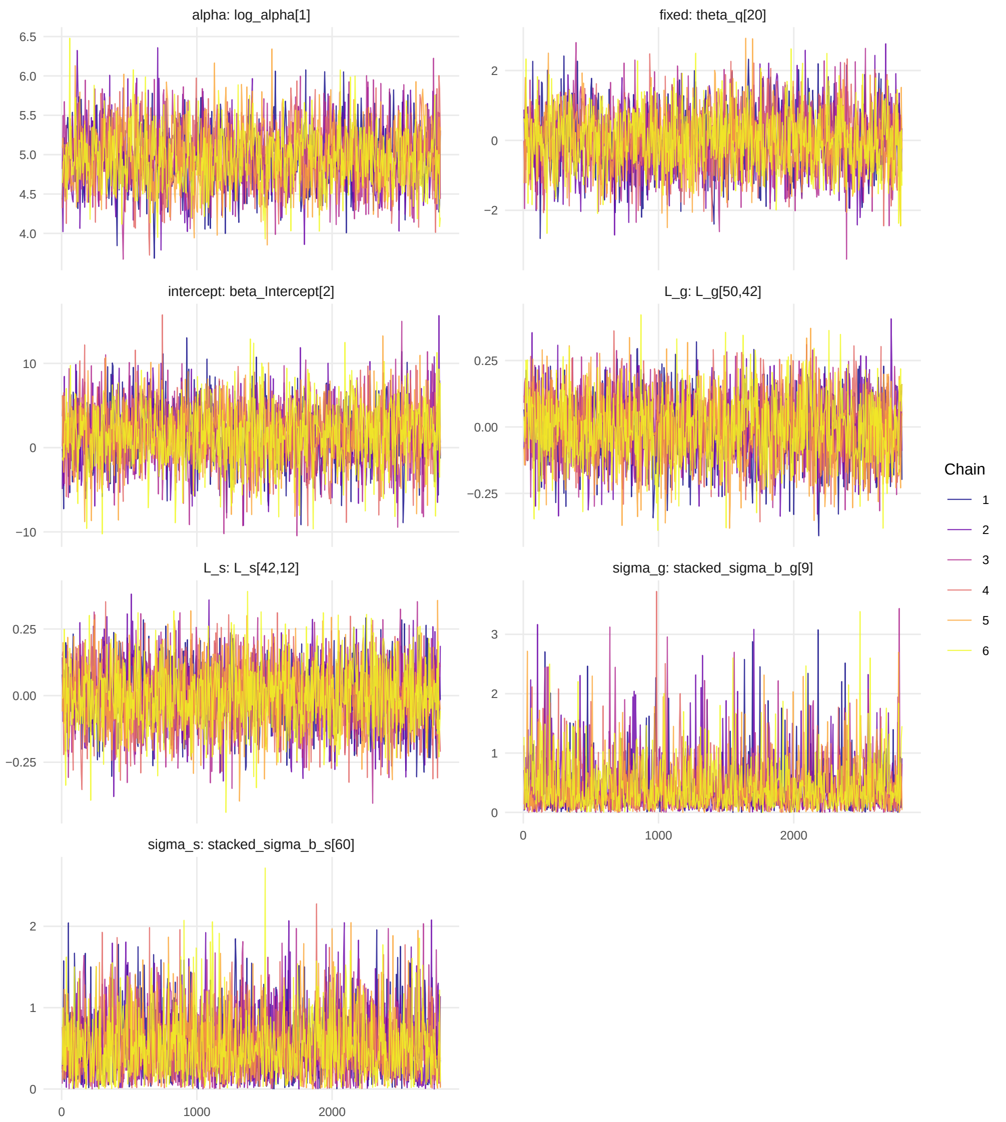


Figure S2: Trace plots for parameters with largest $\hat{R}$ from each family parameters (intercepts, main fixed effects, variance components).

As expected for variance components under half-Normal priors, the posterior distributions for random-effect SDs are skewed toward zero. This reflects limited information for those variance terms rather than convergence issues. R-hat values were 1.0 and ESS >4000 across SDs, confirming stable sampling.

**Estimation time.** Computation time was monitored during both hierarchical model estimation and personal diet inferences for the Dialysis case. Table S4 reports the warmup and sampling durations for each of the six parallel Markov chains of the hierarchical model. Sampling was executed on a laptop equipped with an Intel Core Ultra 5 135H processor (14 cores/18 threads, 32 GB RAM). The Stan sampler utilized the available CPU cores to run chains in parallel, while GPU acceleration was not used.

Across chains, the total mean runtime was approximately **62,775 s (17.4 h)**, comprising **20,837 s for warmup** and **41,938 s for sampling** on average. After the hierarchical model had converged, personal recommendations were inferred efficiently, averaging **1.77 s (± 0.44 s)** per individual.

Table S4: Hierarchical model estimation times for the Dialysis case.

| Chain | Warmup (s) | Sampling (s) |
|---|---|---|
| 1 | 19,877.4 | 32,935.2 |
| 2 | 23,138.5 | 31,800.8 |
| 3 | 20,225.1 | 40,010.1 |
| 4 | 22,262.4 | 66,916.3 |
| 5 | 19,666.8 | 32,973.1 |
| 6 | 19,853.3 | 46,991.9 |
| **Mean** | **20,837.3** | **41,937.9** |
| **Total** | **62,775.2 s (17.4 h)** | |

#### Predictive Accuracy: NRMSE

The normalized root mean squared error (NRMSE) was used to evaluate predictive accuracy:

$$\mathrm{NRMSE}_{\mathrm{Y_m}} = \frac{\sqrt{\frac{1}{n}\sum_{j=1}^{n}\left(\hat{Y}_{i_j} - Y_{i_j}\right)^2}}{\bar{Y}} \tag{1}$$

where $n$ is the number of observations, $\hat{Y}_{i_j}$ the predicted value, $Y_{i_j}$ the observed value, and $\bar{Y}$ the mean of observations.

Table S5: NRMSE values for predicted plasma concentrations.

| **Plasma concentration** | **Sysdimet (NRMSE)** | **Dialysis (NRMSE)** |
|---|---|---|
| HDL cholesterol | 0.05 | - |
| LDL cholesterol | 0.05 | - |
| Insulin | 0.14 | - |
| Total cholesterol | 0.04 | - |
| Glucose | 0.03 | - |
| Potassium | - | 0.15 |
| Phosphate | - | 0.31 |
| Albumin | - | 0.18 |

#### Posterior Predictive Checks

We conducted posterior predictive checks using both visual overlays and Bayesian $p$-values. Visual PPCs are shown in Figs. S3 and S4, while Table S6 reports predictive $p$-values for mean, variance, skewness, and upper-tail exceedance.

Across both datasets, posterior predictive $p$-values were generally near 0.5, indicating good calibration. Only a few responses showed mild tail or skewness mismatches, which are expected given the data sparsity.

Table S6: Posterior predictive $p$-values for the Sysdimet and Dialysis models. Values near 0.5 indicate good calibration; values close to 0 or 1 indicate potential misfit for the chosen discrepancy statistic.

| **Model** | **Response** | $p_{\text{mean}}$ | $p_{\text{sd}}$ | $p_{\text{skew}}$ | $p_{\text{tail 90}}$ |
|---|---|---|---|---|---|
| Sysdimet | fS-HDL | 0.556 | 0.502 | 0.634 | 0.226 |
| Sysdimet | fS-LDL | 0.568 | 0.668 | 0.794 | 0.603 |
| Sysdimet | fS-Chol | 0.539 | 0.306 | 0.653 | 0.174 |
| Sysdimet | fS-Ins | 0.557 | 0.714 | 0.784 | 0.809 |
| Sysdimet | fP-Gluc | 0.543 | 0.785 | 0.021 | 0.460 |
| Dialysis | P-K | 0.513 | 0.031 | 0.725 | 0.233 |
| Dialysis | fP-Pi | 0.422 | 0.171 | 0.821 | 0.105 |
| Dialysis | P-Alb | 0.724 | 0.102 | 0.026 | 0.085 |

Our interpretations of the posterior predictive p-values:

- Sysdimet / fS-HDL: well-calibrated mean and variance; 90th percentile a bit low (p=0.23) suggesting slight underestimation of upper tail.
- Sysdimet / fS-LDL: good calibration across all statistics, with no evidence of misfit.

- Sysdimet / fS-Chol: mean OK, but SD and 90th percentile low (p=0.31, 0.17), indicating underestimation of spread and right tail.
- Sysdimet / fS-Ins: well-calibrated mean and variance; 90th percentile somewhat high (p=0.81), hinting at mild overestimation of extremes.
- Sysdimet / fP-Gluc: mean and variance acceptable, but skewness not captured (p=0.02), consistent with underestimation of asymmetry.
- Dialysis / P-K: reasonably well-calibrated overall; only the SD check is low (p=0.03), suggesting simulated spread > observed.
- Dialysis / fP-Pi: good calibration, with some tendency toward narrower tails (p=0.11).
- Dialysis / P-Alb: mean calibration OK, but SD and 90th percentile are low and skewness is extreme (p=0.026); consistent with PPC observation that albumin may be influenced by other (non-nutritional) factors.

**Visual PPCs**

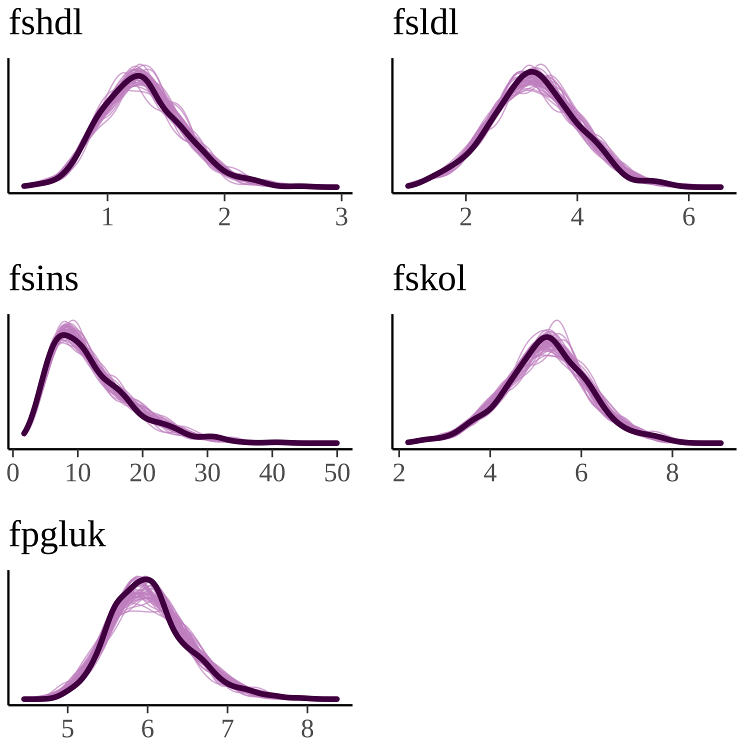


Figure S3: Figure shows a visual posterior predictive checks for the Sysdimet data set, illustrating the fit between observed and simulated plasma concentrations. This model was used to estimate the general and personal effects of nutrients on fasting plasma concentrations of high-density lipoprotein (fshdl), low-density lipoprotein (fsldl), cholesterol (fskol), insulin (fsins), and glucose (fpgluk). The figure was plotted with bayesplot package for R language (v 1.14, https://cran.r-project.org/web/packages/bayesplot/index.html).

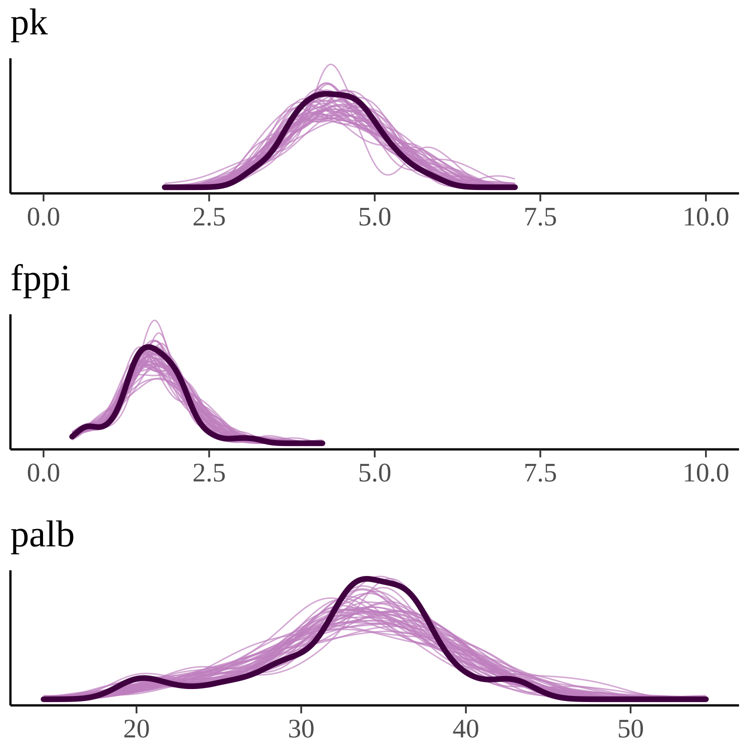


Figure S4: Figure shows a visual posterior predictive checks for the Dialysis data set, illustrating the fit between observed and simulated plasma concentrations. This model was used to estimate the general and personal effects of nutrients on fasting plasma potassium (pk), fasting plasma phosphate (fppi), and plasma albumin (palb). The figure is plotted with bayesplot package for R language (v 1.14, https://cran.r-project.org/web/packages/bayesplot/index.html).

### Section "5.2 Diverse Personal Effects of Nutrients"

Table S7: **Strongest and personally most varying effects of nutrients and medication in the Sysdimet data set.**

| Effect | Avg | SD | Min personal | Max personal |
|---|---|---|---|---|
| Alpha-linolenic acid → Insulin | -7.71 | 0.01 | -7.74 [−13.09; −2.41] | -7.68 [−13.07; −2.36] |
| Linoleic acid → Insulin | 7.58 | 0.01 | 7.56 [2.24; 12.87] | 7.60 [2.32; 12.91] |
| Alpha-linolenic acid → LDL-chol. | -0.84 | 0.00 | -0.84 [−1.49; −0.17] | -0.83 [−1.48; −0.17] |
| Linoleic acid → LDL-chol. | 0.81 | 0.00 | 0.81 [0.16; 1.46] | 0.81 [0.16; 1.46] |
| Alpha-linolenic acid → Total chol. | -0.73 | 0.00 | -0.73 [−1.56; 0.11] | -0.72 [−1.55; 0.11] |
| Linoleic acid → Total chol. | 0.70 | 0.00 | 0.70 [−0.13; 1.51] | 0.70 [−0.13; 1.52] |
| Folate → Insulin | -0.72 | 0.14 | -1.06 [−1.77; −0.48] | -0.31 [−1.00; 0.61] |
| Vitamin C → Insulin | 0.28 | 0.28 | -0.29 [−1.09; 0.37] | 1.03 [0.04; 2.35] |
| Cholesterol → Insulin | 0.50 | 0.09 | 0.32 [−0.29; 0.86] | 0.68 [0.06; 1.41] |
| MUFA → Insulin | 0.44 | 0.23 | -0.09 [−0.96; 0.69] | 0.81 [−0.12; 1.94] |
| Linoleic acid → Glucose | -0.43 | 0.00 | -0.43 [−1.12; 0.26] | -0.43 [−1.12; 0.27] |
| SFA → Insulin | -0.27 | 0.24 | -0.58 [−1.29; 0.10] | 0.25 [−0.63; 1.27] |
| Alpha-linolenic acid → Glucose | 0.41 | 0.00 | 0.41 [−0.29; 1.11] | 0.41 [−0.28; 1.11] |
| Carbohydrates → Insulin | 0.08 | 0.18 | -0.23 [−0.81; 0.30] | 0.49 [−0.17; 1.27] |
| Vitamin D → Insulin | 0.36 | 0.11 | 0.13 [−0.63; 0.79] | 0.55 [−0.13; 1.37] |
| Sucrose → Insulin | 0.23 | 0.04 | 0.16 [−0.24; 0.55] | 0.33 [−0.11; 0.81] |
| PUFA → Insulin | -0.17 | 0.09 | -0.42 [−1.04; 0.12] | -0.04 [−0.62; 0.64] |
| Protein → Insulin | -0.22 | 0.08 | -0.37 [−0.82; 0.03] | -0.05 [−0.49; 0.47] |
| EPA-fatty acid → Insulin | 0.06 | 0.08 | -0.11 [−1.13; 0.94] | 0.30 [−0.76; 1.46] |
| Fiber → Insulin | -0.08 | 0.08 | -0.24 [−0.78; 0.26] | 0.13 [−0.47; 0.86] |

DHA = Docosahexaenoic Acid; EPA = Eicosapentaenoic Acid; MUFA = Monounsaturated fatty acid; LDL-cholesterol = Low-Density Lipoprotein Cholesterol; PUFA = Polyunsaturated fatty acid

The table presents the effects of the adjusted nutrients on the considered concentrations. It shows the average effect ($\hat{\beta}_{jm}$), standard deviation between persons, and minimum and maximum personal effects ($\hat{\beta}_{jm} + \hat{b}_{kjm}$) within the study with 90% credible intervals.

Table S8: **Strongest and personally most varying effects of nutrients in the Dialysis data set.**

| Effect | Avg | SD | Min personal | Max personal |
|---|---|---|---|---|
| Salt → Albumin | 0.97 | 0.89 | -0.62 [−3.56; 1.88] | 2.55 [−0.19; 5.82] |
| Sucrose → Albumin | 1.34 | 0.53 | 0.65 [−2.14; 3.03] | 2.31 [0.28; 4.55] |
| Potassium → Albumin | -0.94 | 0.42 | -1.77 [−5.15; 1.17] | -0.39 [−3.09; 2.41] |
| Vitamin D → Albumin | 0.21 | 0.65 | -0.91 [−4.15; 1.63] | 1.15 [−1.17; 3.77] |
| Water → Albumin | -0.39 | 0.53 | -1.54 [−4.02; 0.67] | 0.20 [−2.12; 2.76] |
| PUFA → Albumin | 0.31 | 0.55 | -0.41 [−2.43; 1.55] | 1.32 [−1.12; 4.04] |
| Calcium → Albumin | -0.81 | 0.23 | -1.20 [−4.09; 1.58] | -0.38 [−3.43; 2.86] |
| MUFA → Albumin | 0.78 | 0.37 | 0.17 [−2.19; 2.34] | 1.39 [−1.00; 4.10] |
| Fiber → Albumin | -0.33 | 0.28 | -1.26 [−4.77; 1.54] | 0.12 [−1.93; 2.18] |
| Salt → Potassium | -0.08 | 0.42 | -0.85 [−1.69; 0.00] | 0.41 [−0.30; 1.18] |
| Phosphorous → Albumin | 0.49 | 0.36 | -0.29 [−4.24; 3.48] | 0.97 [−3.11; 5.22] |
| Carbohydrates → Albumin | 0.21 | 0.31 | -0.44 [−2.50; 1.50] | 0.71 [−1.17; 2.65] |
| SFA → Albumin | 0.04 | 0.14 | -0.46 [−2.97; 1.93] | 0.20 [−1.93; 2.34] |
| SFA → Potassium | 0.27 | 0.07 | 0.20 [−0.40; 0.80] | 0.41 [−0.28; 1.20] |
| Fiber → Potassium | 0.22 | 0.18 | 0.01 [−0.58; 0.58] | 0.54 [−0.36; 1.72] |
| Protein → Albumin | 0.26 | 0.14 | -0.05 [−1.78; 1.54] | 0.45 [−0.94; 1.88] |
| Vitamin D → Potassium | -0.18 | 0.11 | -0.43 [−1.20; 0.21] | 0.05 [−0.24; 0.34] |
| Potassium → Potassium | -0.19 | 0.07 | -0.34 [−1.04; 0.30] | -0.11 [−0.61; 0.42] |
| Phosphorous → Phosphate | 0.16 | 0.01 | 0.14 [−0.31; 0.58] | 0.22 [−0.22; 0.67] |
| Carbohydrates → Potassium | 0.19 | 0.05 | 0.10 [−0.58; 0.77] | 0.25 [−0.46; 0.97] |

MUFA = Monounsaturated fatty acid; PUFA = Polyunsaturated fatty acid; SFA = Saturated fatty acid

The table presents the effects of the adjusted nutrients on the considered concentrations, the average effect ($\hat{\beta}_{jm}$), standard deviation between persons, and minimum and maximum personal effects ($\hat{\beta}_{jm} + \hat{b}_{kjm}$) within the study with 90% credible intervals.

### Section "5.3 Personally Inferred Dietary Recommendations"

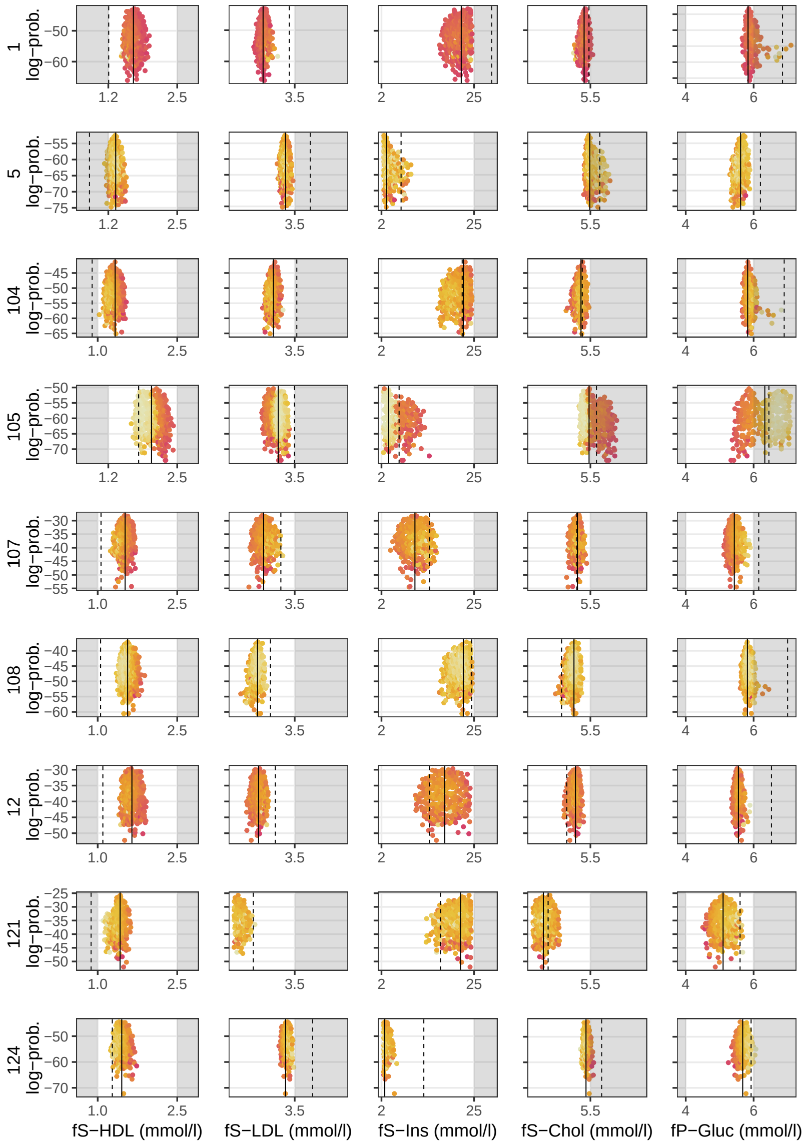


Figure S5: Figure shows the remaining 19 of a total of 24 patients from the control cohort of the Sysdimet study, whose current concentrations deviate from normal ranges but were not reported in the main article. The concentrations of all patients, except number 121, could be improved with personalized diet recommendations. The white areas represent the normal concentration ranges, while the dashed vertical lines mark the patients' current concentrations, some of which fall within the gray areas. The solid lines indicate the concentration levels corresponding to the expected value of the joint multivariate recommendation distribution, where the nutrient intake and concentration predictions are jointly optimized. The figure is plotted with ggplot2 package for R language (v 3.3.5, https://ggplot2.tidyverse.org).

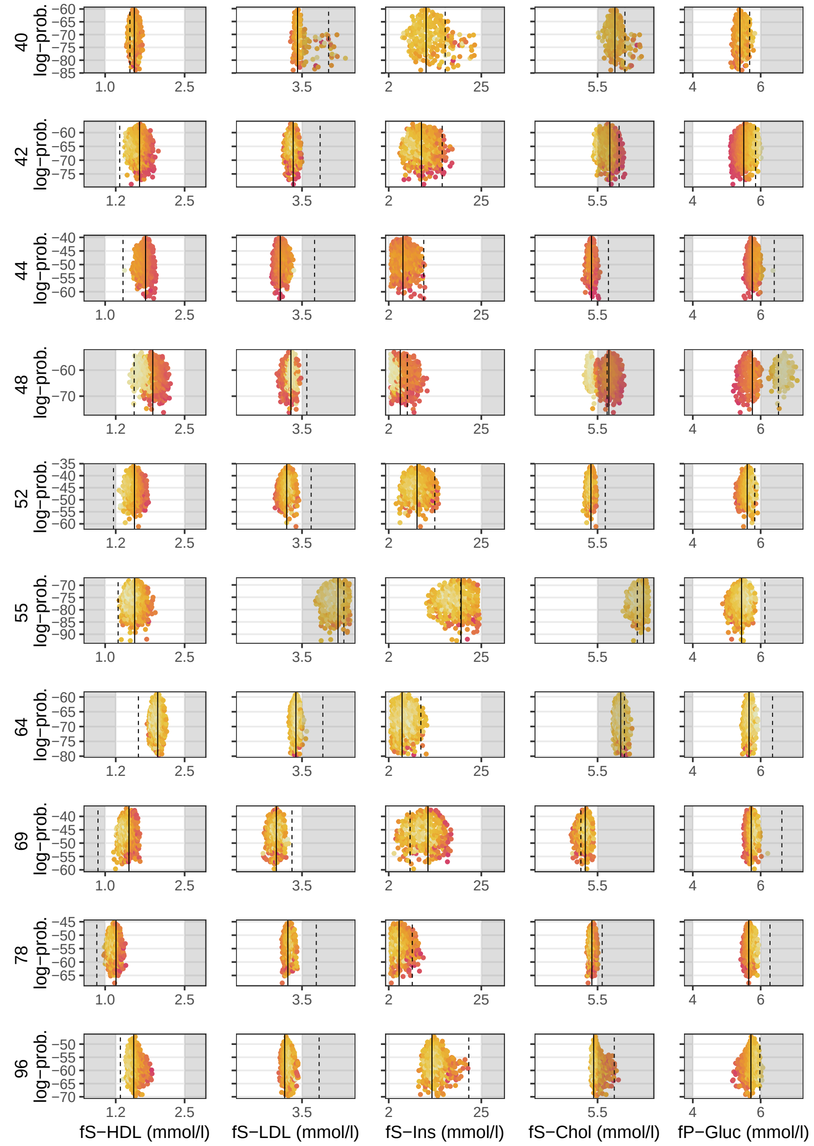

(Continued) Figure shows the remaining 19 of a total of 24 patients from the control cohort of the Sysdimet study, whose current concentrations deviate from normal ranges but were not reported in the main article. The concentrations of all patients, except number 121, could be improved with personalized diet recommendations. The white areas represent the normal concentration ranges, while the dashed vertical lines mark the patients' current concentrations, some of which fall within the gray areas. The solid lines indicate the concentration levels corresponding to the expected value of the joint multivariate recommendation distribution, where the nutrient intake and concentration predictions are jointly optimized. The figure is plotted with ggplot2 package for R language (v 3.3.5, https://ggplot2.tidyverse.org).

Table S9: Personally inferred intake recommendations for all 24 control cohort patients in the Sysdimet study whose current concentrations were out of the normal ranges. The table shows the current intake of nutrients and the recommended levels that are expected to produce normal concentrations and cause the smallest overall change in the current diet. Red and blue colors indicate an increase and decrease to the current levels, respectively, with bold font indicating an over 30% change. Results for individuals whose simulated recommendations could not satisfy all concentration limits are considered non-comparable and shown as all-zero rows.

| | Protein (E%) | Carboh. (E%) | Sucrose (E%) | Fiber (g/d) | SFA (E%) | MUFA (E%) | PUFA (E%) | Linol. acid (E%) | α-linol. acid (E%) | EPA (mg/d) | DHA (mg/d) | Chol. (mg/d) | Folate (μg/d) | Vit. D (μg/d) | Vit. C (mg/d) |
|---|---|---|---|---|---|---|---|---|---|---|---|---|---|---|---|
| 1 | 20.1→ 16.2 [10.7;19.9] | 51.3→ 53.2 [42.3;59.7] | 8.7→ **2.2** [0.1;7.3] | 26.4→ 27.1 [20.3;42.5] | 11.1→ **8.5** [7.1;9.9] | 7.2→ **15.8** [10.7;19.8] | 3.6→ **8.2** [5.4;9.9] | 2.7→ **5.7** [2.6;8.8] | 0.6→ **3.3** [1.0;4.9] | 0.0→ **2.4** [2.2;2.5] | 0.1→ 0.1 [0.0;0.3] | 232→ **48** [1;161] | 373→ **694** [326;973] | 6.1→ **34.0** [8.9;74.7] | 199→ **95** [75;142] |
| 5 | 17.1→ **0.0** [0.0;0.0] | 53.9→ **0.0** [0.0;0.0] | 7.7→ **0.0** [0.0;0.0] | 21.0→ **0.0** [0.0;0.0] | 9.1→ **0.0** [0.0;0.0] | 10.0→ **0.0** [0.0;0.0] | 4.8→ **0.0** [0.0;0.0] | 3.7→ **0.0** [0.0;0.0] | 0.9→ **0.0** [0.0;0.0] | 0.0→ 0.0 [0.0;0.0] | 0.0→ **0.0** [0.0;0.0] | 163→ **0** [0;0] | 226→ **0** [0;0] | 4.8→ **0.0** [0.0;0.0] | 86→ **0** [0;0] |
| 104 | 17.8→ **13.5** [10.2;19.2] | 48.7→ 48.5 [40.4;59.0] | 4.4→ **2.8** [0.1;8.3] | 24.0→ **35.0** [20.8;53.7] | 12.0→ **8.5** [7.1;9.9] | 10.1→ 12.0 [10.1;16.2] | 5.1→ **7.7** [5.2;9.9] | 3.8→ **6.2** [2.8;8.9] | 0.5→ **2.3** [0.6;4.4] | 0.2→ **2.4** [2.2;2.5] | 0.6→ **0.1** [0.0;0.3] | 346→ **62** [2;194] | 282→ 316 [111;617] | 8.3→ **67.1** [25.5;97.1] | 161→ 148 [77;300] |
| 105 | 21.4→ **0.0** [0.0;0.0] | 49.6→ **0.0** [0.0;0.0] | 4.5→ **0.0** [0.0;0.0] | 19.8→ **0.0** [0.0;0.0] | 10.8→ **0.0** [0.0;0.0] | 7.3→ **0.0** [0.0;0.0] | 4.9→ **0.0** [0.0;0.0] | 1.7→ **0.0** [0.0;0.0] | 0.3→ **0.0** [0.0;0.0] | 0.0→ **0.0** [0.0;0.0] | 0.1→ **0.0** [0.0;0.0] | 210→ **0** [0;0] | 311→ **0** [0;0] | 8.3→ **0.0** [0.0;0.0] | 219→ **0** [0;0] |
| 107 | 13.7→ 15.2 [10.3;19.7] | 40.6→ 49.7 [40.5;59.3] | 7.9→ **3.7** [0.1;9.4] | 14.9→ **34.1** [20.6;53.1] | 13.7→ **8.4** [7.1;9.9] | 15.5→ 15.3 [10.4;19.8] | 6.4→ 7.8 [5.2;9.9] | 3.1→ **5.6** [2.7;8.7] | 0.8→ **3.5** [1.2;4.9] | 0.1→ **2.2** [1.7;2.5] | 0.2→ 0.3 [0.0;0.9] | 277→ **122** [6;284] | 239→ **762** [329;992] | 9.9→ **65.2** [17.9;98.5] | 120→ **293** [81;787] |
| 108 | 17.8→ **13.6** [10.1;19.1] | 54.0→ 49.7 [40.5;59.3] | 6.3→ **3.9** [0.2;9.5] | 15.2→ **40.0** [21.7;54.0] | 11.4→ **8.6** [7.1;9.9] | 8.8→ 12.0 [10.1;16.3] | 4.0→ **7.5** [5.2;9.8] | 3.2→ **6.4** [2.8;8.9] | 0.6→ **1.3** [0.5;3.1] | 0.0→ **2.4** [2.2;2.5] | 0.1→ **0.2** [0.0;0.5] | 222→ **80** [3;239] | 203→ 270 [106;618] | 5.6→ **31.8** [8.8;72.4] | 61→ **191** [78;443] |
| 12 | 20.1→ **14.9** [10.3;19.7] | 55.3→ 50.9 [40.7;59.5] | 3.1→ 4.0 [0.2;9.5] | 31.3→ 36.7 [20.9;53.8] | 7.9→ 8.6 [7.1;9.9] | 6.7→ **14.0** [10.2;19.4] | 4.7→ **7.7** [5.2;9.9] | 3.8→ **6.2** [2.8;8.9] | 0.7→ **1.6** [0.5;3.8] | 0.0→ **2.4** [2.1;2.5] | 0.1→ 0.1 [0.0;0.5] | 322→ **106** [4;273] | 286→ 401 [121;846] | 11.5→ **54.2** [11.9;96.6] | 95→ **334** [82;862] |
| 121 | 22.5→ **13.7** [10.2;19.2] | 48.7→ 46.4 [40.2;57.1] | 9.7→ **3.9** [0.2;9.4] | 22.0→ **37.3** [20.9;54.0] | 10.4→ 8.4 [7.1;9.9] | 8.3→ **13.7** [10.2;19.1] | 4.9→ **7.5** [5.1;9.9] | 3.4→ **5.6** [2.7;8.7] | 0.8→ **3.5** [1.2;4.9] | 0.2→ **2.1** [1.3;2.5] | 0.4→ **0.8** [0.0;2.1] | 306→ **149** [10;291] | 271→ **540** [135;958] | 10.0→ **77.3** [34.7;99.2] | 83→ **400** [90;924] |
| 124 | 13.2→ 17.4 [12.1;19.9] | 43.2→ 54.2 [43.2;59.8] | 6.3→ **2.3** [0.1;7.5] | 22.7→ 22.7 [20.1;29.1] | 14.3→ **7.8** [7.0;9.4] | 16.9→ 19.3 [17.5;20.0] | 7.9→ 6.7 [5.1;9.5] | 4.3→ 5.2 [2.6;8.7] | 0.8→ **4.3** [2.9;5.0] | 0.0→ **1.5** [1.1;1.8] | 0.1→ 0.1 [0.0;0.4] | 287→ **80** [2;246] | 205→ **762** [566;963] | 13.9→ **27.0** [8.3;59.7] | 128→ 128 [77;252] |
| 127 | 17.2→ 15.4 [10.5;19.8] | 54.7→ 44.2 [40.2;51.9] | 8.1→ **0.8** [0.0;2.6] | 16.6→ 22.6 [20.1;29.3] | 10.2→ **7.6** [7.0;8.8] | 7.4→ **18.9** [16.1;20.0] | 2.9→ **7.6** [5.2;9.9] | 2.4→ **3.5** [2.5;5.9] | 0.4→ **4.9** [4.5;5.0] | 0.0→ **1.8** [1.5;2.2] | 0.1→ 0.1 [0.0;0.3] | 230→ **48** [1;159] | 149→ **968** [898;998] | 4.8→ **18.8** [7.8;37.0] | 97→ 100 [76;157] |
| 15 | 17.3→ 17.1 [11.6;19.9] | 42.4→ 51.1 [40.9;59.6] | 5.4→ **3.5** [0.1;9.2] | 17.7→ 24.2 [20.1;34.5] | 10.1→ 7.9 [7.0;9.5] | 8.9→ **18.7** [15.6;20.0] | 5.1→ 6.3 [5.0;9.0] | 4.1→ 4.6 [2.6;8.3] | 0.8→ **4.7** [3.9;5.0] | 0.0→ **0.4** [0.1;0.9] | 0.1→ **0.2** [0.0;0.6] | 232→ **91** [3;263] | 252→ **430** [153;773] | 6.7→ **13.3** [7.7;27.0] | 151→ **456** [110;916] |
| 18 | 20.1→ **14.9** [10.3;19.6] | 42.9→ 49.9 [40.7;59.2] | 4.2→ 4.6 [0.3;9.6] | 17.0→ **37.2** [21.0;53.9] | 14.0→ **8.5** [7.1;9.9] | 9.4→ **14.8** [10.3;19.7] | 4.4→ **7.5** [5.1;9.9] | 3.1→ **5.8** [2.7;8.8] | 0.9→ **2.5** [0.6;4.8] | 0.1→ **2.2** [1.6;2.5] | 0.2→ **0.5** [0.0;1.3] | 229→ **146** [8;292] | 203→ **481** [118;947] | 4.1→ **61.7** [14.4;97.9] | 48→ **517** [99;974] |
| 19 | 18.8→ 14.9 [10.3;19.7] | 47.4→ 48.5 [40.3;58.9] | 5.8→ **2.8** [0.1;8.4] | 18.0→ **31.4** [20.4;51.2] | 13.4→ **8.4** [7.1;9.9] | 8.8→ **16.7** [11.2;19.9] | 4.7→ **7.3** [5.1;9.8] | 3.3→ **5.5** [2.6;8.8] | 1.0→ **3.9** [1.9;5.0] | 0.1→ **2.2** [1.7;2.5] | 0.2→ **0.3** [0.0;1.0] | 233→ **131** [6;288] | 233→ **674** [312;969] | 5.9→ **85.0** [52.2;99.5] | 106→ **210** [79;507] |
| 24 | 18.1→ 16.0 [10.6;19.9] | 38.7→ 51.3 [40.9;59.6] | 7.5→ **4.3** [0.2;9.5] | 12.5→ **31.9** [20.5;51.7] | 19.8→ **8.2** [7.0;9.8] | 11.8→ 16.6 [11.0;19.9] | 3.9→ **7.7** [5.2;9.9] | 2.5→ **5.3** [2.6;8.7] | 0.5→ **4.1** [2.3;5.0] | 0.0→ **1.4** [0.6;2.3] | 0.1→ **0.3** [0.0;0.9] | 339→ **100** [3;267] | 247→ **863** [560;996] | 4.8→ **57.6** [13.3;96.5] | 91→ **381** [87;894] |
| 40 | 20.0→ **0.0** [0.0;0.0] | 42.9→ **0.0** [0.0;0.0] | 6.0→ **0.0** [0.0;0.0] | 18.2→ **0.0** [0.0;0.0] | 13.0→ **0.0** [0.0;0.0] | 12.2→ **0.0** [0.0;0.0] | 5.4→ **0.0** [0.0;0.0] | 3.7→ **0.0** [0.0;0.0] | 1.1→ **0.0** [0.0;0.0] | 0.2→ **0.0** [0.0;0.0] | 0.4→ **0.0** [0.0;0.0] | 310→ **0** [0;0] | 182→ **0** [0;0] | 11.9→ **0.0** [0.0;0.0] | 46→ **0** [0;0] |

EPA = Eicosapentaenoic acid, DHA = Docosahexaenoic acid

Table S9: (Continued) Personally inferred intake recommendations for all 24 control cohort patients in the Sysdimet study whose current concentrations were out of the normal ranges. The table shows the current intake of nutrients and the recommended levels that are expected to produce normal concentrations and cause the smallest overall change in the current diet. Red and blue colors indicate an increase and decrease to the current levels, respectively, with bold font indicating an over 30% change. Results for individuals whose simulated recommendations could not satisfy all concentration limits are considered non-comparable and shown as all-zero rows.

| | **Protein** (*E%*) | **Carboh.** (*E%*) | **Sucrose** (*E%*) | **Fiber** (*g/d*) | **SFA** (*E%*) | **MUFA** (*E%*) | **PUFA** (*E%*) | **Linol. acid** (*E%*) | **α-linol. acid** (*E%*) | **EPA** (*mg/d*) | **DHA** (*mg/d*) | **Chol.** (*mg/d*) | **Folate** (*μg/d*) | **Vit. D** (*μg/d*) | **Vit. C** (*mg/d*) |
|---|---|---|---|---|---|---|---|---|---|---|---|---|---|---|---|
| 42 | 15.4→ **0.0** [0.0;0.0] | 52.0→ **0.0** [0.0;0.0] | 5.8→ **0.0** [0.0;0.0] | 27.0→ **0.0** [0.0;0.0] | 11.6→ **0.0** [0.0;0.0] | 9.1→ **0.0** [0.0;0.0] | 4.8→ **0.0** [0.0;0.0] | 3.7→ **0.0** [0.0;0.0] | 0.7→ **0.0** [0.0;0.0] | 0.0→ **0.0** [0.0;0.0] | 0.1→ **0.0** [0.0;0.0] | 319→ **0** [0;0] | 341→ **0** [0;0] | 4.9→ **0.0** [0.0;0.0] | 121→ **0** [0;0] |
| 44 | 13.9→ 17.0 [11.3;19.9] | 44.7→ 53.6 [42.6;59.8] | 8.2→ **1.8** [0.1;6.2] | 17.6→ **28.6** [20.2;46.5] | 13.5→ **8.4** [7.1;9.9] | 11.4→ 15.9 [10.6;19.8] | 4.9→ **7.4** [5.1;9.9] | 3.6→ **5.6** [2.7;8.8] | 0.8→ **3.9** [1.9;5.0] | 0.1→ **2.0** [1.5;2.5] | 0.3→ **0.2** [0.0;0.5] | 292→ **110** [5;278] | 254→ **889** [647;997] | 7.6→ **87.0** [58.8;99.7] | 92→ **191** [79;430] |
| 48 | 16.4→ **0.0** [0.0;0.0] | 41.6→ **0.0** [0.0;0.0] | 4.0→ **0.0** [0.0;0.0] | 19.4→ **0.0** [0.0;0.0] | 16.4→ **0.0** [0.0;0.0] | 12.8→ **0.0** [0.0;0.0] | 5.2→ **0.0** [0.0;0.0] | 2.6→ **0.0** [0.0;0.0] | 1.2→ **0.0** [0.0;0.0] | 0.0→ **0.0** [0.0;0.0] | 0.1→ **0.0** [0.0;0.0] | 224→ **0** [0;0] | 217→ **0** [0;0] | 11.5→ **0.0** [0.0;0.0] | 68→ **0** [0;0] |
| 52 | 16.3→ 16.2 [10.7;19.8] | 45.8→ 50.8 [40.7;59.5] | 5.5→ **3.0** [0.1;8.7] | 17.0→ **29.9** [20.3;49.5] | 12.3→ **8.3** [7.1;9.8] | 10.4→ **17.1** [11.6;19.9] | 6.0→ 7.5 [5.1;9.9] | 3.5→ **5.1** [2.6;8.6] | 0.9→ **4.4** [3.1;5.0] | 0.0→ **1.6** [1.0;2.3] | 0.1→ **0.3** [0.0;0.8] | 185→ **115** [4;279] | 260→ **909** [702;997] | 3.5→ **38.5** [8.9;86.4] | 105→ **203** [80;468] |
| 55 | 24.2→ **0.0** [0.0;0.0] | 41.9→ **0.0** [0.0;0.0] | 2.9→ **0.0** [0.0;0.0] | 14.2→ **0.0** [0.0;0.0] | 9.0→ **0.0** [0.0;0.0] | 7.6→ **0.0** [0.0;0.0] | 4.2→ **0.0** [0.0;0.0] | 3.0→ **0.0** [0.0;0.0] | 0.5→ **0.0** [0.0;0.0] | 0.2→ **0.0** [0.0;0.0] | 0.5→ **0.0** [0.0;0.0] | 189→ **0** [0;0] | 167→ **0** [0;0] | 5.5→ **0.0** [0.0;0.0] | 84→ **0** [0;0] |
| 64 | 21.4→ **0.0** [0.0;0.0] | 45.6→ **0.0** [0.0;0.0] | 4.0→ **0.0** [0.0;0.0] | 16.5→ **0.0** [0.0;0.0] | 11.5→ **0.0** [0.0;0.0] | 10.3→ **0.0** [0.0;0.0] | 4.9→ **0.0** [0.0;0.0] | 3.4→ **0.0** [0.0;0.0] | 0.7→ **0.0** [0.0;0.0] | 0.1→ **0.0** [0.0;0.0] | 0.2→ **0.0** [0.0;0.0] | 306→ **0** [0;0] | 167→ **0** [0;0] | 6.1→ **0.0** [0.0;0.0] | 54→ **0** [0;0] |
| 69 | 18.0→ 14.6 [10.2;19.6] | 51.1→ 50.2 [40.7;59.4] | 11.5→ **3.9** [0.1;9.5] | 20.8→ **38.0** [21.3;54.1] | 12.2→ **8.6** [7.1;9.9] | 9.6→ 13.5 [10.1;19.1] | 3.9→ **7.7** [5.2;9.9] | 3.0→ **6.3** [2.8;8.9] | 0.6→ **1.8** [0.6;4.1] | 0.1→ **2.3** [2.0;2.5] | 0.3→ **0.1** [0.0;0.5] | 411→ **111** [4;282] | 346→ 351 [113;743] | 9.6→ **72.5** [19.7;99.4] | 264→ 255 [81;695] |
| 78 | 16.6→ 15.5 [10.5;19.7] | 44.8→ 47.4 [40.3;58.0] | 3.4→ **1.9** [0.1;6.3] | 23.3→ 25.1 [20.1;37.9] | 10.3→ 8.0 [7.0;9.7] | 9.6→ **16.9** [11.7;19.9] | 5.0→ 7.0 [5.1;9.8] | 4.0→ 4.6 [2.6;8.3] | 0.6→ **4.6** [3.8;5.0] | 0.0→ **2.3** [1.9;2.5] | 0.1→ 0.1 [0.0;0.5] | 261→ **75** [2;235] | 200→ **920** [736;997] | 3.6→ **46.0** [16.7;82.2] | 54→ **153** [77;321] |
| 96 | 17.2→ 17.7 [12.5;19.9] | 47.0→ 54.9 [44.6;59.9] | 8.5→ **1.5** [0.0;4.8] | 23.1→ 23.4 [20.1;31.3] | 12.3→ **7.9** [7.0;9.7] | 10.0→ **18.8** [15.7;20.0] | 4.5→ **8.2** [5.4;9.9] | 3.1→ 4.3 [2.5;7.8] | 0.5→ **4.8** [4.2;5.0] | 0.1→ **1.7** [1.4;2.1] | 0.3→ **0.1** [0.0;0.3] | 269→ **40** [2;130] | 305→ **960** [863;999] | 6.0→ **15.2** [7.7;32.2] | 174→ **114** [76;219] |

EPA = Eicosapentaenoic acid, DHA = Docosahexaenoic acid

The general observation from the results is that saturated fatty acid (SFA) intake should be reduced and replaced with monounsaturated (MUFA) and polyunsaturated (PUFA) fatty acids, particularly eicosapentaenoic acid (EPA), linoleic acid (LA), and alpha-lipoic acid (ALA). These recommendations align with Nordic Nutrition Recommendations 2023.

Although the diet inference model remains well-defined and produces intake recommendations even when one or more concentration limits cannot be met, such partially valid recommendations are not comparable with fully valid ones and are therefore omitted from the results and shown as all-zero rows. However, they remain informative to examine in concentration simulations (Figs. S5 and S7) and contributions of nutrients (Figs. 5 and 6). For example, in the Sysdimet dataset, participant 42 could not achieve a plasma cholesterol concentration below its upper limit simultaneously with the other concentrations in any simulated diet configuration, as illustrated in Fig. S5.

These partially valid recommendations may still hold value. In future work, concentration limits could be assigned a priority structure so that less critical limits can be selectively relaxed under controlled conditions.

#### Reasoning for Recommended High Intakes of Vitamins C and D

High personal recommendations for vitamin C and D intake are due to the estimated personal characteristics of these individuals (most notably subjects S18, S19, S104, S107, and S121). Figure S7 gives a more detailed view of the personal effects of these vitamins. In Sysdimet study, many subjects have problems meeting the upper limits of glucose and insulin concentrations, and the model uses the estimated decreasing effect of vitamin C to decrease these concentrations. These high intake recommendations are computational last resort for the inference model to reach the limits, and in a possible clinical guide, other options should be considered.

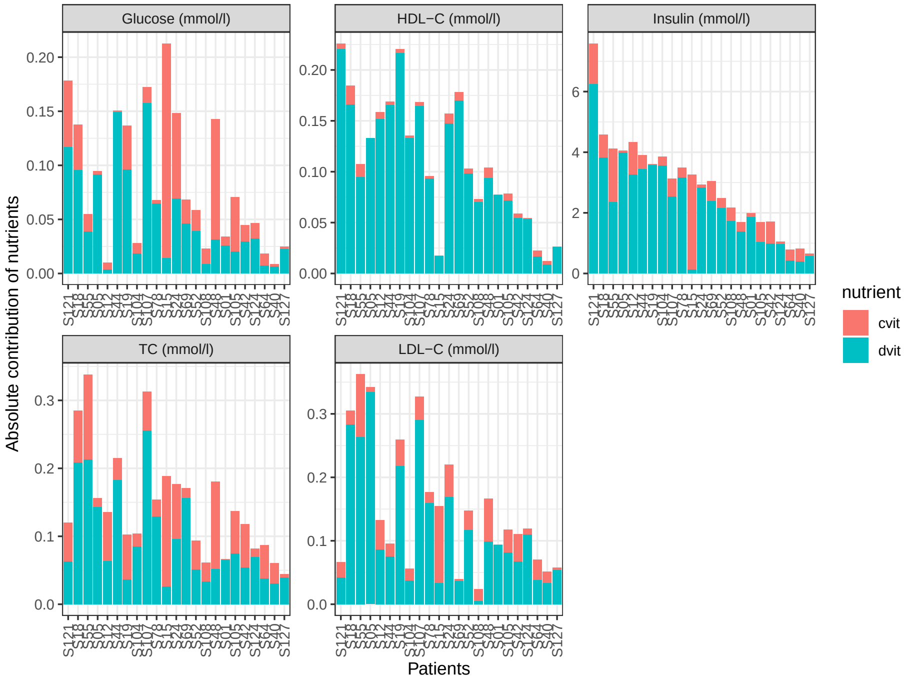


Figure S6: Elaborated effects of vitamins C and D in Sysdimet study. The figure is plotted with ggplot2 package for R language (v 3.3.5, https://ggplot2.tidyverse.org). (HDL-C = HDL cholesterol, LDL-C = LDL cholesterol, TC = Total cholesterol)

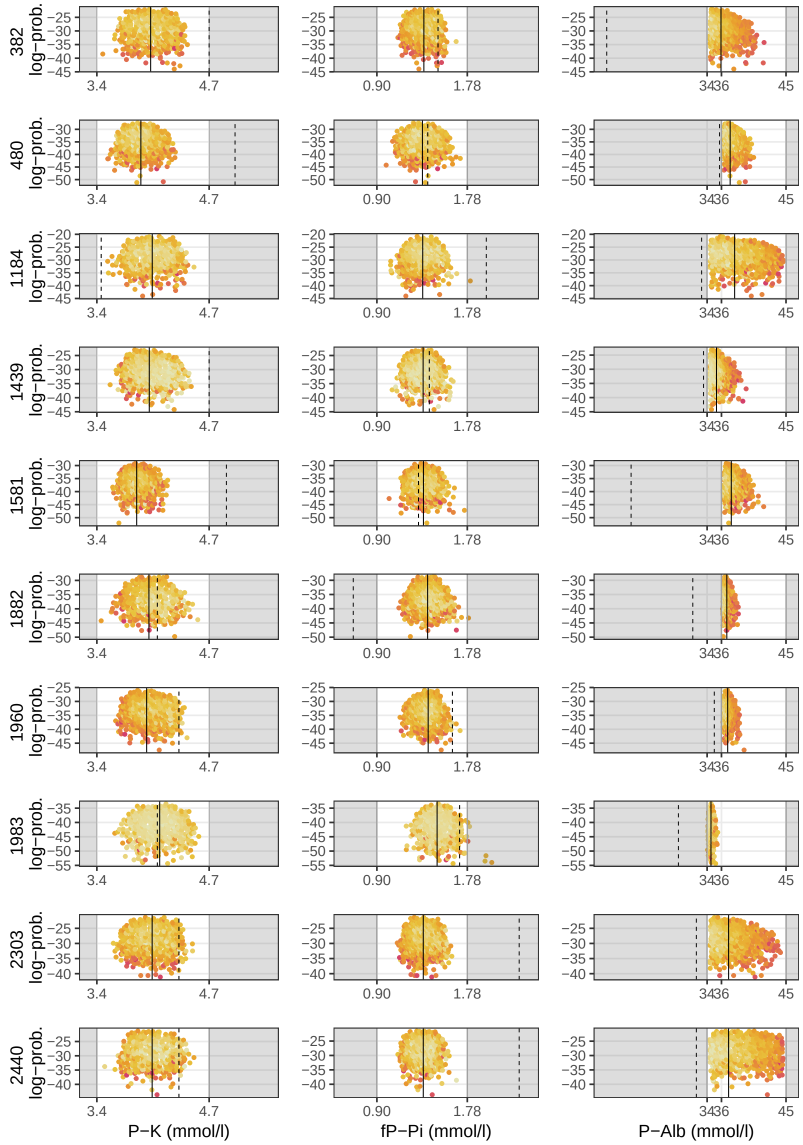


Figure S7: These are 10 of 35 renal patients whose current concentrations deviate from the normal ranges but could be improved with the personal diet recommendations. The white areas represent the normal concentration ranges, while the dashed vertical lines mark the patients' current concentrations, some of which fall within the gray areas. The solid lines indicate the concentration levels corresponding to the expected value of the joint multivariate recommendation distribution, where the nutrient intake and concentration predictions are jointly optimized. The figure is plotted with ggplot2 package for R language (v 3.3.5, https://ggplot2.tidyverse.org).

Table S10: Personally inferred intake recommendations for 30 of 35 renal patients with current concentrations out of the normal ranges. The table shows the current intake of nutrients and the recommended levels expected to produce normal concentrations and cause the smallest overall change in the current diet. Red and blue colors indicate an increase and decrease to the current levels, respectively, with bold font with underlining indicating an over 30% change. Results for individuals whose simulated recommendations could not satisfy all concentration limits are considered non-comparable and shown as all-zero rows.

| | **Protein** (*E%*) | **Carboh.** (*E%*) | **Sucrose** (*E%*) | **Fiber** (*g/d*) | **SFA** (*E%*) | **MUFA** (*E%*) | **PUFA** (*E%*) | **Potassium** (*mg/d*) | **Phosp.** (*mg/d*) | **Calcium** (*mg/d*) | **Salt** (*mg/d*) | **Water** (*ml/d*) | **Vit. D** (*μg/d*) |
|---|---|---|---|---|---|---|---|---|---|---|---|---|---|
| 382 | 15.0→ 15.7 [10.5;19.8] | 48.9→ 48.7 [40.4;59.2] | 12.1→ **5.4** [0.4;9.8] | 12.3→ **47.3** [34.7;54.7] | 10.6→ 8.6 [7.1;9.9] | 14.2→ 14.7 [10.2;19.6] | 6.8→ 7.7 [5.2;9.9] | 1838→ 2383 [1628;3541] | 694→ **2090** [1017;2928] | 367→ **1154** [438;2210] | 9275→ **3248** [1583;5537] | 1585→ 1361 [539;2411] | 5.4→ **61.8** [47.8;74.5] |
| 480 | 15.3→ 15.0 [10.4;19.7] | 55.0→ 54.3 [43.1;59.8] | 12.3→ **8.6** [5.3;10.0] | 18.2→ **33.4** [20.7;51.4] | 8.1→ 8.7 [7.1;9.9] | 11.7→ **18.5** [15.1;20.0] | 6.0→ 8.4 [5.6;9.9] | 2919→ **1981** [1612;2847] | 1025→ **1939** [767;2912] | 514→ 658 [410;1207] | 5640→ 5112 [3723;5734] | 2031→ **979** [522;1771] | 7.7→ **11.0** [7.6;18.9] |
| 1184 | 14.8→ 15.4 [10.4;19.8] | 34.0→ **49.7** [40.6;59.4] | 4.9→ 5.0 [0.3;9.7] | 20.2→ **43.7** [30.3;54.3] | 13.3→ **8.5** [7.1;9.9] | 21.7→ **15.0** [10.2;19.7] | 9.4→ 7.6 [5.2;9.9] | 3392→ 2620 [1651;3637] | 1100→ **1708** [576;2863] | 440→ **1376** [463;2423] | 7932→ **3561** [1610;5617] | 2245→ **1481** [559;2442] | 9.1→ **45.5** [29.2;62.7] |
| 1439 | 11.2→ **16.3** [10.8;19.9] | 46.3→ 51.9 [41.2;59.6] | 15.2→ **7.1** [1.8;9.9] | 12.2→ **41.3** [23.2;54.2] | 16.2→ **8.6** [7.1;9.9] | 13.8→ 16.7 [11.3;19.9] | 4.6→ **7.2** [5.1;9.7] | 1954→ 2333 [1622;3506] | 700→ **2428** [1473;2974] | 446→ 636 [407;1147] | 3798→ 4137 [1743;5687] | 1057→ 1365 [548;2398] | 1.8→ **47.8** [20.5;70.2] |
| 1581 | 12.1→ 14.4 [10.2;19.6] | 47.8→ 54.6 [43.8;59.8] | 16.4→ **8.3** [4.5;10.0] | 26.4→ 27.3 [20.3;39.3] | 15.0→ **8.8** [7.2;10.0] | 11.4→ **18.6** [15.6;20.0] | 4.2→ **8.0** [5.3;9.9] | 5546→ **1911** [1610;2628] | 2076→ **1105** [361;2172] | 903→ **575** [406;973] | 13800→ **5207** [3974;5730] | 3100→ **981** [523;1724] | 17.6→ **9.6** [7.6;14.4] |
| 1882 | 13.6→ 18.3 [14.5;19.9] | 41.1→ 50.5 [41.0;59.3] | 8.8→ 9.0 [6.5;10.0] | 27.6→ **46.6** [29.9;54.7] | 13.4→ **8.6** [7.1;9.9] | 17.4→ 18.2 [14.3;20.0] | 8.6→ 7.9 [5.3;9.9] | 3015→ **1917** [1608;2664] | 939→ **2634** [1925;2988] | 492→ 500 [403;730] | 7582→ **4930** [3111;5725] | 1571→ **937** [516;1857] | 8.2→ **47.9** [24.4;63.6] |
| 1960 | 14.1→ 18.3 [14.7;19.9] | 55.5→ 50.5 [40.7;59.5] | 9.2→ 8.7 [6.0;10.0] | 6.9→ **51.2** [43.0;54.9] | 6.4→ 8.6 [7.1;9.9] | 11.1→ **16.9** [11.2;19.9] | 8.5→ 7.8 [5.3;9.9] | 1119→ **2087** [1611;3174] | 320→ **1663** [620;2780] | 160→ **563** [405;950] | 2239→ **4788** [2795;5716] | 1488→ 1390 [548;2421] | 3.0→ **59.4** [41.1;75.9] |
| 1983 | 15.8→ 19.1 [16.9;20.0] | 55.4→ 44.0 [40.1;52.6] | 10.6→ 9.4 [8.0;10.0] | 14.6→ **38.3** [24.1;53.4] | 10.2→ 8.3 [7.1;9.9] | 8.9→ **18.1** [14.1;19.9] | 4.1→ **8.9** [6.3;10.0] | 1854→ 1831 [1607;2392] | 642→ **2419** [1546;2975] | 315→ **473** [402;646] | 4674→ 5287 [4225;5736] | 1331→ **701** [505;1168] | 4.3→ **22.5** [8.3;43.3] |
| 2303 | 14.1→ 15.7 [10.5;19.8] | 47.0→ 49.2 [40.5;59.1] | 14.5→ **5.1** [0.3;9.7] | 12.6→ **45.7** [32.6;54.4] | 15.1→ **8.6** [7.1;9.9] | 14.5→ 14.9 [10.3;19.6] | 5.9→ 7.7 [5.1;9.9] | 2921→ 2563 [1644;3634] | 938→ **1693** [626;2810] | 476→ **1414** [466;2430] | 3554→ 3549 [1630;5624] | 1766→ 1446 [540;2434] | 6.4→ **45.3** [30.8;59.7] |
| 2440 | 16.8→ 15.2 [10.3;19.7] | 48.8→ 49.5 [40.5;59.4] | 12.3→ **4.9** [0.3;9.7] | 14.7→ **40.2** [25.6;53.7] | 11.8→ **8.5** [7.1;9.9] | 11.9→ 14.9 [10.3;19.8] | 5.9→ 7.6 [5.2;9.9] | 3168→ 2678 [1659;3650] | 1236→ 1311 [385;2527] | 922→ **1458** [467;2441] | 5476→ **3697** [1606;5644] | 1821→ 1479 [556;2456] | 7.2→ **55.0** [34.3;77.2] |
| 2638 | 14.7→ 14.2 [10.2;19.4] | 39.4→ 54.1 [42.7;59.8] | 4.9→ **8.2** [4.2;10.0] | 16.7→ **26.9** [20.3;37.5] | 18.8→ **8.8** [7.2;10.0] | 16.0→ 18.5 [15.2;20.0] | 7.2→ 8.1 [5.4;9.9] | 3192→ **1940** [1610;2704] | 1091→ 1333 [385;2489] | 590→ 592 [407;1029] | 6554→ 5236 [4076;5733] | 1427→ 1141 [525;2085] | 7.0→ 10.0 [7.6;16.0] |
| 2714 | 14.8→ 16.1 [10.7;19.8] | 43.7→ 50.1 [40.7;59.3] | 12.0→ **6.1** [0.7;9.9] | 20.4→ **45.8** [32.0;54.6] | 13.3→ **8.5** [7.1;9.9] | 14.7→ 15.6 [10.4;19.8] | 6.7→ 7.6 [5.2;9.9] | 3043→ 2596 [1649;3647] | 1520→ 1930 [657;2931] | 935→ 942 [420;1930] | 11763→ **3983** [1715;5680] | 1804→ 1448 [556;2434] | 7.2→ **74.8** [41.8;97.7] |
| 2884 | 16.8→ 17.5 [12.4;19.9] | 40.1→ 50.5 [40.9;59.5] | 6.8→ 8.0 [3.5;9.9] | 10.9→ **50.8** [41.8;54.9] | 12.8→ **8.6** [7.1;9.9] | 13.8→ 16.5 [11.1;19.9] | 8.7→ 7.7 [5.2;9.9] | 1824→ 2234 [1622;3398] | 738→ **2082** [929;2936] | 416→ **630** [406;1158] | 4648→ 4506 [2170;5706] | 1246→ 1183 [527;2282] | 4.9→ **73.0** [54.3;89.4] |
| 2928 | 10.0→ **16.5** [11.1;19.8] | 60.4→ 51.7 [41.0;59.6] | 15.6→ **7.6** [3.2;9.9] | 16.6→ **47.4** [32.2;54.7] | 11.7→ **8.8** [7.2;9.9] | 6.9→ **17.6** [13.0;19.9] | 5.3→ **8.4** [5.6;9.9] | 1862→ 2174 [1620;3290] | 775→ 613 [311;1305] | 440→ 561 [405;935] | 4622→ 5230 [3968;5734] | 1427→ **925** [517;1667] | 6.5→ **11.0** [7.6;17.9] |
| 4025 | 15.1→ 14.2 [10.2;19.5] | 42.3→ 55.5 [45.3;59.9] | 9.7→ 8.7 [5.5;10.0] | 14.1→ **32.8** [20.8;51.2] | 10.5→ 8.9 [7.2;10.0] | 17.1→ 18.9 [16.3;20.0] | 9.8→ 8.3 [5.5;9.9] | 1485→ 1847 [1606;2447] | 637→ **1439** [504;2519] | 338→ **542** [404;881] | 3568→ **5339** [4379;5740] | 2451→ **970** [522;1726] | 7.1→ 9.1 [7.5;12.9] |
| 4710 | 19.4→ 18.0 [13.4;19.9] | 44.0→ 48.8 [40.4;58.9] | 14.3→ **8.6** [5.4;10.0] | 9.9→ **51.8** [44.3;54.9] | 10.7→ 8.5 [7.1;9.9] | 14.6→ 16.5 [11.0;19.9] | 6.5→ 7.9 [5.3;9.9] | 2160→ 2018 [1614;2934] | 736→ **1274** [380;2363] | 258→ **549** [405;905] | 4908→ 4780 [2608;5725] | 1487→ **909** [513;1812] | 5.1→ **76.8** [60.4;91.7] |
| 4741 | 16.0→ 16.7 [11.1;19.9] | 36.7→ **53.7** [42.5;59.8] | 6.8→ 8.0 [3.5;9.9] | 12.4→ **38.8** [23.3;53.0] | 14.9→ **8.7** [7.1;9.9] | 17.7→ 17.4 [12.0;19.9] | 8.2→ 6.8 [5.1;9.6] | 2232→ 2078 [1615;3097] | 888→ **2624** [1850;2987] | 447→ 564 [406;942] | 6264→ **4374** [1959;5709] | 1095→ 1233 [526;2362] | 9.0→ **55.3** [32.2;77.6] |
| 4903 | 18.8→ 16.0 [10.6;19.8] | 31.2→ **50.2** [40.7;59.5] | 5.2→ 6.3 [0.8;9.9] | 19.4→ **43.1** [26.1;54.4] | 16.9→ **8.5** [7.1;9.9] | 18.4→ 15.7 [10.6;19.8] | 7.8→ 7.6 [5.2;9.9] | 3630→ **2571** [1640;3626] | 1518→ 1890 [673;2903] | 1111→ **764** [413;1456] | 8468→ **3979** [1775;5674] | 2403→ **1381** [540;2403] | 9.1→ **39.1** [13.2;63.6] |
| 4962 | 20.6→ 18.2 [13.9;19.9] | 49.0→ 47.9 [40.3;58.5] | 7.4→ 8.3 [4.6;10.0] | 21.0→ **52.0** [45.0;54.9] | 8.1→ 8.5 [7.1;9.9] | 9.9→ **15.9** [10.6;19.8] | 7.7→ 8.3 [5.4;9.9] | 2822→ 2236 [1618;3403] | 1220→ 1057 [353;2100] | 451→ 613 [405;1077] | 7221→ **4716** [2452;5714] | 1718→ **1044** [514;2169] | 1.4→ **68.6** [50.8;83.9] |
| 4999 | 17.2→ 15.8 [10.5;19.8] | 44.5→ 54.3 [43.4;59.7] | 10.7→ **7.9** [3.6;9.9] | 32.8→ **48.5** [34.1;54.8] | 10.7→ 9.1 [7.5;10.0] | 14.0→ 18.5 [15.2;20.0] | 7.8→ 8.2 [5.4;9.9] | 4211→ **2003** [1611;2904] | 1476→ 1194 [356;2470] | 553→ 574 [404;997] | 7176→ **5379** [4440;5740] | 2513→ **1568** [619;2393] | 12.0→ 11.7 [7.7;20.3] |

Table S10: (Continued) Personally inferred intake recommendations for 30 of 35 renal patients with current concentrations out of the normal ranges. The table shows the current intake of nutrients and the recommended levels expected to produce normal concentrations and cause the smallest overall change in the current diet. Red and blue colors indicate an increase and decrease to the current levels, respectively, with bold font indicating an over 30% change. Results for individuals whose simulated recommendations could not satisfy all concentration limits are considered non-comparable and shown as all-zero rows.

| | Protein (*E%*) | Carboh. (*E%*) | Sucrose (*E%*) | Fiber (*g/d*) | SFA (*E%*) | MUFA (*E%*) | PUFA (*E%*) | Potassium (*mg/d*) | Phosp. (*mg/d*) | Calcium (*mg/d*) | Salt (*mg/d*) | Water (*ml/d*) | Vit. D (*µg/d*) |
|---|---|---|---|---|---|---|---|---|---|---|---|---|---|
| 5181 | 13.4→ 15.8 [10.5;19.8] | 48.9→ 53.7 [42.6;59.8] | 14.7→ **7.8** [3.2;9.9] | 9.2→ **27.8** [20.4;38.3] | 16.3→ **8.7** [7.1;9.9] | 10.9→ **17.7** [12.9;19.9] | 4.0→ **6.6** [5.1;9.3] | 2190→ 2188 [1623;3293] | 1029→ **2777** [2320;2993] | 873→ **562** [404;914] | 2914→ **4270** [1877;5702] | 1275→ 1440 [552;2423] | 8.1→ **36.1** [18.0;55.9] |
| 5751 | 16.6→ 17.3 [11.9;19.9] | 34.6→ 47.5 [40.3;58.2] | 2.6→ **7.9** [3.4;9.9] | 8.8→ **45.7** [27.0;54.6] | 15.5→ **8.5** [7.1;9.9] | 20.5→ **15.6** [10.4;19.7] | 8.9→ 8.0 [5.3;9.9] | 2786→ 2260 [1623;3396] | 984→ 1090 [349;2199] | 627→ 612 [408;1060] | 2701→ **4463** [2083;5699] | 1494→ **1044** [519;2143] | 9.6→ **61.3** [35.7;79.4] |
| 5775 | 12.2→ 12.9 [10.1;18.3] | 40.0→ **57.7** [52.2;59.9] | 8.5→ 9.1 [7.0;10.0] | 14.9→ **44.1** [27.3;54.6] | 18.9→ **9.4** [8.2;10.0] | 14.9→ 19.4 [18.1;20.0] | 6.2→ 8.2 [5.5;9.9] | 2882→ **1722** [1603;2011] | 1064→ 1405 [488;2215] | 608→ 468 [402;641] | 5460→ 5568 [5096;5745] | 1496→ 1811 [896;2460] | 10.3→ 8.2 [7.5;10.1] |
| 5981 | 13.4→ 18.1 [14.1;19.9] | 48.9→ 45.5 [40.2;55.8] | 14.7→ **8.3** [4.5;10.0] | 9.2→ **49.7** [37.5;54.9] | 16.3→ **8.4** [7.1;9.9] | 10.9→ 15.2 [10.4;19.7] | 4.0→ **8.5** [5.7;10.0] | 2190→ 2312 [1627;3483] | 1029→ 1005 [329;2203] | 873→ **655** [409;1163] | 2914→ **4727** [2634;5721] | 1275→ **952** [512;1921] | 8.1→ **74.2** [51.3;91.0] |
| 6428 | 16.5→ 16.1 [10.6;19.8] | 40.9→ 48.2 [40.4;58.9] | 6.5→ 5.6 [0.4;9.8] | 21.2→ **49.5** [39.0;54.8] | 12.6→ **8.6** [7.1;9.9] | 15.4→ 14.7 [10.2;19.7] | 7.0→ 7.9 [5.2;9.9] | 4105→ **2347** [1627;3506] | 1085→ **1842** [821;2830] | 480→ **1361** [449;2424] | 7748→ **3375** [1584;5561] | 2273→ **1382** [545;2414] | 5.7→ **57.3** [45.2;69.2] |
| 6591 | 17.0→ 15.4 [10.4;19.8] | 37.6→ 49.5 [40.5;59.4] | 6.5→ 5.1 [0.3;9.7] | 18.5→ **44.8** [31.7;54.4] | 14.3→ **8.6** [7.1;9.9] | 15.6→ 14.8 [10.2;19.8] | 8.4→ 7.5 [5.2;9.9] | 3218→ 2600 [1638;3641] | 1237→ 1477 [480;2676] | 496→ **1481** [500;2427] | 7367→ **3570** [1619;5604] | 2268→ **1489** [558;2434] | 15.4→ **49.3** [33.5;67.5] |
| 6770 | 12.2→ **0.0** [0.0;0.0] | 43.3→ **0.0** [0.0;0.0] | 8.8→ **0.0** [0.0;0.0] | 21.2→ **0.0** [0.0;0.0] | 14.2→ **0.0** [0.0;0.0] | 17.0→ **0.0** [0.0;0.0] | 8.7→ **0.0** [0.0;0.0] | 4194→ **0** [0;0] | 1334→ **0** [0;0] | 640→ **0** [0;0] | 12065→ **0** [0;0] | 1850→ **0** [0;0] | 4.9→ **0.0** [0.0;0.0] |
| 7086 | 16.8→ 14.3 [10.3;19.5] | 44.0→ 56.4 [48.4;59.9] | 11.4→ 8.9 [6.4;10.0] | 18.5→ **38.5** [22.7;53.4] | 17.6→ **9.1** [7.3;10.0] | 11.3→ **19.1** [17.0;20.0] | 3.5→ **8.5** [5.7;10.0] | 3362→ **1800** [1605;2275] | 1160→ 1358 [389;2569] | 950→ **500** [402;754] | 4878→ 5443 [4702;5739] | 2134→ **1035** [534;1853] | 4.7→ **9.0** [7.5;12.5] |
| 7370 | 15.8→ 18.5 [15.0;20.0] | 35.5→ 42.6 [40.1;48.6] | 8.6→ 8.1 [3.6;9.9] | 17.0→ 22.3 [20.1;27.8] | 14.3→ **8.0** [7.0;9.6] | 19.8→ **15.1** [10.3;19.7] | 9.6→ 9.2 [7.4;10.0] | 3224→ 2704 [1654;3653] | 1137→ 1580 [755;2387] | 554→ 537 [404;887] | 8465→ **4717** [2467;5718] | 1609→ **894** [512;1842] | 19.9→ **47.5** [21.9;74.2] |
| 8080 | 13.7→ 18.2 [13.9;19.9] | 38.9→ 47.0 [40.3;57.7] | 4.5→ **8.7** [5.6;10.0] | 18.0→ **47.6** [30.7;54.8] | 13.2→ **8.5** [7.1;9.9] | 19.5→ 16.3 [10.8;19.9] | 9.4→ 8.1 [5.3;9.9] | 2631→ 2050 [1613;3074] | 1006→ 1202 [375;2276] | 484→ 537 [405;855] | 7678→ **4820** [2801;5719] | 1276→ **894** [513;1760] | 8.7→ **71.6** [38.9;93.3] |
| 9093 | 10.4→ **15.7** [10.5;19.8] | 46.1→ 49.0 [40.5;59.3] | 9.9→ **5.1** [0.3;9.7] | 12.6→ **46.9** [34.3;54.7] | 13.3→ **8.5** [7.1;9.9] | 13.8→ 14.8 [10.2;19.7] | 12.6→ **7.7** [5.2;9.9] | 2355→ 2550 [1643;3611] | 768→ **1471** [460;2597] | 470→ **1488** [486;2449] | 4958→ **3583** [1607;5631] | 1178→ 1447 [556;2429] | 9.4→ **52.9** [37.4;68.1] |
| 9464 | 15.2→ 18.6 [15.4;19.9] | 36.1→ 46.1 [40.2;56.7] | 10.0→ 8.8 [5.9;10.0] | 20.9→ **44.8** [26.8;54.6] | 23.6→ **8.4** [7.1;9.9] | 15.3→ 16.7 [11.0;19.9] | 4.5→ **8.5** [5.7;10.0] | 3424→ **2079** [1612;3146] | 1927→ 2076 [1033;2916] | 1504→ **513** [403;784] | 11639→ **4942** [3127;5726] | 2495→ **873** [509;1701] | 11.4→ **29.1** [15.8;39.4] |

### Section "5.3 Personally Inferred Dietary Recommendations"

These tables present the contributions of nutrients to concentrations on a normalized scale, allowing all nutrients to be compared regardless of their original units. The tables show the minimum and maximum contributions among the patients to highlight the variance. Each contribution reflects the product of the effect of a nutrient and the level of intake, showing how changes in intake influence concentrations. In particular, a nutrient with an increasing effect on concentration may have a negative net contribution if intake decreases.

Table S11: Most personally decreasing and increasing nutrient contributions in the Sysdimet dataset, ordered by decreasing average contribution.

| | **HDL-C** | | **LDL-C** | | **Insulin** | | **Total Chol.** | | **Glucose** | |
|---|---|---|---|---|---|---|---|---|---|---|
| **Nutrient** | Min | Max | Min | Max | Min | Max | Min | Max | Min | Max |
| Linoleic acid | 0.05 | 0.05 | -0.44 | -0.40 | -4.08 | -3.72 | -0.37 | -0.34 | 0.21 | 0.23 |
| Folate | -0.09 | 0.00 | -0.28 | -0.01 | -6.28 | -0.27 | -0.52 | -0.03 | -0.01 | 0.21 |
| EPA | 0.04 | 0.41 | -0.62 | -0.05 | -0.37 | 4.33 | 0.08 | 0.76 | -0.89 | -0.12 |
| Vit. D | 0.01 | 0.22 | -0.07 | 0.33 | 0.12 | 6.25 | -0.18 | 0.26 | -0.16 | 0.00 |
| Alpha-linolenic acid | -0.05 | 0.00 | 0.01 | 0.40 | 0.09 | 3.69 | 0.01 | 0.35 | -0.20 | 0.00 |
| MUFA | -0.11 | 0.01 | -0.15 | -0.04 | -0.27 | 1.94 | -0.20 | -0.04 | 0.02 | 0.08 |
| Vit. C | -0.02 | 0.01 | 0.00 | 0.12 | -0.66 | 3.14 | 0.00 | 0.16 | -0.20 | 0.05 |
| Cholesterol | -0.01 | 0.00 | -0.11 | -0.01 | -1.06 | -0.36 | -0.10 | -0.02 | -0.06 | 0.00 |
| SFA | 0.02 | 0.03 | -0.09 | -0.02 | -0.27 | 0.69 | -0.12 | -0.01 | 0.02 | 0.06 |
| Protein | 0.01 | 0.06 | 0.00 | 0.06 | 0.04 | 0.40 | 0.00 | 0.12 | -0.04 | 0.01 |
| Sucrose | 0.00 | 0.02 | -0.10 | 0.02 | -0.44 | -0.11 | -0.10 | 0.00 | -0.01 | 0.03 |
| PUFA | -0.01 | 0.01 | -0.04 | 0.06 | -0.45 | -0.05 | -0.05 | 0.03 | -0.03 | 0.01 |
| Carbohydrates | -0.05 | -0.01 | 0.00 | 0.02 | -0.09 | 0.50 | -0.04 | -0.01 | -0.02 | 0.00 |
| Fiber | 0.00 | 0.01 | 0.00 | 0.11 | -0.24 | 0.07 | 0.00 | 0.13 | -0.05 | 0.00 |
| DHA | -0.01 | 0.01 | -0.04 | 0.07 | -0.06 | 0.11 | -0.02 | 0.03 | -0.06 | 0.13 |

DHA = Docosahexaenoic acid; EPA = Eicosapentaenoic acid; MUFA = Monounsaturated fatty acid; PUFA = Polyunsaturated fatty acid

Table S12: Most personally decreasing and increasing nutrient contributions in the Dialysis dataset, ordered by decreasing average contribution.

| | **Potassium** | | **Phosphate** | | **Albumin** | |
|---|---|---|---|---|---|---|
| **Nutrient** | Min | Max | Min | Max | Min | Max |
| Vit. D | -4.70 | 0.37 | -0.16 | 1.26 | -0.67 | 10.77 |
| Fiber | 0.03 | 2.72 | -1.12 | -0.15 | -3.29 | 0.64 |
| Phosphorus | -0.06 | 0.48 | -0.18 | 1.00 | -0.49 | 2.57 |
| Salt | -0.24 | 0.91 | 0.00 | 0.21 | -1.23 | 0.66 |
| Potassium | -0.01 | 0.22 | -0.07 | 0.14 | -0.02 | 1.65 |
| Calcium | -0.24 | 0.03 | -0.39 | 0.04 | -3.40 | 0.34 |
| MUFA | 0.00 | 0.21 | 0.00 | 0.18 | 0.00 | 1.81 |
| Carbohydrates | -0.03 | 0.30 | -0.02 | 0.20 | -0.30 | 1.03 |
| Water | -0.19 | 0.00 | -0.28 | -0.01 | -0.10 | 2.09 |
| SFA | -0.53 | -0.25 | -0.14 | 0.04 | -0.25 | 0.56 |
| Sucrose | -0.07 | 0.14 | 0.00 | 0.07 | -1.11 | 0.04 |
| PUFA | -0.15 | 0.04 | -0.13 | 0.05 | -0.10 | 0.61 |
| Protein | -0.04 | 0.05 | -0.10 | 0.08 | -0.11 | 0.52 |

MUFA = Monounsaturated fatty acid; PUFA = Polyunsaturated fatty acid; SFA = Saturated fatty acid

The reported contributions represent the net effect of each nutrient on the modeled concentrations, calculated as the product of its personal effect and the corresponding intake level (effect $\times$ intake change). Positive values indicate that the nutrient's current or recommended intake raises the concentration, whereas negative values indicate a lowering influence after all interactions and compensations are accounted for.

### Section "6 Discussion"

Table S13: Comparison of the inferred nutrient effects and intake recommendations with nutrition literature. Table compares the diet inference results of this study and nutrition literature. The results of the nutrient effect are mostly found in Tables S7 and S8. Some weaker effects are not within the top 20 strongest effects in the tables and are reported only here.

| Nutrient | Effect | Recommendation | Literature |
|---|---|---|---|
| Sucrose | Increases plasma albumin concentration with personal effects ranging from 0.65 to 2.31; Mixed effects between individuals on potassium (-0.12 to 0.10) and insulin (0.16 to 0.33) | Decrease intake to maintain albumin within target levels; contributes to balancing albumin and potassium concentrations | Limited literature available on sucrose to albumin effect |
| Protein | Decreases insulin (-0.05 to -0.37); Mainly increases albumin (-0.05 to 0.45) | Adjust intake towards 15 E% recommendation; Contributes to reaching albumin lower limit in renal patients | Increases albumin(2); decreases insulin (3), HDL-C and TC (4) |
| Carbohydrates | Mixed effects on insulin (-0.23 to 0.49) and albumin (-0.44 to 0.71); Increases potassium (0.10 to 0.25) | Increase intake towards 50 E% recommendation; Contributes to decreasing insulin and increasing albumin and potassium | Decreases insulin for low-GI carbohydrates (5); can increase albumin in dialysis patients (6) |
| Saturated fatty acids (SFA) | Mixed effects on insulin (-0.58 to 0.25) and albumin (-0.46 to 0.20). Increases potassium (0.20 to 0.41) | Decrease intake towards 8.5 E% recommendation; may contribute to albumin increase in some patients | Decrease in SFA reduces serum insulin (7) |
| Monounsaturated fatty acids (MUFA) | Mixed effects on insulin (-0.09 to 0.81); Increases albumin (0.17 to 1.39) | Increase intake towards 15 E% recommendation; cCntributes to insulin reduction and albumin increase | MUFA intake linked to improved insulin sensitivity and albumin levels; supports increasing towards 15 E% recommendation (8) |
| Polyunsaturated fatty acids (PUFA) | Decreases insulin (-0.04 to -0.42); Mixed effects on albumin (-0.41 to 1.32) | Increase intake to reduce insulin; Contributes to managing albumin concentration | PUFA intake associated with reduced insulin levels and variable effects on albumin (8) |
| Fiber | Mixed effects on insulin (-0.24 to 0.13) and albumin (-1.26 to 0.12) | Increase intake towards 30 g/d recommendation; Contributes to decreasing phosphate and increasing potassium | Can increase albumin as inflammation decreases (9) |
| Vitamin D | Increases insulin (0.13 to 0.55); Mixed effects on albumin (-0.91 to 1.15) and potassium (-0.43 to 0.05); Decreases TC (-0.04 to -0.09), and glucose (-0.03 to -0.07) | Important in boosting albumin above lower limit; contributes to decreasing potassium and glucose (see Fig. 6) | Increase in albumin concentration reported for renal patients (10) |
| Calcium | Decreases albumin (-0.38 to -1.20); Mixed effects on plasma phosphate (-0.13 to 0.10) | Increase toward 800 mg/d to lower phosphate while monitoring albumin decrease | Indirect correlations between calcium intake, and both phosphate and albumin concentrations are reported. |
| Potassium | Primarily decreases albumin (-0.39 to -1.77) | Adjust intake based on albumin impact; Contributes to balancing potassium and albumin | |
| Salt | Mixed effects on albumin (-0.62 to 2.55) and potassium (-0.85 to 0.41) | "Moderate or limit salt to help keep phosphate within limits; individualized targets in recommendations span 2–5 g/d | High potassium intake is inversely associated with the development of albuminuria in chronic kidney disease patients (11). |

E% = Energy percentage; SFA = Saturated fatty acids; MUFA = Monounsaturated fatty acids; PUFA = Polyunsaturated fatty acids; TC = Total cholesterol

Table S13: (Continues) Comparison of the inferred nutrient effects and intake recommendations with nutrition literature. Table compares the diet inference results of this study and nutrition literature. The results of the nutrient effect are mostly found in Tables S7 and S8. Some weaker effects are not within the top 20 strongest effects in the tables and are reported only here.

| Nutrient | Effect | Recommendation | Literature |
|---|---|---|---|
| Phosphorus | Mixed effects on albumin (-0.29 to 0.97); increases phosphate (0.14 to 0.22) | Increase intake to reach albumin lower limit; contributes to balancing potassium | Diverse literature on phosphorous increasing phosphate concentration on renal patients. |
| Water | Mixed effects on albumin (-1.54 to 0.20) | Maintain general recommendation of 1500 ml/d; contributes to increasing albumin | |
| Linoleic acid | Increases insulin (7.56 to 7.60), LDL-C (0.81), and TC (0.70); decreases glucose (-0.43) | Decrease intake to contribute to lowering insulin, LDL-C, and TC | Diverse literature on increasing insulin. |
| Alpha-linolenic acid | Decreases insulin (-7.68 to -7.74), LDL-C (-0.83 to -0.84), and TC (-0.72 to -0.73); increases glucose (0.41) | Increase intake to contribute to lowering LDL-C and TC | Significantly decreases triglycerides, TC, LDL-C (12); High ALA intake is associated with a lower prevalence of insulin resistance (13) |
| Cholesterol | Increases insulin (0.32 to 0.68); Increases LDL-C (0.01 to 0.08) and TC (0.01 to 0.06) | Decrease intake to contribute to lowering insulin, LDL-C, and TC | Increases LDL-C and TC (8) |
| Folate | Decreases insulin (-0.31 to -1.06) | Adjust intake to balance insulin levels; contributes to insulin reduction | Decreases insulin (8) |
| Vitamin C | Mixed effects on insulin (-0.29 to 1.03) and glucose (-0.04 to 0.06) | Increase for individuals in whom the personal effect on glucose is negative (see Fig. 5) | Increases HDL-C (8; 14) |
| EPA | Decreases glucose (-0.04 to -0.06); Minor mixed effects on insulin (-0.11 to 0.30); HDL-C (0.00 to 0.03), and TC (0.02 to 0.05) | Increase intake to balance insulin, HDL-C, and TC | Decreases triglyceride levels (8; 14) |
| DHA | Mixed effects on insulin (-0.02 to 0.15); Increases glucose (0.09 to 0.10) | Adjust intake to balance glucose and insulin; Net effect is personal | Decreases triglyceride levels (8; 14) |

EPA = Eicosapentaenoic acid; DHA = Docosahexaenoic acid; LDL-C = Low-density lipoprotein cholesterol; HDL-C = High-density lipoprotein cholesterol; TC = Total cholesterol

[4] Anderson JW, Johnstone B, Cook-Newell M. Meta-analysis of the effects of soy

protein intake on serum lipids.. *The New England journal of medicine.* 1995;333 5:276-82. doi: 10.1056/NEJM199508033330502 40
